# Intramuscular Versus Intravenous SARS-CoV-2 Neutralizing Antibody Sotrovimab for Treatment of COVID-19 (COMET-TAIL): A Randomized Non-inferiority Clinical Trial

**DOI:** 10.1101/2023.03.21.23287410

**Authors:** Adrienne E. Shapiro, Elias Sarkis, Jude Acloque, Almena Free, Yaneicy Gonzalez-Rojas, Rubaba Hussain, Erick Juarez, Jaynier Moya, Naval Parikh, David Inman, Deborah Cebrik, Ahmed Nader, Nadia Noormohamed, Qianwen Wang, Andrew Skingsley, Daren Austin, Amanda Peppercorn, Maria L. Agostini, Sergio Parra, Sophia Chow, Erik Mogalian, Phillip S. Pang, David K. Hong, Jennifer E. Sager, Wendy W. Yeh, Elizabeth L. Alexander, Leah A. Gaffney, Anita Kohli

**Author notes:** Correspondence to: Dr Anita Kohli, Arizona Clinical Trials/Arizona Liver Health, 2201 West Fairview St, Chandler AZ 85248. Alternate corresponding author: Dr Leah Gaffney, Vir Biotechnology, Inc., 499 Illinois Street, Suite 500, San Francisco, CA 94158.

## Abstract

**Background:** Convenient administration of coronavirus disease 2019 (COVID-19) treatment in community settings is desirable. Sotrovimab is a pan-sarbecovirus dual-action monoclonal antibody formulated for intravenous (IV) or intramuscular (IM) administration for early treatment of mild/moderate COVID-19.

**Methods:** This phase 3, randomized, multicenter, open-label study tested non-inferiority of IM to IV administration using a 3.5% absolute non-inferiority margin. From June to August 2021, patients aged ≥12 years with COVID-19, not hospitalized or receiving supplemental oxygen, and at high risk for progression were randomized 1:1:1 to a single 500-mg IV sotrovimab infusion or 500-mg or 250-mg IM sotrovimab injection. The primary composite endpoint was progression to all-cause hospitalization for >24 hours for acute management of illness or all-cause death through day 29.

**Results:** Sotrovimab 500 mg IM was non-inferior to 500 mg IV: 10/376 (2.7%) participants in the sotrovimab 500-mg IM group versus 5/378 (1.3%) in the sotrovimab 500-mg IV group met the primary endpoint (absolute adjusted risk difference: 1.06% [95% confidence interval [CI]: −1.15%, 3.26%]). The CI upper limit was lower than the prespecified non-inferiority margin of 3.5%. 250-mg IM group enrollment was discontinued early because a greater proportion of hospitalizations was seen in that group versus the 500-mg groups. Serious adverse events occurred in <1% to 2% of participants across groups. Four participants experienced serious disease related events and died (500 mg IM: 2/393 [<1%]; 250 mg IM: 2/195 [1%]).

**Conclusions:** Sotrovimab 500-mg IM injection was well tolerated and non-inferior to IV administration. IM administration could expand outpatient treatment access for COVID-19.

**Registration:** ClinicalTrials.gov Identifier: NCT04913675

**Key Points:** Sotrovimab 500-mg IM was non-inferior to sotrovimab 500-mg IV for treatment of mild/moderate COVID-19 in high-risk patients, measured by all-cause hospitalization >24h or death through day 29, and was well-tolerated. Sotrovimab IM should provide easier outpatient access to COVID-19 treatment.

## Introduction

Severe coronavirus disease 2019 (COVID-19) is associated with a substantial burden on healthcare resources, particularly among unvaccinated patients and patients with risk factors for progression to severe disease. To prevent disease progression and hospitalization in high-risk patients with mild to moderate COVID-19, several monoclonal antibodies (mAbs) have been authorized for early treatment [1-6]. Sotrovimab targets a conserved epitope in the severe acute respiratory syndrome coronavirus 2 (SARS-CoV-2) spike protein, demonstrating potent neutralizing activity against wild-type and most SARS-CoV-2 variants, including B.1.617.2 (Delta) and B.1.1.529 (Omicron, BA.1 and BA1.1) [7-10]. Observational clinical data show sotrovimab was clinically effective against Omicron BA.1, BA.2 and BA.5 [11-15]. A moderate decrease in *in vitro* neutralization activity has been reported for sotrovimab against Omicron BA.2, BA.2.12.1, BA.4, and BA.5 authentic virus isolates (15.7-, 25.1-, 48.4- and 21.6-fold change in IC_50_ relative to wild type, respectively) [7, 16]. Although sotrovimab was not utilized in the US starting with the emergence of Omicron BA.2 due to these moderate decreases in activity, sotrovimab continued to be used for treatment of high-risk patients with COVID-19 in other countries, including the United Kingdom, Germany and Japan, which is supported by recent observational clinical data suggesting continued clinical effectiveness through Omicron BA.2 and BA.5 waves [12-15]. Understanding the feasibility of an intramuscular (IM) route of administration for sotrovimab may inform development of future anti-SARS-CoV-2 mAbs, ensuring greater feasibility of dosing and broader access.

Available mAbs for early treatment require IV infusion, which might be limited by requirements for infrastructure, staffing, isolation, and infection control [17]. There remains a need for treatments to prevent COVID-19 progression and decrease barriers for their administration. Sotrovimab has been formulated for IV or IM administration; IM administration would allow greater access to therapy and decrease healthcare burden.

The phase 3 COMET-ICE trial demonstrated the efficacy and safety of IV sotrovimab in patients with mild to moderate COVID-19 at high risk for disease progression from August 2020 to March 2021 [4, 5]. In the primary analysis (n=1057), sotrovimab indicated a statistically significant reduction in hospitalization for >24 hours for acute management of any illness or death due to any cause through day 29 in the sotrovimab-treated group versus placebo group (adjusted relative risk reduction, 79%; 95% confidence interval [CI], 91%-50%; p<0.001) [5]. IV sotrovimab was well tolerated with no unanticipated safety signals.

The COMET-TAIL study evaluated efficacy, safety, and tolerability of IM sotrovimab versus IV sotrovimab in high-risk patients for the treatment of mild to moderate COVID-19.

## Methods

### Study design

This phase 3, randomized, multicenter, open-label study was designed as a non-inferiority study, with a 3.5% non-inferiority margin, and was not placebo-controlled based on clinical efficacy of IV sotrovimab [4, 5] and endorsement of mAbs in treatment guidelines at the time of the study [18, 19]. A 3.5% non-inferiority margin was chosen based on feedback and scientific reasoning in collaboration with the US FDA. IM doses of 250 mg and 500 mg were selected to ensure sotrovimab concentrations in lung were maintained at or above levels anticipated to be neutralizing for the duration of the treatment window. Enrollment occurred from June 2021 through August 2021 when Delta was the predominant circulating variant.

The study was conducted in accordance with the consensus ethical principles derived from the Declaration of Helsinki and Council for International Organizations of Medical Sciences International Ethical Guidelines, applicable International Council for Harmonisation Good Clinical Practice guidelines, and applicable laws and regulations. Ethics approval was obtained from institutional review boards and ethics committees. Written informed consent/assent was provided by all participants.

### Participants

Eligible patients were aged ≥12 years at time of consent and at high risk for progression of COVID-19, including age ≥55 years and presence of comorbidities (e.g., diabetes, obesity, chronic kidney disease, congenital heart disease, congestive heart failure, chronic lung diseases, sickle cell disease, neurodevelopmental disorders, immunosuppressive disease or receiving immunosuppressive medications, or chronic liver disease). Initially, eligibility was considered independent of vaccination status. The protocol was amended on June 29, 2021, to exclude fully vaccinated immunocompetent participants (defined as those with at least 14 days since receiving the final dose in a COVID-19 vaccine series) because they may have reduced rates of progression (protocol amendment 2).

Participants had a positive SARS-CoV-2 test result by any validated diagnostic test (e.g., reverse transcriptase-polymerase chain reaction [RT-PCR], antigen-based testing on any specimen type), oxygen saturation ≥94% while on room air, and COVID-19 symptoms. Eligible participants received sotrovimab ≤7 days from the onset of symptoms.

Individuals who were hospitalized or likely to require hospitalization within 24 hours (as assessed by the investigator) and those with severe COVID-19 (i.e., shortness of breath at rest, respiratory distress, or requiring supplemental oxygen for COVID-19) were excluded.

### Randomization and intervention

Participants were randomly assigned 1:1:1 to receive a single 500-mg IV infusion or IM injection of sotrovimab (500 mg or 250 mg), with stratification based on age (12-17, 18-64, and ≥65 years), COVID-19 vaccination history (receipt of any COVID-19 vaccine dose), and geographic region.

After 15-minute IV infusion or IM injection, participants were monitored for 30 minutes, vital signs were measured every 15 minutes, and solicited assessment of injection-site reactions (IM only) occurred at 15 and 30 minutes. Participants were monitored for 36 weeks on an outpatient basis with collection of nasopharyngeal swabs for virology, blood draws for pharmacokinetic (PK) sampling, and safety laboratory tests (**Figure 1**). Sparse PK samples were collected through week 24 and sotrovimab serum concentrations were determined using an electrochemiluminescent method validated on the Meso Scale Discovery (Rockville, MD, USA) platform. This analysis includes efficacy data through day 29, safety data through week 36, and PK data through week 24.

**Figure 1.**
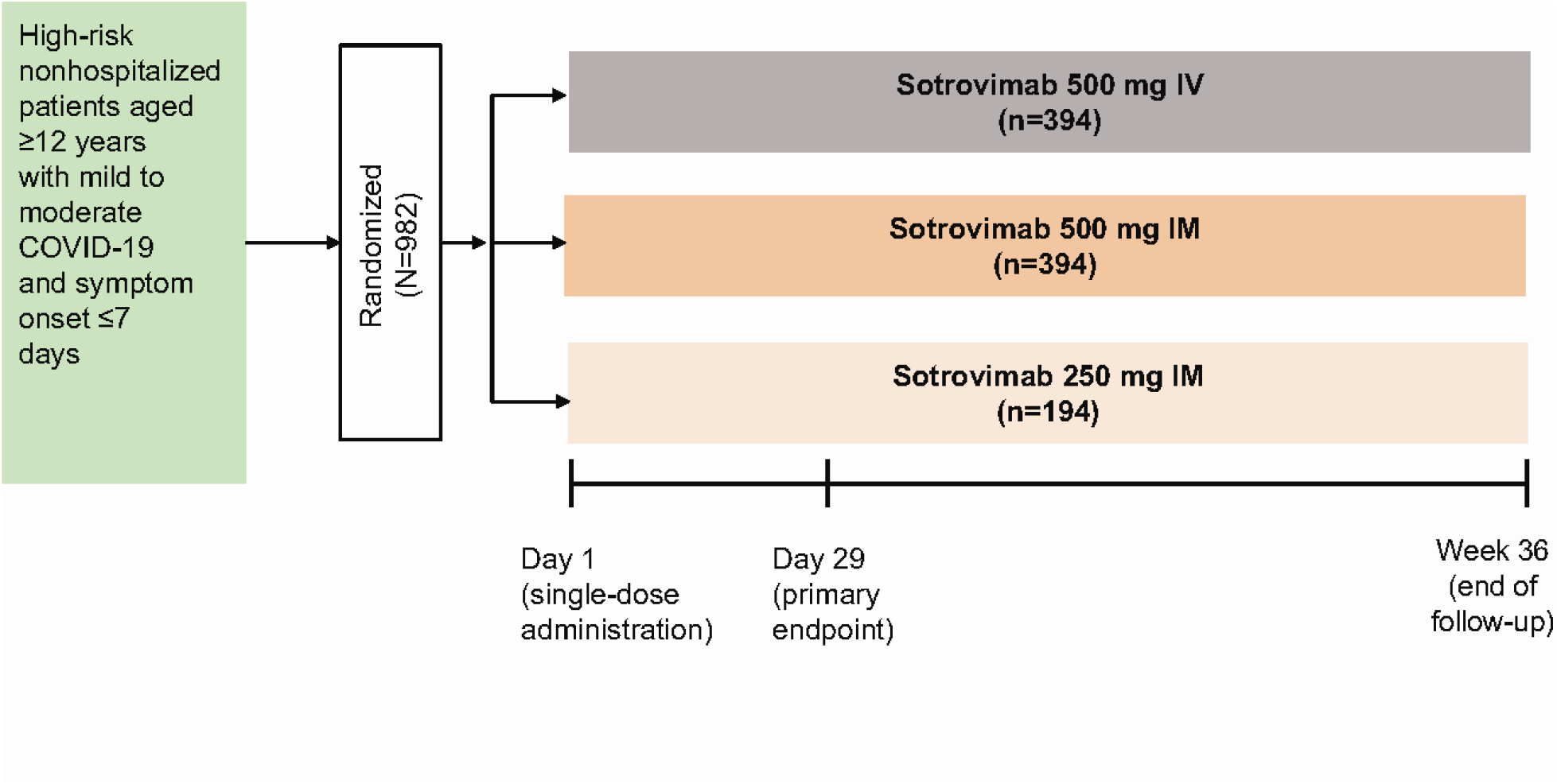
Study design.

### Outcomes

The primary endpoint was a composite of progression to hospitalization for >24 hours for acute management of any illness or death due to any cause through day 29. Secondary efficacy endpoints included SARS-CoV-2 viral load in nasal secretions measured by quantitative RT-PCR (qRT-PCR; key secondary endpoint 1); a composite of emergency department (ED) visit for management of any illness, hospitalization for acute management of any illness for any duration, or death due to any cause (key secondary endpoint 2); and development of severe and/or critical respiratory COVID-19 as manifested by requirement for respiratory support (including oxygen).

Adverse events (AEs), serious AEs, AEs of special interest, and disease-related events (DREs) were assessed through week 36. AEs of special interest included systemic and local infusion/injection-related reactions and local tolerability (injection-site reactions). Injection-site reactions in the IM groups were solicited at days 1, 3, 5, and 8 post-injection and reported separate from AEs. DREs were defined as AEs related to expected COVID-19 progression, signs, or symptoms, unless more severe than expected or if the investigator considered it related to study drug.

### Statistical analysis

A sample size of 340 participants per treatment group was expected to provide approximately 90% power to demonstrate that IM injection of sotrovimab was non-inferior to IV infusion of sotrovimab for the primary endpoint with a one-sided 2.5% type I error rate, assuming COVID-19 progression rates of 2% in the sotrovimab IM and IV groups and a 3.5% non-inferiority margin on the risk difference scale.

The intent-to-treat (ITT) population included all randomized participants, excluding those who were immunocompetent and fully vaccinated under the original protocol. The primary analysis population was based on the ITT population but excluded participants not meeting key eligibility criteria (e.g., those without a positive baseline SARS-CoV-2 test). Safety was assessed in all randomized participants exposed to study treatment (as-treated population).

The primary efficacy estimand used a hypothetical strategy to account for all intercurrent events (i.e., not receiving randomized treatment, discontinuation of study treatment, and use of medication not permitted during the study). Data observed after an intercurrent event were set to missing. Missing data were imputed under a missing at random assumption using multiple imputation. A post-hoc change was made to the multiple imputation algorithm from daily to weekly imputation due to the bias that was observed in the imputed progression rates from the daily imputation algorithm. Further details can be found in the appendix. A supplementary estimand was conducted in the efficacy population by handling all intercurrent events with a treatment policy strategy (i.e., regardless of the intercurrent events). A sensitivity analysis for the primary endpoint was conducted to determine the impact of imputing missing data as progressions.

The proportion of participants meeting the primary endpoint and key secondary endpoint 2 were compared between treatments using a binomial regression model with identity link function and adjusted for treatment group, age (<65, ≥65 years), and sex as covariates. The adjusted risk difference and associated 95% CI were computed to test non-inferiority of IM versus IV sotrovimab, which was declared if the upper bound of the two-sided 95% CI for the adjusted risk difference was <3.5%. Participants in the ITT population with a laboratory-confirmed quantifiable baseline nasopharyngeal swab at day 1 (virology population) were evaluated for mean area under the curve of SARS-CoV-2 viral load in nasal secretions as measured by qRT-PCR from day 1 to day 8 (AUC_d1-8_). Viral loads from IM and IV doses were compared for equivalence based on the two-sided 90% CI for the treatment ratio falling within equivalence bounds of 0.5 to 2.0.

A gate-keeping hierarchical testing procedure was used for testing the key secondary efficacy endpoints (Supplementary Figure 1). All statistical analyses were conducted using SAS, version 9.4 (SAS Institute Inc., Cary, NC, USA).

During the study, the safety review team noted a discrepancy in the rate of progression to hospitalization in the 250-mg IM group versus the 500-mg IM and 500-mg IV groups. An ad hoc interim data set was reviewed by an independent data monitoring committee and enrollment into the 250-mg IM group was subsequently discontinued. The study was changed to a two-group design with 1:1 randomization up to approximately 340 participants in each group (500 mg IM and 500 mg IV). The 250-mg IM group was removed from the testing hierarchy and data are summarized descriptively.

## Results

Between June 10 and August 19, 2021, 1039 participants were screened: 982 participants were randomly assigned to sotrovimab 500 mg IV (n=394), sotrovimab 500 mg IM (n=394), and sotrovimab 250 mg IM (n=194; **Figure 2**). Of the 982 participants, 29 were randomized under the original protocol and immunocompetent and fully vaccinated, thus excluded. An additional 16 participants were excluded: 14 who were immunocompetent and fully vaccinated and inadvertently enrolled under protocol amendment 2 and two participants without a positive SARS-CoV-2 test result. Thus, the primary analysis population consisted of 937 patients (500 mg IV, n=378; 500 mg IM, n=376; and 250 mg IM, n=183).

**Figure 2.**
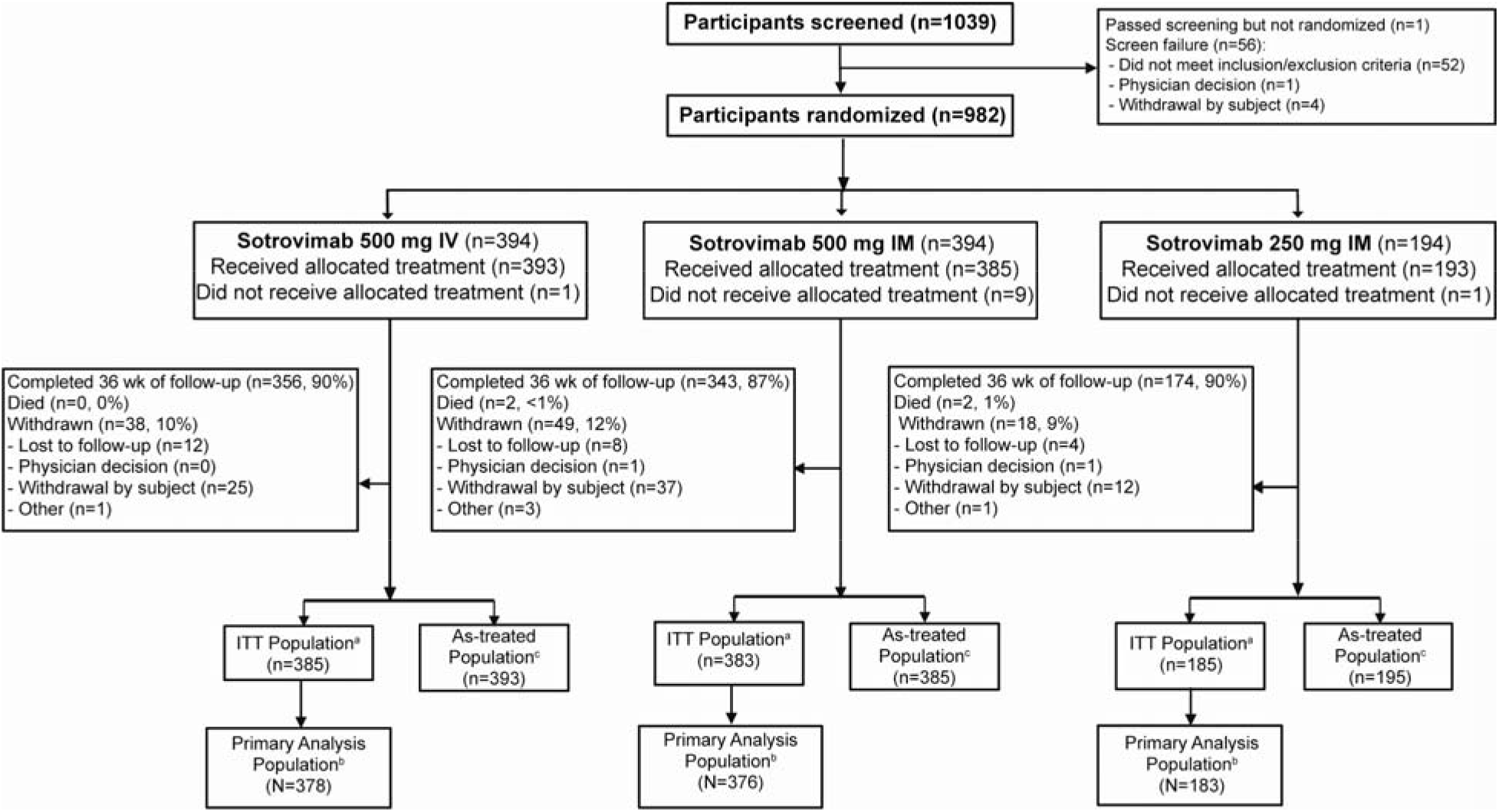
Patient enrollment and treatment assignment. ^a^ITT population includes all randomly assigned participants, excluding immunocompetent, fully vaccinated participants randomly assigned under the original protocol. ^b^Primary analysis population is defined as the ITT population minus those who had key inclusion/exclusion criteria violations. ^c^The as-treated (safety) population includes all participants who received study treatment. Two participants who were randomized to sotrovimab 500 mg IM received 250 mg IM and are included in the sotrovimab 250 mg IM as-treated population.

Participants were enrolled in Ukraine (<1%) and the United States (>99%), predominantly in Florida (Supplementary Table 1). Demographic and baseline characteristics were balanced across the sotrovimab groups, except for sex (**Table 1**). Approximately 23% were aged ≥65 years; most were Hispanic or Latino. The most common risk factors for COVID-19 progression were obesity, age ≥55 years, chronic lung disease, and diabetes; 3% of participants in each group had immunosuppressive disease. Nearly one-third of participants had ≥2 risk factors for COVID-19 progression. Most participants (86%–88%) had symptom duration of ≤5 days at baseline.

**Table 1.** Demographics and baseline characteristics (primary analysis population)

|  | **Sotrovimab 500 mg IV**<br>**(n=378)** | **Sotrovimab 500 mg IM**<br>**(n=376)** | **Sotrovimab 250 mg IM**<br>**(n=183)** |
| --- | --- | --- | --- |
| Sex |  |  |  |
| Female | 218 (58%) | 187 (50%) | 106 (58%) |
| Male | 160 (42%) | 189 (50%) | 77 (42%) |
| Age (years) | 51.0 (38.0-65.0) | 52.0 (37.5-64.5) | 48.0 (37.0-57.0) |
| Age group (years) |  |  |  |
| 12-17 | 2 (<1%) | 0 (0%) | 1 (<1%) |
| 18-64 | 281 (74%) | 282 (75%) | 156 (85%) |
| ≥65 | 95 (25%) | 94 (25%) | 26 (14%) |
| 65-74 | 65 (17%) | 56 (15%) | 17 (9%) |
| 75-84 | 24 (6%) | 32 (9%) | 8 (4%) |
| ≥85 | 6 (2%) | 6 (2%) | 1 (<1%) |
| Ethnicity |  |  |  |
| Hispanic or Latino | 312 (83%) | 320 (85%) | 157 (86%) |
| Not Hispanic or Latino | 66 (17%) | 56 (15%) | 26 (14%) |
| Race |  |  |  |
| Asian | 0 (0%) | 2 (<1%) | 0 (0%) |
| Black or African American | 14 (4%) | 17 (5%) | 8 (4%) |
| Mixed race | 1 (<1%) | 1 (<1%) | 1 (<1%) |
| Native Hawaiian or other Pacific Islander | 1 (<1%) | 0 (0%) | 0 (0%) |
| White | 360 (96%) | 352 (94%) | 172 (95%) |
| Other | 0 (0%) | 3 (<1%) | 1 (<1%) |
| BMI (kg/m<sup>2</sup>) | 30.97 (27.54-33.15) | 30.83 (27.75-32.70) | 31.23 (27.43-33.15) |
| Conditions as risk factor for COVID-19 progression |  |  |  |
| Any condition | 376 (>99%) | 374 (>99%) | 181 (99%) |
| Obesity<sup>a</sup> | 237 (63%) | 233 (62%) | 115 (63%) |
| Age ≥55 years | 158 (42%) | 163 (43%) | 62 (34%) |
| Chronic lung diseases | 59 (16%) | 69 (18%) | 43 (23%) |
| Diabetes | 48 (13%) | 46 (12%) | 22 (12%) |
| Immunosuppressive disease or immunosuppressive medications | 11 (3%) | 12 (3%) | 5 (3%) |
| Chronic liver disease | 4 (1%) | 4 (1%) | 4 (2%) |
| Congestive heart failure (NYHA class II or more) | 4 (1%) | 3 (<1%) | 1 (<1%) |
| Chronic kidney disease (eGFR <60 by MDRD equation) | 0 (0%) | 2 (<1%) | 0 (0%) |
| Congenital heart disease | 1 (<1%) | 0 (0%) | 1 (<1%) |
| Sickle cell disease | 1 (<1%) | 0 (0%) | 0 (0%) |
| Number of conditions met |  |  |  |
| 0 | 2 (<1%) | 2 (<1%) | 2 (1%) |
| 1 | 266 (70%) | 253 (67%) | 126 (69%) |
| 2 | 79 (21%) | 87 (23%) | 39 (21%) |
| 3 | 26 (7%) | 31 (8%) | 15 (8%) |
| >3 | 5 (1%) | 3 (<1%) | 1 (<1%) |
| COVID-19 vaccination history<sup>b</sup> |  |  |  |
| Yes | 17 (4%) | 18 (5%) | 11 (6%) |
| Symptom duration (days) |  |  |  |
| ≤5 | 324 (86%) | 332 (88%) | 160 (87%) |
| 5–7 | 54 (14%) | 44 (12%) | 23 (13%) |

Data are n (%) or median (IQR). BMI=body mass index. eGFR=estimated glomerular filtration rate. IQR=interquartile range.
MDRD=Modification of Diet in Renal Disease. NYHA=New York Heart Association. <sup>a</sup>Obesity defined as BMI ≥85th percentile for age/gender based on Centers for Disease Control and Prevention growth charts. <sup>b</sup>COVID-19 vaccination history was defined as receipt of at least one dose of any COVID-19 vaccination prior to randomization.

In the sotrovimab 500-mg IM group, 10/376 (2.7%) participants versus 5/378 (1.3%) participants in the sotrovimab 500-mg IV group met progression criteria for the primary endpoint (adjusted absolute risk difference: 1.06% [95% CI: −1.15% to 3.26%]; **Table 2**). The upper limit of the CI was lower than the prespecified non-inferiority margin of 3.5%, indicating that 500 mg IM is non-inferior to 500 mg IV for treatment of mild/moderate COVID-19. Among the five participants in the 500-mg IV group who were hospitalized >24 hours, two events were reported as COVID-19 related and three events were due to other causes (acute renal failure of donor kidney, appendicitis, and elevated glucose level). For the ten participants in the 500-mg IM group who were hospitalized >24 hours, six events were COVID-19 related and four were due to other causes (worsening bacterial pneumonia, acute appendicitis, shingles, and decompensated heart failure). Two participants in the 500-mg IM group who were hospitalized for COVID-19– related events died. Ten participants (5.5%) receiving 250 mg IM progressed, with nine events related to COVID-19 and one event due to other causes (exacerbation of chronic obstructive pulmonary disease). Two participants in the 250-mg IM group progressed to hospitalization for COVID-19–related events and died after day 29.

**Table 2.** Primary and secondary efficacy outcomes through day 29.

|  | Sotrovimab 500 mg IV | Sotrovimab 500 mg IM | Sotrovimab 250 mg IM |
| --- | --- | --- | --- |
| **Primary outcome (primary analysis population, hypothetical estimand)<sup>a</sup>** | n=378 | n=376 | n=183 |
| Hospitalized >24 hours or death, due to any cause | 5 (1.3%) | 10 (2.7%) | 10 (5.5%) |
| Hospitalized >24 hours due to any cause | 5 (1.3%) | 10 (2.7%) | 10 (5.5%) |
| Death due to any cause | 0 (0%) | 2 (0.5%) | 0 (0%) |
| Alive and not hospitalized | 364 (96.3%) | 351 (93.4%) | 170 (92.9%) |
| Missing | 9 (2.4%) | 15 (4.0%) | 3 (1.6%) |
| Absolute adjusted risk difference (95% CI): 500 mg IV vs 500 mg IM | 1.06% (−1.15% to 3.26%) |  |  |
| **Secondary outcomes** |  |  |  |
| 1. Mean AUC<sub>d1-8</sub> SARS-CoV-2 viral load (log10 copies/mL) (virology population) | n=287 | n=278 | n=136 |
| Geometric mean (% CV) | 25.42 (34.14%) | 25.56 (36.94%) | 25.46 (37.90%) |
| Adjusted LS geometric mean | 25.03 | 25.96 |  |
| Ratio (90% CI) | 1.04 (1.00 to 1.07) |  |  |
| 2. Hospitalization, ED visit, or death due to any cause (primary analysis population, hypothetical estimand)<sup>a</sup> | n=378 | n=376 | n=183 |
| Hospitalization, ED visit, or death due to any cause | 9 (2.4%) | 12 (3.2%) | 11 (6.0%) |
| Hospitalized | 5 (1.3%) | 10 (2.7%) | 10 (5.5%) |
| ED visit | 5 (1.3%) | 3 (0.8%) | 3 (1.6%) |
| Death | 0 (0%) | 2 (0.5%) | 0 (0%) |
| Alive and not hospitalized and no ED visit | 360 (95.2%) | 349 (92.8%) | 169 (92.3%) |
| Missing | 9 (2.4%) | 15 (4.0%) | 3 (1.6%) |
| Adjusted risk difference (95% CI): 500 mg IV vs 500 mg IM | 0.86% (−1.56% to 3.28%) |  |  |
| 3. Progression to severe/critical respiratory | n=385 | n=383 | n=185 |
| COVID-19 (intent-to-treat population) |  |  |  |
| Progression to severe/critical respiratory COVID-19<sup>b</sup> | 1 (0.3%) | 6 (1.6%) | 8 (4.3%) |
| Low-flow nasal cannula/face mask (severe) | 1 (0.3%) | 2 (0.5%) | 4 (2.2%) |
| Non-rebreather mask or high-flow nasal cannula/non-invasive ventilation (including continuous positive airway pressure support) | 0 (0%) | 2 (0.5%) | 2 (1.1%) |
| Mechanical ventilation/extracorporeal membrane oxygenation | 0 (0%) | 0 (0%) | 2 (1.1%) |
| Death | 0 (0%) | 2 (0.5%) | 0 (0%) |
| No progression to severe/critical respiratory COVID-19 | 375 (97.4%) | 362 (94.5%) | 174 (94.1%) |
| Missing | 9 (2.3%) | 15 (3.9%) | 3 (1.6%) |

Data are n (%) unless noted otherwise. CV=coefficient of variation. LS=least squares. qRT-PCR=quantitative reverse transcriptase-polymerase chain reaction. SARS-CoV-2=severe acute respiratory syndrome coronavirus 2. <sup>a</sup>Patients are counted in each subcategory of progression experienced up to the time point in question and so may be included in more than one category. <sup>b</sup>Severe respiratory COVID-19 was defined as a requirement for supplemental oxygen by either nasal cannula, face mask, high-flow oxygen devices, or non-invasive ventilation. Critical respiratory COVID-19 was defined as a requirement for invasive mechanical ventilation or extracorporeal membrane oxygenation. A patient's worst respiratory status over day 1 to day 29 is reported, with death as the maximal value.

For key secondary endpoint 1 of SARS-CoV-2 viral load in nasal secretions in the virology population (n=757), the adjusted mean viral load AUC_d1-8_ was equivalent between sotrovimab 500 mg IV (25.03 log10 copies/mL) and 500 mg IM (25.96 log10 copies/mL; 90% CI: 1.00 to 1.07), falling within the bounds of 0.5 to 2.0. The unadjusted mean AUC_d1-8_ for sotrovimab 250 mg IM was 25.46 log10 copies/mL (**Table 2**). Absolute viral load over time is shown in Supplementary Figure 2. Overall, there was no clear difference in the change in viral load over time across treatment arms.

For key secondary endpoint 2 of COVID-19 progression to ED visit, hospitalization for any duration, or death, nine (2.4%) participants in the 500-mg IV group and 12 (3.2%) in the 500-mg IM group met the progression criteria, with an adjusted risk difference of 0.86% and 95% CI: −1.56% to 3.28%; the upper limit of the CI was lower than the prespecified non-inferiority margin of 3.5% (**Table 2**). Progression to severe and/or critical respiratory COVID-19 occurred in one (0.3%) and six (1.6%) participants in the sotrovimab 500-mg IV and IM groups, respectively (**Table 2**). The participant who progressed after sotrovimab 500 mg IV required low-flow oxygen by nasal cannula or face mask. In the 500-mg IM sotrovimab group, two participants required low-flow oxygen and two participants required a non-rebreather mask or high-flow oxygen. Another two participants required supplemental oxygen via bilevel positive airway pressure and invasive mechanical ventilation; both participants subsequently died. Eleven participants (6.0%) receiving sotrovimab 250 mg IM met criteria for the key secondary endpoint 2 and eight participants (4.3%) progressed to severe and/or critical respiratory COVID-19 (**Table 2**).

Based on tipping point analyses, the outcome for the primary endpoint would switch from non-inferior to not non-inferior if the underlying progression rate in the missing data was >10.2% (or approximately 2/13 participants in the 500-mg IM group and 1/9 participants in the 500-mg IV group; Supplementary Figure 3). Results using the treatment policy estimand were consistent with those of the hypothetical estimand (Supplementary Table 2).

Overall, the incidence of AEs was low and similar between IV and IM treatment groups through week 36 (**Table 3**). Grade 3 or 4 AEs occurred in 2% of participants in each treatment group and none of these AEs were considered related to sotrovimab. Serious AEs were reported in three (<1%), seven (2%), and three (2%) participants in the 500-mg IV, 500-mg IM, and 250-mg IM sotrovimab groups, respectively. No serious AEs were considered related to treatment.

**Table 3.** Summary of AEs through week 36 (as-treated population^a^)

|  | **Sotrovimab 500**<br>**mg IV**<br>**(n=393)** | **Sotrovimab 500 mg**<br>**IM**<br>**(n=385)** | **Sotrovimab 250**<br>**mg IM**<br>**(n=195)** |
| --- | --- | --- | --- |
| Any AE | 39 (10%) | 41 (11%) | 26 (13%) |
| Related to study treatment<sup>b</sup> | 7 (2%) | 4 (1%) | 3 (2%) |
| Leading to permanent discontinuation<br>of study treatment | 0 (0%) | 0 (0%) | 0 (0%) |
| Leading to dose interruption/delay | 1 (<1%) | 0 (0%) | 0 (0%) |
| Any grade 3 or 4 AE | 8 (2%) | 8 (2%) | 4 (2%) |
| Related to study treatment<sup>b</sup> | 0 (0%) | 0 (0%) | 0 (0%) |
| Any serious AE | 3 (<1%) | 7 (2%) | 3 (2%) |
| Related to study treatment<sup>b</sup> | 0 (0%) | 0 (0%) | 0 (0%) |
| Fatal | 0 (0%) | 0 (0%) | 0 (0%) |
| Any injection/infusion-related reaction | 2 (<1%) | 1 (<1%) | 1 (<1%) |
| Chills | 1 (<1%) | 0 (0%) | 0 (0%) |
| Hypersensitivity | 1 (<1%) | 0 (0%) | 0 (0%) |
| Pyrexia | 1 (<1%) | 0 (0%) | 0 (0%) |
| Pruritus | 0 (0%) | 1 (<1%) | 0 (0%) |
| Asthma | 0 (0%) | 0 (0%) | 1 (<1%) |
| Any DRE | 18 (5%) | 16 (4%) | 20 (10%) |
| Leading to study discontinuation | 0 (0%) | 2 (<1%) | 2 (1%) |
| Any grade 3 or 4 DRE | 4 (1%) | 4 (1%) | 6 (3%) |
| Leading to study discontinuation | 0 (0%) | 0 (0%) | 0 (0%) |
| Any serious DRE | 3 (<1%) | 6 (2%) | 10 (5%) |
| Leading to study discontinuation | 0 (0%) | 2 (<1%) | 2 (1%) |
| Fatal | 0 (0%) | 2 (<1%) | 2 (1%) |
| DREs in ≥2 patients across treatment<br>groups |  |  |  |
| COVID-19 pneumonia | 2 (<1%) | 4 (1%) | 4 (2%) |
| Pneumonia | 1 (<1%) | 2 (<1%) | 5 (3%) |
| Increased lipase level | 1 (<1%) | 4 (1%) | 2 (1%) |
| Cough | 3 (<1%) | 0 (0%) | 2 (1%) |
| Acidosis | 2 (<1%) | 2 (<1%) | 0 (0%) |
| Dyspnea | 1 (<1%) | 2 (<1%) | 1 (<1%) |
| Pharyngeal erythema | 1 (<1%) | 2 (<1%) | 1 (<1%) |
| Bronchitis | 1 (<1%) | 0 (0%) | 2 (1%) |
| Back pain | 0 (0%) | 1 (<1%) | 1 (<1%) |
| COVID-19 | 0 (0%) | 2 (<1%) | 0 (0%) |
| Dehydration | 1 (<1%) | 0 (0%) | 1 (<1%) |
| Headache | 1 (<1%) | 1 (<1%) | 0 (0%) |
| Pyrexia | 0 (0%) | 0 (0%) | 2 (1%) |
| Thrombocytopenia | 1 (<1%) | 0 (0%) | 1 (<1%) |
| Thrombocytosis | 1 (<1%) | 1 (<1%) | 0 (0%) |

Data are n (%). <sup>a</sup>The as-treated population includes all patients who received study intervention and are analyzed according to the treatment received. <sup>b</sup>Relatedness was determined by individual study investigators.

Appendicitis was reported in one participant each in the 500-mg IV and IM groups. The other serious AEs, which occurred in one participant each, are listed in the Supplementary Data. AEs related to expected progression, signs, or symptoms of COVID-19 were reported separately as DREs. The most frequent DREs were COVID-19 pneumonia and pneumonia (**Table 3**). Two participants in the 500-mg IM group and two participants in the 250-mg IM group experienced serious DREs and died (Supplementary Data). No participants in the 500-mg IV group died.

Four participants had injection/infusion-related reactions (500 mg IV, n=2; 500 mg IM, n=1; 250 mg IM, n=1; **Table 3**). Solicited injection-site reactions following sotrovimab 500 mg IM and 250 mg IM were grade 1 in 39 (10%) and 22 (11%) participants, grade 2 in seven (2%) and two (1%) participants, and grade 3 in one (<1%) and zero participants, respectively. Grade 1 pain and tenderness at 15 to 30 minutes post-dose were the most common injection-site reactions. Few events occurred at day 3 and beyond.

In the 500-mg IV and IM groups, median serum concentrations of sotrovimab in participants with progression of COVID-19 were comparable to those who did not progress to hospitalization (Supplementary Figure 4 and Supplementary Table 3). However, in the 250-mg IM treatment group, median serum sotrovimab concentrations trended lower in participants who experienced progression (8.7 μg/mL on day 8) compared with participants without progression (14.0 μg/mL on day 8), suggesting interparticipant variability might explain clinical outcomes at this lower IM dose.

## Discussion

Despite the availability of mAb treatments for SARS-CoV-2, access to care can be limited due to logistical challenges owing to IV administration. An IM route of administration could improve patient access, allowing delivery in most clinical settings. In this study, 500 mg of sotrovimab administered by IM injection was non-inferior to the same dose administered by IV infusion for the early treatment of mild to moderate COVID-19 in high-risk, non-hospitalized participants. No clinically meaningful safety and tolerability issues were observed over 36 weeks, and data support post-dose monitoring for 30 minutes, which may further reduce healthcare burden. Infusion-related reactions were rare, and nearly all injection-site reactions were mild and resolved quickly. These findings support the use of 500 mg of sotrovimab given by IM injection for susceptible variants.

Notably, two participants died in the 500-mg IM group, versus no deaths in the 500-mg IV group. It is unknown whether the observed difference in deaths was due to chance or due to some other factor. Furthermore, the increased rate of progression in the 250-mg IM group compared with either of the 500-mg groups cannot be explained by differences in viral load, because the AUC_d1-8_ values were similar across the groups, consistent with findings that upper airway viral load is not an optimal biomarker for efficacy of SARS-CoV-2 treatments [1, 20, 21]. However, an exposure response analysis suggested that serum concentrations on day 5 and day 8 were significant predictors of response. Reductions in day 5 and day 8 serum levels were associated with higher model-predicted occurrence of progression [22].

Enrollment predominantly occurred in the state of Florida and coincided with a surge in the SARS-CoV-2 Delta variant in the southern US. Of 764 participants with sequencing results data available, 674 (88.2%) were infected with the Delta variant [23]. The enrolled population was mostly Hispanic/Latino, a population often underrepresented in clinical trials, but there was limited racial diversity in the trial. In addition, only 3% of patients were immunocompromised. Although there was no placebo group, the overall low absolute rate of hospitalization (<3%) in this study in both 500-mg groups supports continued efficacy of sotrovimab against the Delta variant [7]. Another potential limitation of the study is that the amount of missing data exceeds the number of primary outcome events, meaning the conclusion is sensitive to method of handling missing data. In addition, this trial did not include patient-related outcomes, such as symptom duration.

The global population continues building immunity and the proportion of infected patients progressing to severe disease is decreasing; however, current estimates indicate several thousand people continue to die every week due to COVID-19 globally (>6000-41000 per week in 2023) [24]. Recent data from robust observational clinical studies in England and France suggest comparable clinical effectiveness of sotrovimab in patients infected with Omicron BA.1, BA.2 and BA.5, despite a moderate reduction in in vitro neutralization activity of sotrovimab against BA.2, BA.5 and its sub-lineages [12-15, 25]. As the pandemic evolves to an endemic phase, it will continue to be important to understand which patients benefit most from treatments to prevent severe disease. Anti-SARS-CoV-2 mAbs such as sotrovimab continue to fulfill an unmet need by providing a safe, tolerable, single dose treatment option without drug-drug interactions.

The availability of a therapy that can be administered by the IM route will be especially critical for underserved populations and those without access to IV infusion centers. The knowledge gained from this study will support development of more feasible, non-IV (e.g. IM or subcutaneous) options of anti-SARS-CoV2 mAbs. Such options will be critical to expand access to underserved populations, decrease burden on the healthcare system, and protect vulnerable patient populations.

## Supporting information

COMET-TAIL Manuscript_Supp_v2

COMET-TAIL_Supp_FinalProtocolSAP

## Data Availability

The data collected for this study will not be made available to others. The study protocol and statistical analysis plan are included in the supplementary data.

## Funding

The study was supported by Vir Biotechnology, Inc. in collaboration with GSK.

## Acknowledgments

The authors thank Courtney St. Amour, PhD, of Lumanity Scientific Inc., for medical writing support, which was funded by Vir Biotechnology, Inc. and GSK, and Joseph Hogan, MS, Emma Gierman, MPH, and Katrina Wheeler, BS, of Vir Biotechnology, Inc., for clinical operations support.

## Author Contributions

PSP, DKH, EA, WWY, EM, JES, DA, SC, and AP conceptualized and designed the study. All authors acquired, analyzed and/or interpreted the data. DI and DC conducted the statistical analyses. AK, LAG, and DI accessed and verified the data. All authors drafted the manuscript and critically reviewed and revised the manuscript for important intellectual content. All authors had full access to all the data in the study, take responsibility for the accuracy of the analysis, and had authority over manuscript preparation and the decision to submit the manuscript for publication.

## Conflict of Interest Disclosures

AES, JA, AF, YG-R, RH, EJ, JM, and NP report acting as trial investigators for Vir Biotechnology and receiving non-financial support from Vir Biotechnology during the conduct of the study. AK reports acting as a trial investigator for Vir Biotechnology and receiving nonfinancial support from Vir Biotechnology during the conduct of the study; grants and consulting fees from Gilead Biosciences; clinical trial payments from Regeneron, Vir Biotechnology, GSK, and Gilead Biosciences; and serving on data safety monitoring or advisory boards for Gilead Biosciences. ES reports acting as a trial investigator for Vir Biotechnology and receiving nonfinancial support from Vir Biotechnology during the conduct of the study; research support from AbbVie, Eli Lilly, Otsuka, Eisai, and Ironshore; and serving on speaker bureaus for Janssen, Teva, and AbbVie. DC, MLA, SP, SC, EM, JES, WWY, EA, and LAG are employees of Vir Biotechnology and report stock ownership in Vir Biotechnology and third-party funding from GSK to Vir Biotechnology for the submitted work. PSP is an employee of Vir Biotechnology and reports stock ownership in Vir Biotechnology and third-party funding from GSK to Vir Biotechnology for the submitted work and reports patents pending for sotrovimab for the treatment of COVID-19. DKH was an employee of Vir Biotechnology and held stock in Vir Biotechnology at the time of the study; he is currently an employee of Janssen Pharmaceutical Companies of Johnson & Johnson and reports stock ownership in Johnson & Johnson. DI, AS, DA, AP, AN, NN, and QW are employees of GSK and report stock ownership in GSK.

## Notes

### Competing Interest Statement

Adrienne E. Shapiro, Jude Acloque, Almena Free, Yaneicy Gonzalez-Rojas, Rubaba Hussain, Erick Juarez, Jaynier Moya, and Naval Parikh report acting as trial investigators for Vir Biotechnology and receiving non-financial support from Vir Biotechnology during the conduct of the study. Anita Kohli reports acting as a trial investigator for Vir Biotechnology and receiving nonfinancial support from Vir Biotechnology during the conduct of the study; grants and consulting fees from Gilead Biosciences; clinical trial payments from Regeneron, Vir Biotechnology, GSK, and Gilead Biosciences; and serving on data safety monitoring or advisory boards for Gilead Biosciences. Elias Sarkis reports acting as a trial investigator for Vir Biotechnology and receiving nonfinancial support from Vir Biotechnology during the conduct of the study; research support from AbbVie, Eli Lilly, Otsuka, Eisai, and Ironshore; and serving on speaker bureaus for Janssen, Teva, and AbbVie. Deborah Cebrik, Maria L. Agostini, Sergio Parra, Sophia Chow, Erik Mogalian, Jennifer E. Sager, Wendy W. Yeh, Elizabeth L. Alexander, and Leah A. Gaffney are employees of Vir Biotechnology and report stock ownership in Vir Biotechnology and third-party funding from GSK to Vir Biotechnology for the submitted work. Phillip S. Pang is an employee of Vir Biotechnology and reports stock ownership in Vir Biotechnology and third-party funding from GSK to Vir Biotechnology for the submitted work and reports patents pending for sotrovimab for the treatment of COVID-19. David K. Hong was an employee of Vir Biotechnology and held stock in Vir Biotechnology at the time of the study; he is currently an employee of Janssen Pharmaceutical Companies of Johnson & Johnson and reports stock ownership in Johnson & Johnson. David Inman, Andrew Skingsley, Daren Austin, Amanda Peppercorn, Ahmed Nader, Nadia Noormohamed, and Qianwen Wang are employees of GSK and report stock ownership in GSK.

### Clinical Trial

NCT04913675

### Author Declarations

Ethics committee/IRB of Advarra Institutional Review Board, Columbia, Maryland, USA; CEQ at Medical Center of Limited Liability Company Harmoniya krasy, Kyiv, Ukraine; and CPP Sud-Est II - Groupement Hospitalier Est, Bron, France gave ethical approval for this work.

