## Supplementary material for "Intramuscular Versus Intravenous SARS-CoV-2 Neutralizing Antibody Sotrovimab for Treatment of COVID-19 (COMET-TAIL): A Randomized Non-inferiority Clinical Trial": COMET-TAIL Manuscript_Supp_v2

**Supplementary Data**

**Table of Contents**

| COMET-TAIL Investigators | 2 |
| --- | --- |
| eResults | 4 |
| Figure 1. Hierarchal testing procedure | 5 |
| Figure 2. Mean (SE) SARS-CoV-2 viral load in nasal secretions through day 29 (virology population) | 6 |
| Figure 3. Tipping point analysis of proportion of participants who had progression of COVID-19 (hospitalization >24 hours or death due to any cause) through day 29 using the hypothetical estimand | 7 |
| Figure 4. Boxplots of sotrovimab serum concentration by progression of COVID-19 (any hospitalization >24 hours or death due to any cause) through day 29 | 8 |
| Table 1. Enrollment by trial site location (randomly assigned participants) | 9 |
| Table 2. Primary and secondary efficacy outcomes through day 29 using the treatment policy estimand (intent-to-treat population) | 10 |
| Table 3. Sotrovimab serum concentration by progression of COVID-19 and treatment group through day 29 | 11 |

**COMET-TAIL Investigators**

Gerard Acloque ─ Universal Medical and Research Center, LLC, Florida, USA

Jude Acloque ─ BioClinical Research Alliance, Florida, USA

Ana Acosta ─ Advance Medical Research Center, Florida, USA

Hassan Ali ─ Allied Biomedical Research Institute, Florida, USA

Jorge Amaya ─ D&H National Research Centers, Florida, USA

Michael Bahrami ─ MedBio Trials, Florida, USA

Armando Curra ─ Miramax Clinical Research, Inc., Florida, USA

Jorge Diaz ─ Doral Medical Research, Florida, USA

Marta Dobryanska ─ Medical Center of Limited Liability Company “Harmoniya krasy”, Kyiv, Ukraine

Ankur Doshi ─ Primecare Medical Group, Texas, USA

Victor Escobar ─ 1960 Family Practice P.A. - Red Oak, Texas, USA

Ladynez Espinal ─ Admed Research-Miramar, Florida, USA

Alfredo Fernandez ─ Clinical Trials of Tampa, Florida, USA

Marta Fernandez ─ US Associates in Research Inc., Florida, USA

Almena Free ─ Pinnacle Research Group, Alabama, USA

Hiram Garcia ─ Rio Grande Valley Clinical Research Institute, Texas, USA

Marcy Goisse ─ Frontier Clinical Research, LLC, Pennsylvania, USA

Yaneicy Gonzalez-Rojas ─ Optimus U Corp, Florida, USA

Juan Gutierrez ─ Continental Clinical Research, LLC, Florida, USA

Luis Hernandez ─ Innovation Medical Research Center, Florida, USA

Reinaldo Hernandez-Loy ─ Dynamic Medical Research, LLC – Miami, Florida, USA

Rubaba Hussain ─ Prime Global Research, Inc., New York, USA

Erick Juarez ─ Florida International Medical Research, Florida, USA

Anita Kohli ─ The Institute for Liver Health-Tucson, Arizona, USA

John Kowalczyk ─ American Institute of Research, California, USA

Glen Leavitt ─ Leavitt Clinical Research, Idaho, USA

Luis Martinez ─ Universal Axon Clinical Research, Florida, USA

Eric Melvin ─ Clinical Trials of America-NC, LLC, North Carolina, USA

Shilpi Mittal ─ Care United Research, LLC, Texas, USA

Bharat Mocherla ─ Las Vegas Medical Research, Nevada, USA

Jaynier Moya ─ Pines Care Research Center Inc., Florida, USA

Naval Parikh ─ Napa Research, Florida, USA

Adrian Perez ─ Inpatient Research Clinic, LLC, Florida, USA

Karelia Ruiz ─ New Generation Medical Research, Florida, USA

Elvys Sacerio Valacarcel ─ Florida Healthcare System, Florida, USA

Yessica Sachdeva ─ Arizona Clinical Trials, Arizona, USA

Elias Sarkis ─ Sarkis Clinical Trials, Florida, USA

Gilberto Seco ─ Homestead Associates in Research Inc., Florida, USA

Adrienne Shapiro ─ Fred Hutch COVID-19 Clinical Research Center, Washington, USA

Lawrence Sher ─ Peninsula Research Associates - CRN – PPDS, California, USA

Guillermo Somodevilla ─ Cordova Research Institute, Florida, USA

Diego Torres ─ Ormond Beach Clinical Research, Florida, USA

Raidel Valdes ─ Crespo Model Research Center, LLC, Florida, USA

Luis Zepeda ─ Vilo Research Group, LLC, Texas, USA

**eResults**

*Serious adverse events (AEs)*

In addition to appendicitis, which was reported in one participant each in the 500-mg intravenous (IV) and intramuscular (IM) sotrovimab groups, the following other serious AEs occurred in one participant each: rejection of kidney transplant, increased blood glucose level, and acute kidney injury (each in the 500-mg IV sotrovimab group); herpes zoster, infective exacerbation of chronic obstructive airways disease, bacterial pneumonia, cerebrovascular accident, hemiparesis, cardiac failure, and hyponatremia (each in the 500-mg IM sotrovimab group); and chronic obstructive pulmonary disease, pulmonary embolism, intracranial mass, and seizure (each in the 250-mg IM sotrovimab group).

*Description of fatal events in the 500-mg IM group*

A man aged 41-45 years with a medical history notable for morbid obesity (body mass index [BMI]=69 kg/m^2^) and hypertension was randomly assigned to receive intramuscular (IM) sotrovimab 500 mg 5 days after symptom onset. He experienced shortness of breath and was admitted to the hospital with a serious disease-related event of coronavirus disease 2019 (COVID-19) pneumonia the same day as receiving sotrovimab. The patient was hospitalized in the intensive care unit (ICU), where he received supplemental oxygen via bilevel positive airway pressure and died on day 12; the primary cause of death was respiratory failure with a secondary cause of COVID-19.

A man aged 81-85 years with a medical history notable for obesity (BMI=31 kg/m^2^) and hypertension received sotrovimab 500 mg IM 4 days after symptom onset. On day 12, the patient experienced shortness of breath and went to the hospital, where he was found to have COVID-19 pneumonia with severe hypoxemia. He was admitted to the ICU, where he received invasive mechanical ventilation. On day 22, the patient had respiratory arrest and died. The cause of death was COVID-19 pneumonia.

*Description of fatal events in the 250-mg IM group*

A man aged 31-35 years with a medical history notable for obesity (BMI=34 kg/m^2^) received sotrovimab 250 mg IM 3 days after symptom onset. On day 5, he experienced a serious disease-related event of acute respiratory failure and was hospitalized in an inpatient/general ward, where he received supplemental oxygen via high-flow nasal cannula and was then switched to invasive mechanical ventilation. On day 46, the patient died due to acute respiratory failure.

A woman aged 66-70 years with a medical history notable for obesity (BMI=32 kg/m^2^) and hypertension received sotrovimab 250 mg IM 3 days after symptom onset. On day 2, she experienced shortness of breath and was taken to the hospital and admitted to the ICU with a serious disease-related event of severe pneumonia. On day 14, she received invasive mechanical ventilation and subsequently died on day 33 due to COVID-19.

**Supplementary Figure 1. Hierarchal testing procedure.** AUC_d1-8_=area under the curve of SARS-CoV-2 viral load in nasal secretions as measured by qRT-PCR from day 1 to day 8. COVID-19=coronavirus disease 2019. qRT-PCR= quantitative reverse transcriptase-polymerase chain reaction. SARS-CoV-2=severe acute respiratory syndrome coronavirus 2.

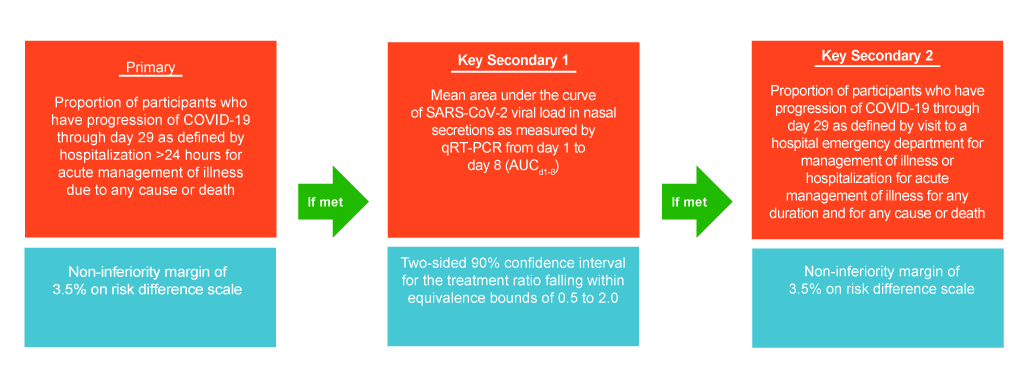

**Supplementary Figure 2. Mean (SE) SARS-CoV-2 viral load in nasal secretions through day 29 (virology population).** SARS-CoV-2 viral load was measured in nasal secretions by qRT-PCR from participants in the intent-to-treat population who had a laboratory-confirmed quantifiable baseline nasopharyngeal swab at day 1 (virology population). BL=baseline. qRT-PCR= quantitative reverse transcriptase-polymerase chain reaction. IM=intramuscular. IV-intravenous. SARS-CoV-2=severe acute respiratory syndrome coronavirus 2.

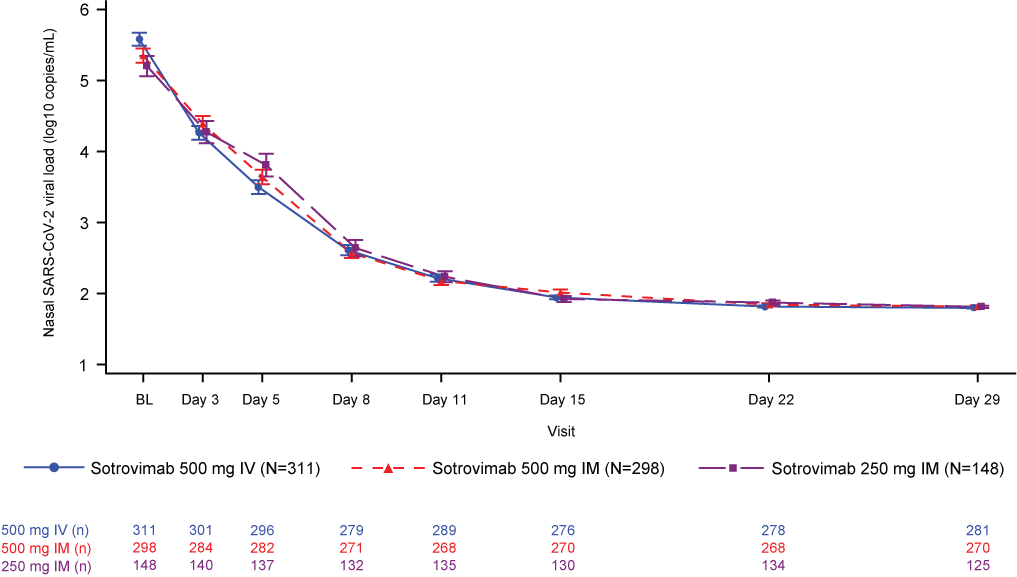

**Supplementary Figure 3. Tipping point analysis of proportion of participants who had progression of COVID-19 (hospitalization >24 hours or death due to any cause) through day 29 using the hypothetical estimand.** Analysis was performed using a binomial regression model with identity link function and with treatment (sotrovimab 500 mg IM, 500 mg IV), age (<65, ≥65 years), and sex (male, female) as covariates. Data occurring after occurrence of an intercurrent event was set to missing. Numbers in boxes represent the sotrovimab 500 mg IM vs 500 mg IV risk difference (%). Dashed lines represent the observed response rates on non-missing data. COVID-19=coronavirus disease 2019. IM=intramuscular. IV=intravenous.

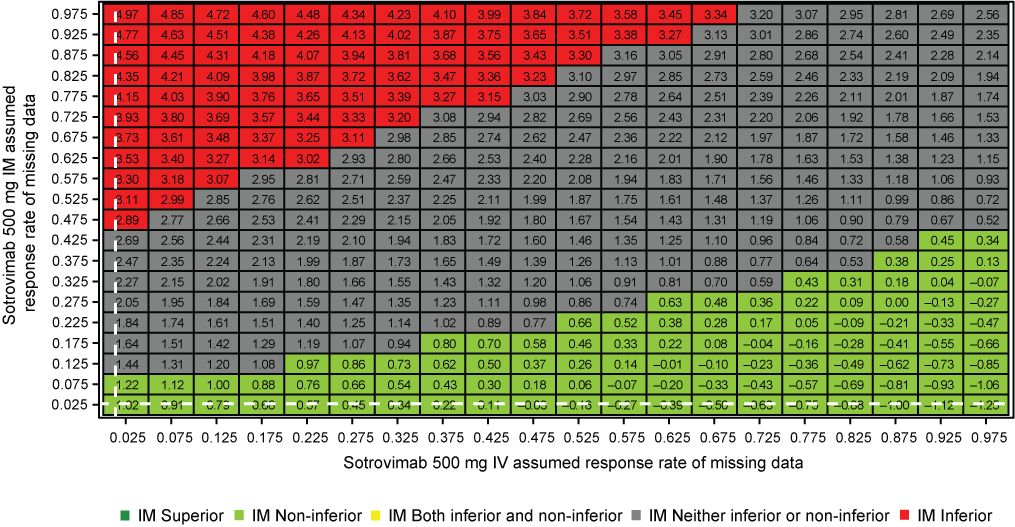

**Supplementary Figure 4. Boxplots of sotrovimab serum concentration by progression of COVID-19 (any hospitalization >24 hours or death due to any cause) through day 29.** COVID-19=coronavirus disease 2019. IM=intramuscular. IV=intravenous.

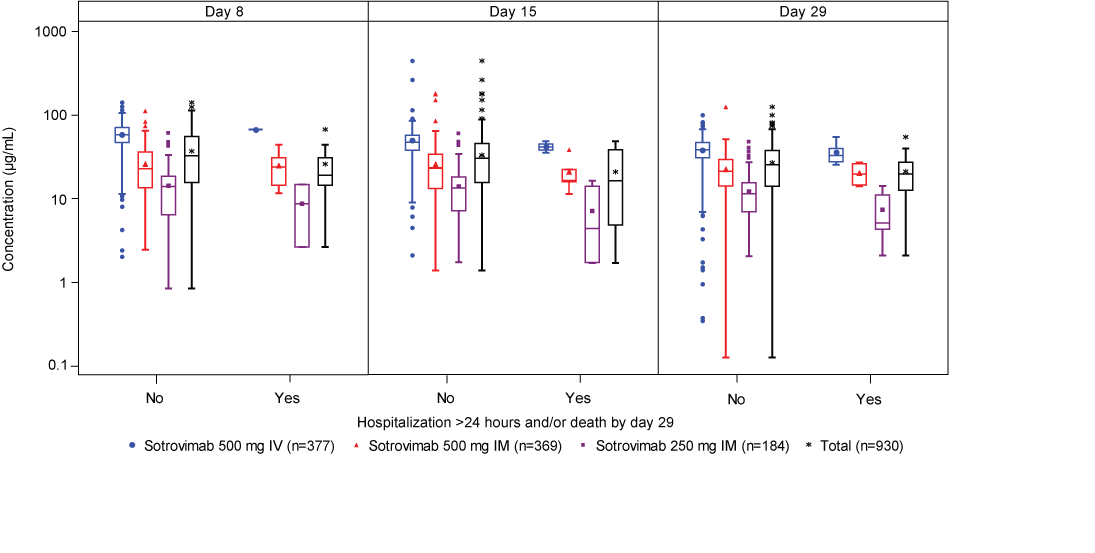

| **Location** | **Study sites** | **Patients enrolled** |
| --- | --- | --- |
| Ukraine |  |  |
| Kyiv | 1 | 2 |
| United States |  |  |
| Alabama | 1 | 15 |
| Arizona | 2 | 17 |
| California | 2 | 25 |
| Florida | 27 | 832 |
| Idaho | 1 | 1 |
| Nevada | 1 | 23 |
| New York | 1 | 11 |
| North Carolina | 1 | 2 |
| Pennsylvania | 1 | 1 |
| Texas | 5 | 36 |
| Washington | 1 | 17 |

Data are n.

**Supplementary Table 1. Enrollment by trial site location (randomly assigned participants)**

|  | **Sotrovimab 500 mg IV** | **Sotrovimab 500 mg IM** | **Sotrovimab 250 mg IM** |
| --- | --- | --- | --- |
| **Primary outcome^*^** | n=385 | n=383 | n=185 |
| Hospitalized >24 hours or death, due to any cause | 5 (1.3%) | 10 (2.6%) | 10 (5.4%) |
| Hospitalized >24 hours due to any cause | 5 (1.3%) | 10 (2.6%) | 10 (5.4%) |
| Death due to any cause | 0 (0%) | 2 (0.5%) | 0 (0%) |
| Alive and not hospitalized | 371 (96.4%) | 358 (93.5%) | 172 (93.0%) |
| Missing | 9 (2.3%) | 15 (3.9%) | 3 (1.6%) |
| Absolute adjusted risk difference (95% CI): 500 mg IV vs 500 mg IM | 1.04% (–1.12% to 3.20%) | |  |
| **Secondary outcome** |  |  |  |
| Hospitalization, ED visit, or death due to any cause^*^ | n=385 | n=383 | n=185 |
| Hospitalization, ED visit, or death due to any cause | 9 (2.3%) | 12 (3.1%) | 11 (5.9%) |
| Hospitalized | 5 (1.3%) | 10 (2.6%) | 10 (5.4%) |
| ED visit | 5 (1.3%) | 3 (0.8%) | 3 (1.6%) |
| Death | 0 (0%) | 2 (0.5%) | 0 (0%) |
| Alive and not hospitalized and no ED visit | 367 (95.3%) | 356 (93.0%) | 171 (92.4%) |
| Missing | 9 (2.3%) | 15 (3.9%) | 3 (1.6%) |
| Adjusted risk difference (95% CI): 500 mg IV vs 500 mg IM | 0.84% (–1.53% to 3.22%) | |  |

Data are n (%) unless noted otherwise. CI=confidence interval. ED=emergency department. IM=intramuscular. IV=intravenous. ^*^Patients are counted in each subcategory of progression experienced up to the time point in question and so may be included in >1 category.

**Supplementary Table 2. Primary and secondary efficacy outcomes through day 29 using the treatment policy estimand (intent-to-treat population)**

|  | **Sotrovimab 500 mg IV (n=377)** | | **Sotrovimab 500 mg IM (n=369)** | | **Sotrovimab 250 mg IM (n=184)** | |
| --- | --- | --- | --- | --- | --- | --- |
| **Sotrovimab serum concentration (μg/mL) by study visit** | **COVID-19 progression** | **No progression** | **COVID-19 progression** | **No progression** | **COVID-19 progression** | **No progression** |
| **Day 8** | (n=1) 67.3 | (n=319) 58.2 (2.0-141.0) | (n=6) 24.0 (11.6-44.3) | (n=317) 22.8 (2.5-112.0) | (n=2)  8.7 (2.6-14.8) | (n=153) 14.0 (0.8-60.8) |
| **Day 15** | (n=4)  41.4 (35.5-48.6) | (n=318) 47.4 (2.1-445.0) | (n=5)  16.5 (11.4-38.5) | (n=322) 23.4 (1.4-181.0) | (n=6)  4.4 (1.7-16.4) | (n=148) 13.5 (1.7-59.7) |
| **Day 29** | (n=5)  32.9 (25.5-54.6) | (n=338) 38.6 (0.3-99.5) | (n=6)  19.8 (14.1-27.0) | (n=333) 21.3 (0.1-125.0) | (n=5)  5.1 (2.1-14.2) | (n=159) 11.5 (2.1-47.6) |

Data are median (range). COVID-19=coronavirus disease 2019. IM=intramuscular. IV=intravenous. ^*^Defined as any hospitalization >24 hours or death due to any cause.

**Supplementary Table 3. Sotrovimab serum concentration by progression of COVID-19^*^ and treatment group through day 29 (hypothetical estimand)**
