## Supplementary material for "Intramuscular Versus Intravenous SARS-CoV-2 Neutralizing Antibody Sotrovimab for Treatment of COVID-19 (COMET-TAIL): A Randomized Non-inferiority Clinical Trial": COMET-TAIL_Supp_FinalProtocolSAP

### CLINICAL STUDY PROTOCOL

**Protocol Title:** A Phase 3 randomized, multi-center, open label study to assess the efficacy, safety, and tolerability of monoclonal antibody VIR-7831 (sotrovimab) given intramuscularly versus intravenously for the treatment of mild/moderate coronavirus disease 2019 (COVID-19) in high-risk non-hospitalized patients

**Protocol Number**: VIR-7831-5008 (GSK Study 217114)

**Compound Number or Name:** VIR-7831 (sotrovimab; GSK4182136)

Protocol Version & Date: Amendment #3, 04 October 2021

**Brief Title: Intramuscular VIR-7831 (sotrovimab) for mild/moderate COVID-19**

**Study Phase: Phase III**

**Acronym:** COMET-TAIL (**CO**VID-19 **M**onoclonal antibody **E**fficacy **T**rial– **T**reatment of **A**cute COVID-19 with **I**ntramuscular monoc**L**onal antibody)

**Sponsor Name and Legal Registered Address:**

This study is sponsored by Vir Biotechnology, Inc.
GlaxoSmithKline is supporting Vir Biotechnology, Inc. in the conduct of this study.

Vir Biotechnology, Inc.
499 Illinois Street, Suite 500
San Francisco, CA 94158
USA

**Regulatory Agency Identifying Number(s):**

**IND:** 149315
**EudraCT:** 2021-000623-13
**NCT:** NCT04913675

Copyright 2021 Vir Biotechnology, Inc. and the GlaxoSmithKline group of companies. All rights reserved. Unauthorized copying or use of this information is prohibited.

### INVESTIGATOR SIGNATURE PAGE

VIR BIOTECHNOLOGY, INC

499 ILLINOIS STREET SUITE 500

SAN FRANCISCO, CA 94158

STUDY ACKNOWLEDGMENT

A Phase 3 randomized, multi-center, open label study to assess the efficacy, safety, and tolerability of monoclonal antibody VIR-7831 (sotrovimab) given intramuscularly versus intravenously for the treatment of mild/moderate coronavirus disease 2019 (COVID-19) in high-risk non-hospitalized patients, Protocol Amendment 3, 04 October 2021

This protocol has been approved by Vir Biotechnology, Inc. The following signature documents this approval.

| | |
|---|---|
| PPD, PhD<br>Vir Study Director | *{See Appended Electronic Signature Page}* |
| **Printed Name** | **Signature and Date** |

#### INVESTIGATOR STATEMENT

I have read the protocol, including all appendices, and I agree that it contains all necessary details for me and my staff to conduct this study as described. I will conduct this study as outlined herein and will make a reasonable effort to complete the study within the time designated.

I will provide all study personnel under my supervision copies of the protocol and access to all information provided by Vir Biotechnology, Inc. I will discuss this material with them to ensure they are fully informed about the drugs and the study.

| | |
|---|---|
| **Principal Investigator Printed Name** | **Signature** |
| | **Date** |

### LIST OF FIGURES

Figure 1: Mean (+ standard deviation [SD]) Sotrovimab Serum Concentration-Time Plots (Linear and Semi-log): COMET-ICE Lead-In Phase ........ 19

### 1. PROTOCOL SUMMARY

#### 1.1. Synopsis

**Title:** A Phase 3 randomized, multi-center, open label study to assess the efficacy, safety, and tolerability of monoclonal antibody VIR-7831 (sotrovimab) given intramuscularly versus intravenously for the treatment of mild/moderate coronavirus disease 2019 (COVID-19) in high-risk non-hospitalized patients.

**Rationale:**

There is an urgent medical need for therapeutics for the treatment of Severe Acute Respiratory Syndrome coronavirus-2 (SARS-CoV-2) infection, the cause of coronavirus disease 2019 (COVID-19). Early treatment of mild and moderate disease in outpatients could prevent the more severe sequelae of COVID-19 requiring hospitalization, such as respiratory failure, thromboembolic disease leading to pulmonary embolism and stroke, arrhythmias, and shock. Furthermore, a potent treatment given early in disease could ameliorate the severity and duration of COVID-19 and potentially reduce transmission. The ability to administer such a treatment easily in community settings is desirable.

Vir Biotechnology, Inc. (Vir) has developed a fully human neutralizing anti-SARS-CoV-2 antibody (Pinto 2020), sotrovimab (also known as VIR-7831; GSK4182136), which has an Fc-modification ("LS") that is designed to improve bioavailability in the respiratory mucosa (Hope 2019) and increase half-life (Ko 2014). Furthermore, Vir has formulated this monoclonal antibody for both intravenous (IV) and intramuscular (IM) administration.

As of June 2021, there are three anti-SARS-CoV-2 monoclonal antibody treatments that are authorized under Emergency Use Authorization (EUA) from the FDA for the early treatment of COVID-19: bamlanivimab/etesevimab (Eli Lilly 2021b), casirivimab/imdevimab (Regeneron 2021), and sotrovimab (FDA 2021). These mAb treatments are administered intravenously which has limited their availability as hospitals and clinics generally provide infusions in dedicated facilities usually reserved for chemotherapy or other biologics (Goldstein 2020). There is a need to develop a more convenient method of administration so patients can have easier access to anti-SARS-CoV-2 antibody treatments. Sotrovimab, administered intravenously, is currently being evaluated in a Phase 3 randomized, multi-center study to assess the safety and efficacy for the early treatment of COVID-19 in non-hospitalized patients (VIR-7831-5001; GSK study 214367; COMET-ICE). The COMET-ICE study uses the first generation formulation of sotrovimab (Gen1) which can only be given intravenously. The second generation formulation (Gen2) of sotrovimab can be administered intravenously or intramuscularly. On 10 Mar 2021, the independent data monitoring committee of COMET-ICE recommended that the study be stopped for enrollment due to evidence of clinical efficacy. This was based on an interim analysis of data from 583 patients which demonstrated an 85% (p=0.002) reduction in hospitalization or death in patients receiving sotrovimab compared to placebo (Vir 2021).

This current study aims to assess the efficacy, safety, and tolerability of intramuscular (IM) sotrovimab versus intravenous (IV) sotrovimab when given to high-risk patients for the treatment of mild/moderate COVID-19. This study will evaluate the clinical efficacy of two dose levels of IM sotrovimab compared with IV sotrovimab in preventing progression of COVID-19 by utilizing a non-inferiority (NI) design.

Gen2 sotrovimab material is also currently being studied to evaluate safety, tolerability, and pharmacokinetics in patients with mild to moderate COVID-19 [GSK216912 (VIR-7831-5006), also known as COMET-PEAK]. In COMET-PEAK, Part A is comparing the safety and pharmacokinetics of Gen1 and Gen2 sotrovimab when administered intravenously. Part B of COMET-PEAK will also compare the virologic activity of intravenous versus intramuscular administration of sotrovimab in patients with mild to moderate COVID-19 at low-risk of disease progression. Intramuscular injection of sotrovimab will also be evaluated in a planned study of prophylaxis against COVID-19 [GSK214368 (VIR-7831-5003), also known as COMET-STAR].

**Objectives and Endpoints:**

| Objectives | Endpoints |
|---|---|
| **Primary** | |
| Evaluate the efficacy of two dose levels of intramuscular sotrovimab versus intravenous sotrovimab in preventing the progression of mild/moderate COVID-19 | Progression of COVID-19 through Day 29 as defined by:<br>1. Hospitalization > 24 hours for acute management of illness due to any cause<br>OR<br>2. Death |
| **Secondary** | |
| **Safety**<br>Describe the safety and tolerability of intramuscular and intravenous sotrovimab | • Occurrence of adverse events (AEs)<br>• Occurrence of serious adverse events (SAEs)<br>• Occurrence of adverse events of special interest (AESI)<br>• Occurrence of disease related events (DRE) |
| Assess the immunogenicity of sotrovimab | Incidence and titers (if applicable) of serum anti-drug antibody (ADA) to sotrovimab |
| **Efficacy**<br>Evaluate the efficacy of two dose levels of intramuscular versus intravenous sotrovimab on the progression of mild/moderate COVID-19 | Progression of COVID-19 through Day 29 as defined by:<br>1. Visit to a hospital emergency room for management of illness<br>OR<br>2. Hospitalization for acute management of illness for any duration and for any cause<br>OR<br>3. Death |

| Objectives | Endpoints |
|---|---|
| Evaluate the efficacy of two dose levels of intramuscular versus intravenous sotrovimab in preventing COVID-19 respiratory disease progression | Development of severe and/or critical respiratory COVID-19 as manifested by requirement for respiratory support (including oxygen) at Day 8, Day 15, Day 22, and Day 29 |
| Compare the virologic activity of sotrovimab given IM (two dose levels) or IV in reducing SARS-CoV-2 viral load | • Mean area under the curve of SARS-CoV-2 viral load in nasal secretions as measured by qRT-PCR from Day 1 to Day 8 ($AUC_{D1-8}$)<br>• Change from baseline in viral load by qRT-PCR at Day 8<br>• Proportion of participants with a persistently high SARS-CoV-2 viral load at Day 8 by qRT-PCR |
| **Pharmacokinetics**<br>Assess the pharmacokinetics (PK) of sotrovimab in serum following IV and IM (two dose levels) administration | IV and IM sotrovimab pharmacokinetics (PK) in serum |
| **Exploratory** | |
| Describe the effect of two doses levels of intramuscular versus intravenous sotrovimab on incidence and duration of time on total hospital length of stay (LOS), incidence and duration of time on a ventilator, and ICU length of stay | • Incidence of hospitalization through Day 29<br>• Total hospital LOS<br>• Proportion of participants requiring ICU stay or mechanical ventilation through Day 29<br>• Total ICU LOS |
| Monitor SARS-CoV-2 resistant mutants against sotrovimab | • SARS-CoV-2 resistance mutants to sotrovimab at baseline<br>• Emergence of viral resistance mutants to mAb by SARS-CoV-2 |
| Compare the virologic activity of sotrovimab given IM (two dose levels) or IV in reducing SARS-CoV-2 viral load | • Change from baseline in viral load in nasal secretions by qRT-PCR during follow-up period at Day 5, Day 11, Day 15, Day 22 and Day 29<br>• Undetectable SARS-CoV-2 in nasal secretions by qRT-PCR at Day 3, Day 5, Day 8, Day 11, Day 15, Day 22 and Day 29<br>• Mean area under the curve of SARS-CoV-2 viral load as measured by qRT-PCR from Day 1 to Day 5 ($AUC_{D1-5}$) and Day 1 to 11 ($AUC_{D1-11}$) |

| Objectives | Endpoints |
|---|---|
| Compare the effect of different sample collection methods in SARS-CoV-2 viral load (e.g. nasopharyngeal swab, saliva) | • SARS-CoV-2 viral load measured by qRT-PCR |
| Evaluate the effect of sotrovimab on the development of SARS-CoV-2 antibodies | • SARS-CoV-2 anti-N antibody at Day 1 and Day 29 |
| Describe the effect of sotrovimab on Long COVID symptoms | • Incidence and severity of Long COVID symptoms at W12, W24 and W36 |

**Overall Design:**

This study is a Phase 3 randomized, multi-center, open label, non-inferiority study of intramuscular (IM) versus intravenous (IV) administration of sotrovimab, a monoclonal antibody (mAb) against SARS-CoV-2 for the treatment of mild/moderate COVID-19 in participants aged 12 years old and older at high risk of disease progression. The study will randomize 1:1:1 to receive a single dose IV infusion of sotrovimab or a single IM injection of sotrovimab at one of two dose levels.

Participants with early, mild/moderate COVID-19 who are not on supplemental oxygen and at risk for disease progression will receive an IV infusion or IM injection of sotrovimab. The IV sotrovimab dose will be 500 mg and will be infused over 15 minutes. The IM sotrovimab dose will be either 250 mg or 500 mg given at a single timepoint. The 250 mg IM dose will be given either as a single 250 mg (4ml) injection in the dorsogluteal muscle or as two 2 ml injections in each deltoid muscle. The 500 mg IM dose will be administered as two 4 ml injections in each dorsogluteal muscle.

After IV infusion or IM injection, participants will be monitored for 30 minutes with vital signs assessments performed every 15 minutes. Injection site reaction assessments, for IM arms only, will be performed at 15 and 30 minutes after injection. All participants will be actively monitored on an outpatient basis with frequent collection of nasopharyngeal swabs for virology and blood draws for PK sampling, as well as weekly in-clinic evaluations at Weeks 1, 2, 3, and 4 as detailed in the Schedule of Activities.

Starting at Week 8, participants will be monitored monthly via phone call or in-clinic evaluation to assess for the incidence and severity of subsequent COVID-19 illness, if any, for a total of 36 weeks from dosing.

**Number of Participants:**

Approximately 1020 participants were to be enrolled and randomized 1:1:1 to receive a single 500 mg IV infusion of sotrovimab, a single 500 mg IM injection of sotrovimab, or a single 250 mg IM injection of sotrovimab. An interim analysis (IA) was originally planned for an independent data monitoring committee (IDMC) to evaluate each dose level of IM sotrovimab versus IV sotrovimab for safety, futility and efficacy when approximately 50% of participants enrolled and reached Day 29. During the conduct of the study, the joint safety review team (JSRT) noted a discrepancy in the rate of progression to hospitalization occurring in the 250 mg IM arm compared with the 500 mg

IM and IV arms, and an ad hoc interim dataset was escalated to the IDMC for review. As a result of this review, enrollment into the 250 mg IM arm was discontinued and the study design was changed to a 2-arm study with 1:1 randomization up to ~340 participants/arm into the remaining 2 arms of 500 mg IM and 500 mg IV.

**Intervention Groups and Duration:**

Screening assessments will be performed within 48 hours prior to randomization. Screening can occur on the same day as randomization. Eligible participants will be treated with a single IM or IV dose of sotrovimab on Day 1 and followed for up to 36 weeks.

Participants will be stratified based on:

1. Age: 12-17 years old, 18-64 years old, and ≥65 years old
2. COVID-19 Vaccination History: receipt of any COVID-19 vaccine
3. Region of the world due to potential differences in standard of care for COVID-19 management and different circulating SARS-CoV-2 variants.

**Saliva SARS-CoV-2 qRT-PCR substudy**

To evaluate the comparability of SARS-CoV-2 quantitative RT-PCR when performed on saliva versus nasopharyngeal (NP) swabs, an optional sub-study may be performed. Up to 200 subjects will have paired saliva and NP collected at 4 timepoints during the study (D1, D8, D15, D22).

#### 1.2. Study Schema

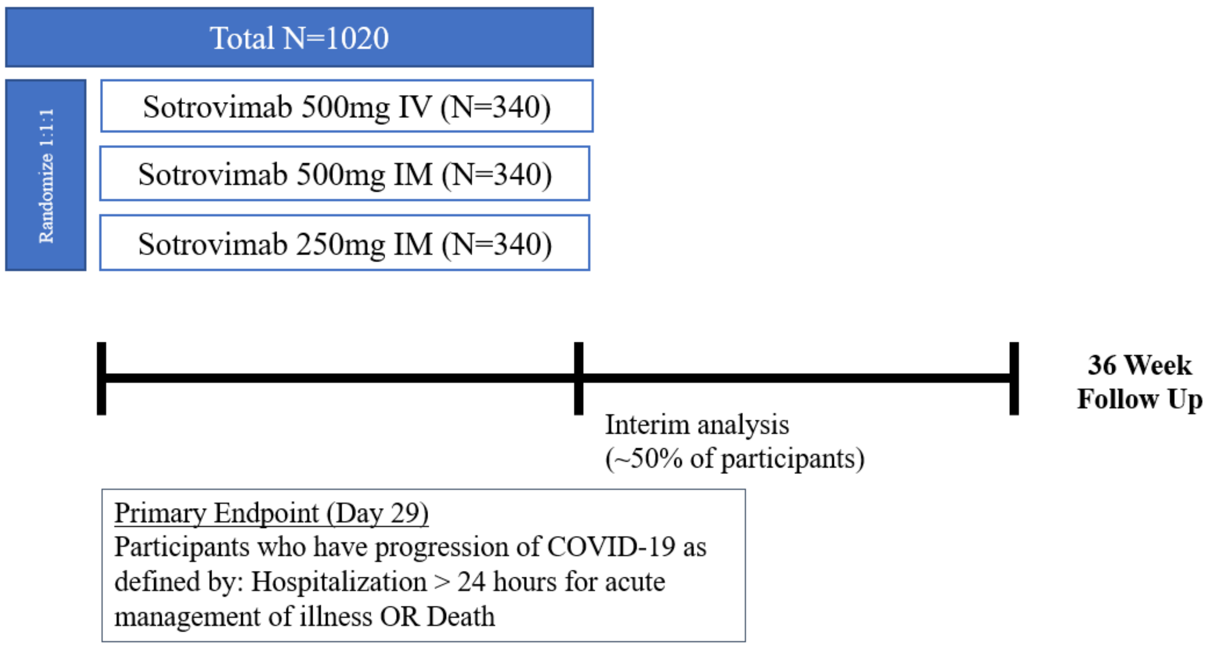

#### 1.3. Schedule of Activities

##### Table 1: Schedule of Activities

| Study Stage | Screening | Dosing | Follow-up Period | | | | | | | | | | | | | | |
|---|---|---|---|---|---|---|---|---|---|---|---|---|---|---|---|---|---|
| Study Visit Week | | | W1 | | | W2 | | W3 | W4 | W8 [j] | W12 | W16 [j] | W20 | W24 (EW[k]) | W28 [j] | W32 [j] | W36 [j] (EOS/EW[k]) |
| Study Visit Day ± Visit Window | D -2 to -1 | D1 | D3 | D5±1 | D8±1 | D11±1 | D15±1 | D22±1 | D29±2 | D57±4 | D85±7 | D113±7 | D141±7 | D169±7 | D197±7 | D224±7 | D252±7 |
| Informed consent | X | | | | | | | | | | | | | | | | |
| Demography | X | | | | | | | | | | | | | | | | |
| Medical history | X | | | | | | | | | | | | | | | | |
| Inclusion/exclusion criteria including baseline COVID-19 symptoms | X | | | | | | | | | | | | | | | | |
| Physical examination[a] | X[a] | X[a] | | | X[a] | | X[a] | X[a] | X[a] | | | | | | | | |
| Local tolerability assessment | | X[b] | X | X | X | | | | | | | | | | | | |
| Body weight & height | X | | | | | | | | | | | | | | | | |
| Vital signs (including O2 saturation) | X | X[c] | | | X | | X | X | X | | | | | X | | | |
| Local safety lab assessments | X[d] | | | | | | | | | | | | | | | | |
| Laboratory assessments | | X[e] | | | X | | X | | X | | | | | | | | |
| SARS-CoV-2 diagnostic test [i] (point-of-care or local laboratory test) | X[i] | | | | | | | | | | | | | | | | |
| Pregnancy test | X | | | | | | | | | | | | | X | | | |
| Electrocardiogram | X[f] | | | | | | | | | | | | | | | | |
| Randomization | | X | | | | | | | | | | | | | | | |
| Study drug administration | | X | | | | | | | | | | | | | | | |
| Nasopharyngeal swab for virology | | X[e] | X | X | X | X | X | X | X | | | | | | | | |
| Saliva sample for virology (optional substudy) | | X[e] | | | X | | X | X | | | | | | | | | |
| Blood sample for PK analysis | | X[g] | | | X | | X | | X | | X | | X | X | | | |

| **Study Stage** | **Screening** | **Dosing** | **Follow-up Period** | | | | | | | | | | | | | | |
|---|---|---|---|---|---|---|---|---|---|---|---|---|---|---|---|---|---|
| **Study Visit Week** | | | **W1** | | | **W2** | | **W3** | **W4** | **W8** [j] | **W12** | **W16** [j] | **W20** | **W24 (EW[k])** | **W28** [j] | **W32** [j] | **W36** [j] **(EOS/EW[k])** |
| **Study Visit Day ± Visit Window** | **D -2 to -1** | **D1** | **D3** | **D5±1** | **D8±1** | **D11±1** | **D15±1** | **D22±1** | **D29±2** | **D57±4** | **D85±7** | **D113±7** | **D141±7** | **D169±7** | **D197±7** | **D224±7** | **D252±7** |
| Blood sample for anti-drug antibody | | X[e] | | | | | | | X | | X | | X | X[k] | | | |
| Blood sample for anti-N SARS-CoV-2 antibody | | X[e] | | | | | | | X | | | | | | | | |
| Long COVID assessment | | | | | | | | | | | X | | | X | | | X |
| Phone call or clinic visit for subsequent COVID19 illness | | | | | | | | | | X | X | X | X | X | X | X | X |
| Review/record AE/SAE | | X[h] | | | | | | | | | | | | | | | |
| Concomitant medications | | X | | | | | | | | | | | | | | | |

[a] Complete physical examination should be performed on Day 1; all other visits should be symptom-directed physical examinations.
[b] Local tolerability assessments should be performed at 15 mins (± 5 mins) and 30 minutes (± 5 mins) after injection (only for participants who received IM injection).
[c] Vital signs on Day 1 should be recorded prior to dose administration and at 15 mins (± 5 mins) and 30 mins (± 5 mins) after injection or infusion.
[d] All local lab assessments may be performed or not as determined necessary by the investigator or required by local regulations.
[e] On Day 1, sample collection will occur pre-dose.
[f] Electrocardiogram at screening only for participants with a history of cardiovascular disease or diabetes
[g] On Day 1, PK sample will be collected pre-dose.
[h] Adverse events will be assessed up to Week 12 post dose. Serious adverse events (SAEs) will be assessed up to Week 36.
[i] Participants may be tested at screening if symptom onset ≤ 7 days, but dosing (D1 visit) must also occur ≤ 7 days of symptom onset.
[j] Week 8,Week 16, Week 28, Week 32 and Week 36 are planned as site phone calls to participants only.
[k] If subject withdraws early from the study prior to Week 24, then the W24 visit assessments will be performed as the EW visit. If subject withdraws early from the study after Week 24, the W36 visit assessments will be performed as the EW visit.

### 2. INTRODUCTION

#### 2.1. Study Rationale

This study is a Phase 3 randomized, multi-center, open label study of intramuscular (IM) versus intravenous (IV) administration of sotrovimab, a monoclonal antibody (mAb) against SARS-CoV-2 for the prevention of progression of mild/moderate COVID19 in participants 12 years old and older at high risk of disease progression. High-risk adults include participants ≥55 years old regardless of co-morbidities and individuals age 18 or older with comorbidities associated with worse outcomes in adults, including diabetes, obesity (BMI≥30), chronic kidney disease, congestive heart failure, chronic lung diseases (i.e. chronic obstructive pulmonary disease, moderate to severe asthma requiring steroids, interstitial lung disease, cystic fibrosis, and pulmonary hypertension), immunosuppressive disease or immunosuppressive medications, or chronic liver disease (CDC 2021). Adolescents, aged 12-17 years of age, at high-risk for COVID-19 progression include those with obesity (BMI ≥85th percentile for age/gender), chronic kidney disease, sickle-cell disease, congenital heart disease, neurodevelopmental disorders, medically-related technology dependence such as tracheostomy, ventilator-support, or a gastrostomy tube, or chronic lung diseases (i.e. chronic obstructive pulmonary disease, moderate to severe asthma requiring steroids, interstitial lung disease, cystic fibrosis, and pulmonary hypertension), immunosuppressive disease or immunosuppressive medications or chronic liver disease. (CDC 2021; Shekerdemian 2021) The study will include outpatients infected with SARS-CoV-2, the virus that causes COVID-19, as confirmed by local laboratory tests and/or point of care tests.

Enrollment in the study will be performed at study site, based on hospital local practice. If patient agrees to participate and qualifies, his/her primary doctor will be informed as per local regulation of his/her study participation by the study doctor.

Sotrovimab is currently being studied in a Phase 3 placebo-controlled trial evaluating the efficacy of sotrovimab in preventing progression of COVID-19 in high-risk patients with mild/moderate COVID-19 (VIR-7831-50001; COMET-ICE). On 10 Mar 2021, the independent data monitoring committee for COMET-ICE recommended that the study be stopped for enrollment due to evidence of clinical efficacy. This was based on an interim analysis of data from 583 patients which demonstrated an 85% (p=0.002) reduction in hospitalization or death in patients receiving sotrovimab compared to placebo. Sotrovimab was well-tolerated with no safety signals detected. (Vir 2021) In addition, recently both the NIH COVID-19 Treatment Guidelines and the Infectious Diseases Society of America have both recommended monoclonal antibody therapy treatment for high-risk individuals with mild/moderate COVID-19. (NIH 2021)

In light of positive data from COMET-ICE demonstrating overwhelming clinical efficacy of sotrovimab in preventing COVID-19 disease progression and the changes in treatment guidelines noted above, enrollment to placebo is not ethically favorable for participants who are at risk of disease progression as it would unnecessarily subject participants to potential morbidity and mortality despite the availability of an otherwise favorable intervention.

Give the urgent unmet medical need and the ethical considerations of a placebo-controlled design in the setting of already demonstrated clinical efficacy, the design for this current trial is a non-inferiority study comparing the clinical efficacy of two dose levels of IM sotrovimab versus IV sotrovimab using a clinical endpoint of progression of COVID-19 as defined by hospitalisation >24 hours or death by Day 29. The primary endpoint to assess efficacy of treatment is progression of mild/moderate COVID-19 defined by hospitalization >24 hours for acute management of illness or death due to any cause within 29 days of randomization. The transition from outpatient status without hypoxemia to hospitalization for acute care or death is a clinically significant metric. A 29-day period to assess the primary endpoint is appropriate as the median time for COVID-19 related hospital admission has been reported to be 7-11 days and the median time to clinical deterioration, 9-12 days (Huang 2020; Zhou 2020). Key secondary endpoints include evaluations of safety and tolerability for IM administration of sotrovimab, pharmacokinetics and comparability of anti-viral activity of IM versus IV sotrovimab

The study was planned to first evaluate non-inferiority of 500 mg of sotrovimab given IM versus 500mg of sotrovimab given IV. If non-inferiority was demonstrated, an additional non-inferiority evaluation was to be performed of 250mg IM of sotrovimab versus 500mg IV of sotrovimab.

An *ad hoc* meeting of the IDMC was called on on 11-August-2021. The IDMC concurred that recruitment to the sotrovimab 250 mg IM arm be permanently discontinued. The enrollment for sotrovimab 250 mg IM arm was terminated afterwards. Therefore the non-inferiority evaluation of 250mg IM of sotrovimab versus 500mg IV of sotrovimab will not be performed (See Section 9.1).

#### 2.2. Background

A novel betacoronavirus was first reported in December 2019 causing severe pneumonia in Wuhan, China. Since that time, SARS-CoV-2 has spread throughout the world leading to over 145 million confirmed cases and over 3 million deaths as of April 24, 2021 (COVID-19 dashboard, Johns Hopkins).

There is an urgent need for effective treatment throughout the course of illness. Emerging data suggest that morbidity and mortality associated with COVID-19 exceeds that of influenza and risk factors for progression to severe disease and mortality, include lifestyle and demographic factors, and the presence of pre-existing comorbidities. (Gold 2020; Jordan 2020; Weiss 2020; Zheng 2020) Reported case fatality rates vary markedly by country, with current estimates of overall rates ranging from 1.8% to 3.4% and mortality rates exceeding 25% for some populations. (Bialek 2020 Ferguson 2020) Based on current experience in China, the UK, and Italy, experts have estimated that approximately 30% of those who are hospitalized will require critical care and that 50% of those in critical care will die. In addition, an age-dependent proportion of those that do not require critical care will also die (Ferguson 2020). Mortality rates are likely to be impacted by access to appropriate care as intensive care unit (ICU) beds and critical care capacity becomes overwhelmed.

Not only is there a high unmet need for treatment of those with severe disease but also for therapies designed to prevent the progression of disease in those with early infection. Currently available data suggest that SARS-CoV-2 is associated with a relatively long incubation period, with mean incubation period from exposure to symptomatic disease of approximately 5 days with the 95% percentile of the distribution at 12.5 days (Li 2020). In a study of two cohorts of adult inpatients with COVID-19, the median time to progression to severe disease (ICU admission/development of ARDS) from onset-of-symptoms was 12 days (range 8 to 15 days) with a median duration from onset to death or discharge of 21 days (range 17 to 25 days) (Zhou 2020). In addition, viral loads are highest early in the course of disease and tend to fall once severe sequelae have ensued, although remain detectable by RT-PCR well into the course of disease (Wölfel 2020; Zou 2020). Notably, although viral nucleic acid was detected well after symptom resolution in participants with mild infection, shedding of infectious virus in sputum was limited to the first week of symptoms (Wölfel 2020).

There are currently three investigational monoclonal antibody products against SARS-CoV-2 that have emergency use authorization (EUA) for the treatment of mild to moderate COVID-19. Casirivimab/imdevimab was granted an EUA on November 21, 2020 for the treatment of mild to moderate COVID-19 in high-risk patients. Bamlanivimab (LY-CoV555) was granted an EUA on November 9$^{th}$, 2020 for the treatment of patients 12 years old and older who are at high risk of progressing to severe COVID-19 and/or hospitalization. However, given the high levels of resistance that has developed in SARS-CoV-2 to monotherapy with bamlanivimab, the FDA revoked the EUA for bamlanimivab on April 16, 2021 (FDA 2021). Therefore, bamlanivimab is only recommended for treatment in combination with etesevimab, which was granted an EUA on February 9, 2021. These mAb products showed a decline in SARS-CoV-2 respiratory viral load with administration and early data suggests that administration can prevent progression to severe disease in these patients (Chen 2020; Eli Lilly 2021; Weinreich 2021). Sotrovimab was granted an EUA on May 26, 2021 (FDA 2021)

Vir Biotechnology, Inc. (Vir) has developed a fully human neutralizing anti-SARS-CoV-2 antibody, sotrovimab (VIR-7831; GSK4182136), which contains a 2 amino acid Fc-modification (“LS”) that is designed to improve bioavailability in the respiratory mucosa and increase half-life. Sotrovimab binds to a conserved epitope on the SARS-CoV and SARS-CoV-2 spike protein outside the receptor-binding motif and has been shown to neutralize pseudovirus and live virus in several independent laboratories (Pinto 2020). This unique binding site may retain activity against emerging SARS-CoV-2 variants that may be resistant to other mAbs (Wang 2021). The epitope to which sotrovimab binds is comprised of 23 amino acids. Amino acids comprising the epitope are highly conserved with >99.89% conservation among >1,000,000 currently available sequences of SARS-CoV-2 (https://www.gisaid.org). In vitro pseudotyped virus assessment shows that the epitope sequence polymorphisms P337H/L/R/T and E340A/K/G confer reduced susceptibility to sotrovimab. The presence of the highly prevalent D614G variant, either alone or in combination, did not alter neutralization of sotrovimab. Pseudotyped virus in vitro assessments indicate that sotrovimab retains activity against the UK (B.1.1.7: H69-, V70-, Y144-, N501Y, A570D, D614G, P681H, T716I, S982A, D1118H), South Africa (B.1.351: L18F, D80A, D215G, R246I, K417N, E484K, N501Y, D614G, A701V), Brazil (P.1: D138Y, D614G, E484K, H655Y, K417T,

L18F, N501Y, P26S, R190S, T1027I, T20N, V1176F) and California (CAL.20C: D614G, L452R, S13I, W152C) variant spike proteins. (Cathcart 2021; MacCallum 2021) Thus, it is expected that sotrovimab will retain activity against most SARS-CoV-2 strains even when given as monotherapy.

In the Phase 3 placebo-controlled trial evaluating the efficacy of sotrovimab in preventing progression of COVID-19 in high-risk patients with mild/moderate COVID-19 (VIR-7831-50001; COMET-ICE), serum PK from the Lead-in phase is available to date. There were 10 participants in the sotrovimab arm of the Lead-in Phase of this study. One participant discontinued early due to withdrawal of consent following infusion with sotrovimab. The mean maximum observed concentration ($C_{max}$) of 500 mg sotrovimab was 219 µg/mL following a 1 hour IV infusion. The mean serum level on Day 29 is 37.2 µg/mL. The mean clearance (CL) and volume of distribution at steady state ($V_{ss}$) were 125 mL/day and 8.1 L, respectively. The median half-life was 48.8 days. The mean PK profile and parameters are presented in Figure 1 and Table 1 respecitvely.
Mean (+ standard deviation [SD]) Sotrovimab Serum Concentration-Time Plots (Linear and Semi-log): COMET-ICE Lead-In Phase

**Figure 1: Mean (+ standard deviation [SD]) Sotrovimab Serum Concentration-Time Plots (Linear and Semi-log): COMET-ICE Lead-In Phase**

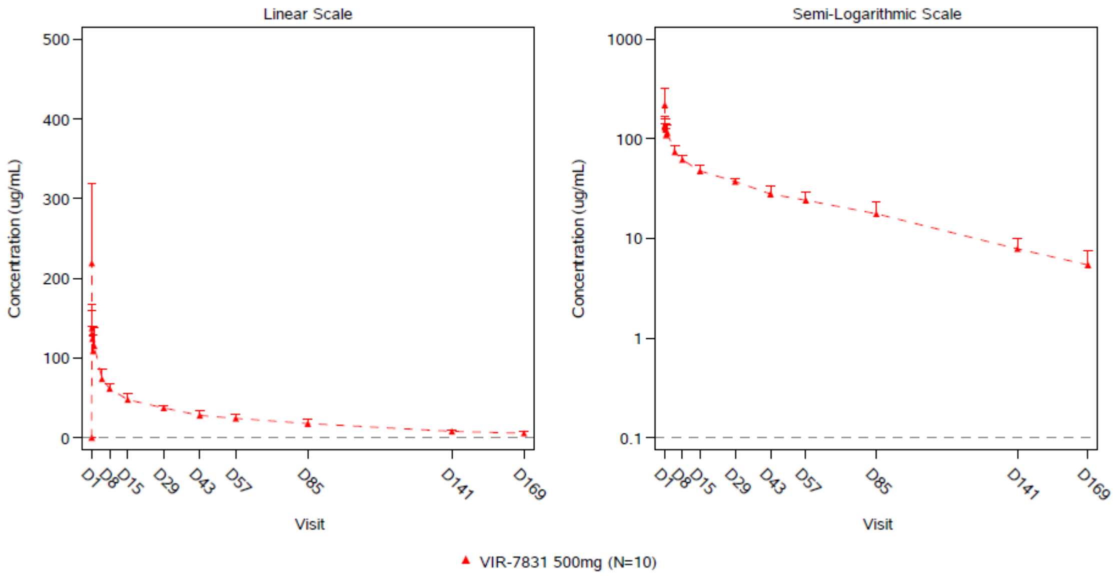

Note: Lower limit of quantification (LLQ)=0.1 µg/mL. Excludes anomalous concentrations identified as sampling errors by the clinical pharmacologist.
Data is based on 1-hour infusion time.

**Table 1: Sotrovimab PK Parameters Following a 500 mg IV Dose: COMET-ICE Lead-in Phase**

| Parameter | Dose 500 mg (N = 9[a]) |
|---|---|
| $C_{max}$, µg/mL | 219 (45.5) |
| $T_{max}$, day | 0.04 (0.04, 0.05) |
| $C_{last}$, µg/mL | 5.41 (37.2) |
| $T_{last}$, day | 161 (160, 167) |
| $AUC_{D1-29}$, day*µg/mL | 1529 (9.6) |
| $AUC_{last}$, day*µg/mL | 3714 (14.5) |
| AUC % Extrapolated | 9.4 (37.9) |
| $AUC_{inf}$, day*µg/mL | 4116 (16.9) |
| CL (mL/day) | 125 (17.9) |
| $V_z$, L | 8.76 (15.7) |
| $V_{ss}$, L | 8.1 (11.1) |
| $t_{1/2}$, day | 48.8 (37.8, 59.4) |

Abbreviations: AUC = area under the curve; AUClast = area under the serum concentration-time curve from time zero to time of last measurable concentration; AUC % Extrapolated = area under the plasma concentration-time curve extrapolated from time to infinity as a percentage of total AUC; AUCD1-29 = area under the serum concentration-time curve from Day 1 to Day 29; AUCinf = area under the serum concentration-time curve from time zero to infinity; Clast = last measurable serum concentration; Cmax = maximum observed concentration; CL = clearance; t1/2 = terminal elimination half life; tmax = time to reach Cmax; tlast = time of the last quantifiable concentration; Vss = volume of distribution at steady state; Vz = apparent volume of distribution during terminal phase.
Note: Parameters are reported as mean (%CV) except for Tmax, Tlast, and t1/2, which are presented as median (min, max).
Data is based on 1-hour infusion time.
[a] N=8 for AUCD1-29 as participant 10016 was missing all PK samples prior to Study Day 5.

Partial sparse serum PK through study Day 29 from 363 participants in the COMET-ICE Expansion phase is available to date. The mean serum concentration of sotrovimab on study Day 29 is 25.8 µg/mL.

Furthermore, sotrovimab has recently been formulated to allow for either intravenous or intramuscular administration. Administration via intramuscular dosing could allow greater access to mAb treatment without the requirement of dedicated infusion centers, as is often required for intravenous administration. IM administration of sotrovimab will be compared to IV administration in this study.

#### 2.3. Benefit/Risk Assessment

Information about the reasonably expected adverse events of sotrovimab may be found in the Investigator’s Brochure.

##### 2.3.1. Risk Assessment

| Potential Risk of Clinical Significance | Summary of Data/Rationale for Risk | Mitigation Strategy |
|---|---|---|
| **Study Intervention: Sotrovimab** | | |
| Immunogenicity | While sotrovimab is a fully human IgG, the development of anti-drug antibodies (ADA) that have the potential to impact safety and/or efficacy are a potential general risk associated with the mAb class of therapeutics. | **Monitoring:**<br>This study will include participant follow-up for a period of 24 Weeks (estimated 5 half-lives) to assess for the potential of immunogenicity (measurement of ADA) as well as if ADA is potentially causally associated with specific safety concerns and/or impact efficacy. |

| Potential Risk of Clinical Significance | Summary of Data/Rationale for Risk | Mitigation Strategy |
|---|---|---|
| Antibody Dependent Enhancement (ADE) due to sub-neutralizing levels of sotrovimab enhancing fusion or leading to FcγR mediated increased viral uptake and replication with virus production | This is a concern related to the potential for participants with sub-neutralizing mAb levels to experience a higher incidence of infection and/or more severe disease compared to participants with no circulating mAb and/or established protective immunity to SARS-CoV-2.<br>ADE associated with Dengue virus 1-4 serotype infections is one of the most widely cited examples in which reinfection with a different serotype can, in a minority of patients, run a more severe course in the setting of limited antibodies generated by prior infection.<br>The potential for enhanced disease in this setting is due to increased uptake of virus by FcR-expressing cells, such as macrophages, and increased viral replication in these cells. Recent data shows that SARS-CoV-2 does not replicate efficiently in macrophages [Hui 2020], suggesting minimal to no risk of ADE via this mechanism.<br><br>As of 10 March 2021, the COMET ICE study enrollment has been stopped based profound efficacy on the recommendation of IDMC. The total of 1057 participants have been randomized to either the IV infusion of sotrovimab Gen1 (500 mg dose) or placebo. Based on the Joint SRT review of blinded data, there has been no confirmed events of ADE. | **Monitoring:**<br>This study will include participant follow up for a period of 36 Weeks to assess for the potential of enhanced disease in the context of waning sotrovimab levels. Assessments for unusually severe disease in mAb-treated participants will be performed by the JSRT to assess for unusually severe disease in mAb-treated participants. Re-infection will be assessed as new SAEs related to COVID-19 infection during follow up period. |

| Potential Risk of Clinical Significance | Summary of Data/Rationale for Risk | Mitigation Strategy |
|---|---|---|
| ADE due to enhanced disease pathology from viral antigen-antibody related immune complex deposition or complement activation and immune cell recruitment in target organs | There is the possibility that a large amount of antibody that binds, but does not neutralize, virus in the presence of a high viral load could result in immune complex deposition and complement activation in tissue sites of high viral replication, such as the lungs, vascular endothelia, renal or cardiovascular tissue [Hamming 2004], leading to tissue damage/immune complex disease<br>This is hypothesized to have contributed to inflammation and airway obstruction observed in the small airways of infants who received a formalin-inactivated (FI) respiratory syncytial virus (RSV) vaccine [Polack 2002] and in a few cases of fatal H1N1 influenza infection [Wu 2010]<br>The potential for enhanced disease in this setting may be due to low affinity or cross-reactive antibodies with poor or no neutralizing activity.<br>Triggering of cytokine release by antibody-virus-FcγR interactions although usually highly beneficial due to their direct antiviral effects and immune cell recruitment to control viral spread in tissues, also has the potential to enhance pathologic changes initiated by the viral infection.<br>Observational data from 20,000 COVID-19 patients treated with convalescent plasma, although not placebo controlled, is suggestive that even polyclonal mixtures of neutralizing and non-neutralizing antibodies can be safely administered [Joyner 2020] | **Monitoring:**<br>• In this study, periodic review of clinical signs and symptoms of COVID-19, clinical chemistry, adverse events, end-organ disease and histopathological diagnoses (as available per routine care) will be performed by the Joint Safety Review Team to identify potential cases of immune complex disease.<br>• Assessments of the overall duration and severity of COVID-19 will be performed by the JSRT to assess for unusually severe disease in mAb-treated participants. |

| Potential Risk of Clinical Significance | Summary of Data/Rationale for Risk | Mitigation Strategy |
|---|---|---|
| | Sotrovimab shows potent binding in vitro as well as neutralization of pseudovirus and live virus thus this risk is deemed to be low.<br><br>As of 10 March 2021, based on data from 583 randomized patients, the COMET-ICE study enrollment has been stopped due to evidence of clinical efficacy based on the recommendation of IDMC. Since then, up to a total of 1057 participants have been randomized to either the IV infusion of sotrovimab Gen1 (500 mg dose) or placebo. Based on the Joint SRT review of blinded data, there has been no confirmed events of ADE.<br><br>The ACTIV-3 TICO study in hospitalized patients with COVID-19 symptoms which was terminated due to futility did not reveal any safety concerns overall and no ADE. | |

| Potential Risk of Clinical Significance | Summary of Data/Rationale for Risk | Mitigation Strategy |
|---|---|---|
| Systemic reactions including hypersensitivity reactions (SR/HSR) | While sotrovimab is a fully human IgG, Systemic reactions SR), including hypersensitivity reactions, are a potential general risk associated with the mAb class of therapeutics.<br>As of 24 April 2021, approximately 1417 participants have been randomized to either the IV infusion of sotrovimab Gen1 (500 mg dose) or placebo in two ongoing clinical studies: 1057 participants in a study evaluating sotrovimab for the treatment of non-hospitalized individuals (COMET ICE Study) with mild to moderate COVID-19, and approximately 360 participants in a study evaluating sotrovimab for the treatment of individuals hospitalized with COVID-19 (ACTIV 3-TICO Study).<br>Based on the assessment of the data, the majority of the SR/HSR in the COMET ICE study have been grade 1 or 2. In the hospitalized study (ACTIV-3 TICO), severe or life-threatening allergic reactions have been infrequently reported during infusion of sotrovimab. There has been a single report of anaphylactic reaction during the IV infusion of sotrovimab.The infusion was stopped and treatment with medications including epinephrine was provided which led to resolution of the event. | **Participant selection**: Participants will be excluded if they have a history of hypersensitivity to other monoclonal antibodies or any of the excipients present in the investigational product.<br>**Monitoring:**<br>• Guidelines for monitoring relevant adverse events encompassing hypersensitivity, angioedema and anaphylaxis as well as for the management of acute anaphylactic shock and minor allergic episodes will be in place at investigational sites.<br>• Joint Safety Review Team reviews will be conducted at regular intervals to determine if a significant safety signal of severe hypersensitivity reaction is identified.<br>• Systemic reactions including HSRs are categorized as AESIs<br>**Mitigation**:<br>Investigators will be provided with general guidance on management of hypersensitivity reactions and such reactions will be managed appropriately per local guidelines/medical judgment. Pre-medications will be permitted at the investigator’s discretion and will be appropriately documented.<br>Vital signs will be monitored prior to dose administration and at 15 mins and 30 mins after infusions/injections.<br>• Investigators will be instructed to discontinue IV infusions or injections for participants who develop Grade 3 or higher systemic reactions. If a participant experiences a Grade 2 SR, |

| Potential Risk of Clinical Significance | Summary of Data/Rationale for Risk | Mitigation Strategy |
|---|---|---|
| | | investigators will be instructed to pause the infusion/injection. The infusion/injection may subsequently resume at a slower pace of infusion, at the investigator's discretion, and/or after symptomatic treatment (e.g., anti-histamines, IV fluids). IV infusion and IM injection will be administered in the clinic with staff trained in emergency care & resuscitation procedures & emergency care kit on hand during treatment & post therapy observation periods. |

| Potential Risk of Clinical Significance | Summary of Data/Rationale for Risk | Mitigation Strategy |
|---|---|---|
| Local Infusion/Injection Site Reactions | Intramuscular injections may be associated with local reactions (e.g. swelling, induration, pain).<br>• A GLP injection site reaction study in minipigs demonstrated tolerability after intramuscular injection<br>• A similar fully human IgG1 monoclonal antibody specific to influenza A (VIR-2482) has been studied in a recent Phase 1 trial. Intramuscular injection of this IgG1 was well-tolerated with minimal injection site reactions (Sager 2020)<br>• Intramuscular dosing of sotrovimab is being evaluated in Part B of the COMET-PEAK study (VIR-7831-5006; GSK216912). This study (COMET-TAIL) will not begin until the JSRT from COMET-PEAK has determined there are no significant local tolerability issues from the Lead-in portion of COMET-PEAK, Part B (N=20 sotrovimab IM recipients) | **Monitoring:**<br>• Local infusion/injection site Reactions will be monitored closely in the Lead-in phase of Part B of the COMET-PEAK GSK216912/VIR-7831-5006 (N=10 in the Gen2 sotrovimab IV arm; N=10 in the Gen 2 sotrovimab IM arm) for 2 hours. Local tolerability data will be assessed by the JSRT. Enrollment into this study will not begin until JSRT review of the Lead-in phase of COMET-PEAK, Part B.<br>• Vital signs will be monitored prior to dose administration and at 15 mins and 30 mins after injection/infusion.<br>• Local infusion/Injection site reactions are categorized as AESIs<br>• Monitor for Local infusion/injection site reactions throughout study.<br>**Mitigation:**<br>Local infusion/injection site reactions will be managed appropriately per local guidelines/medical judgment. |

##### 2.3.2. Benefit Assessment

There is biologic rationale which supports the use of sotrovimab, an antiviral mAb in the early treatment of COVID-19 to prevent progression.

Sotrovimab has been demonstrated in vitro to be a highly potent fully human IgG neutralizing SARS-CoV-2 antibody which has the potential to be an effective therapeutic in mild to critically ill patients with COVID-19. sotrovimab does not bind to normal human tissues in a tissue cross-reactivity study and there were no adverse findings after 2 weeks in the 2-week repeat dose monkey toxicity study. The no -observed -adverse -effect-level (NOAEL) in the monkey study was 500 mg/kg, the highest dose tested.

In addition, intravenous sotrovimab has been shown in a Phase 3 study (COMET-ICE) to be highly effective at preventing progression of mild/moderate COVID-19 when administered to patients who are at high-risk for severe disease. Furthermore, other anti-SARS-CoV-2 mAbs (bamlanivimab +/- etesevimab, casirivimab/imdevimab) have preliminarily been shown to decrease respiratory viral load and prevent progression to hospitalization or severe disease. (Chen 2020; Eli Lilly 2021; Weinreich 2021)

The study population to be enrolled in this study has a high unmet medical need and is similar to the study population from COMET-ICE. Sotrovimab administered intramuscularly may or may not improve the time to clinical response or overall clinical outcome versus IV sotrovimab in an individual participant with mild or moderate COVID-19 who participates in this study. However, there is potential benefit from their participation in this study resulting from their data allowing evaluation of IM sotrovimab as a potential treatment for COVID-19.

##### 2.3.3. Overall Benefit: Risk Conclusion

The overall benefit-risk assessment takes into account the potential benefit of sotrovimab treatment through the potential ability to suppress viral replication and clear infected cells. The benefit in preventing the progression of mild to moderate non-hospitalized COVID 19 in high-risk patients has been demonstrated.

In addition, based on data from ~ 1200 non hospitalized and hospitalized subjects who have received IV infusion of sotrovimab, there have been no significant safety concerns observed. Although there is a theoretical risk of ADE with anti-SARS-CoV2 mAbs in the presence of infection, no evidence of ADE with SARS-CoV-2 has been observed in the available literature to date.

Based on the high unmet medical need and considering the measures taken to minimize risk to participants taking part in this study, the potential risks identified in association with sotrovimab are justified by the anticipated benefits that may be afforded to participants with mild to moderate COVID-19.

### 3. OBJECTIVES AND ENDPOINTS

| Objectives | Endpoints |
|---|---|
| **Primary** | |
| Evaluate the efficacy of two dose levels of intramuscular sotrovimab versus intravenous sotrovimab in preventing the progression of mild/moderate COVID-19 | Progression of COVID-19 through Day 29 as defined by:<br>1. Hospitalization > 24 hours for acute management of illness due to any cause<br>OR<br>2. Death |
| **Secondary** | |
| **Safety**<br>Describe the safety and tolerability of intramuscular and intravenous sotrovimab | • Occurrence of adverse events (AEs)<br>• Occurrence of serious adverse events (SAEs)<br>• Occurrence of adverse events of special interest (AESI)<br>• Occurrence of disease related events (DRE) |
| Assess the immunogenicity of sotrovimab | Incidence and titers (if applicable) of serum anti-drug antibody (ADA) to sotrovimab |
| **Efficacy**<br>Evaluate the efficacy of two dose levels of intramuscular versus intravenous sotrovimab on the progression of mild/moderate COVID-19 | Progression of COVID-19 through Day 29 as defined by:<br>1. Visit to a hospital emergency room for management of illness<br>OR<br>2. Hospitalization for acute management of illness for any duration and for any cause<br>OR<br>3. Death |
| Evaluate the efficacy of two dose levels of intramuscular versus intravenous sotrovimab in preventing COVID-19 respiratory disease progression | Development of severe and/or critical respiratory COVID-19 as manifested by requirement for respiratory support (including oxygen) at Day 8, Day 15, Day 22, and Day 29 |

| Objectives | Endpoints |
|---|---|
| Compare the virologic activity of sotrovimab given IM (two dose levels) or IV in reducing SARS-CoV-2 viral load | • Mean area under the curve of SARS-CoV-2 viral load in nasal secretions as measured by qRT-PCR from Day 1 to Day 8 ($AUC_{D1-8}$)<br>• Change from baseline in viral load by qRT-PCR at Day 8<br>• Proportion of participants with a persistently high SARS-CoV-2 viral load at Day 8 by qRT-PCR |
| **Pharmacokinetics**<br>Assess the pharmacokinetics (PK) of sotrovimab in serum following IV and IM (two dose levels) administration | IV and IM sotrovimab pharmacokinetics (PK) in serum |
| **Exploratory** | |
| Describe the effect of two doses levels of intramuscular versus intravenous sotrovimab on incidence and duration of time on total hospital length of stay (LOS), incidence and duration of time on a ventilator, and ICU length of stay | • Incidence of hospitalization through Day 29<br>• Total hospital LOS<br>• Proportion of participants requiring ICU stay or mechanical ventilation through Day 29<br>• Total ICU LOS |
| Monitor SARS-CoV-2 resistant mutants against sotrovimab | • SARS-CoV-2 resistance mutants to sotrovimab at baseline<br>• Emergence of viral resistance mutants to mAb by SARS-CoV-2 |
| Compare the virologic activity of sotrovimab given IM (two dose levels) or IV in reducing SARS-CoV-2 viral load | • Change from baseline in viral load in nasal secretions by qRT-PCR during follow-up period at Day 5, Day 11, Day 15, Day 22 and Day 29<br>• Undetectable SARS-CoV-2 in nasal secretions by qRT-PCR at Day 3, Day 5, Day 8, Day 11, Day 15, Day 22 and Day 29<br>• Mean area under the curve of SARS-CoV-2 viral load as measured by qRT-PCR from Day 1 to Day 5 ($AUC_{D1-5}$) and Day 1 to 11 ($AUC_{D1-11}$) |
| Compare the effect of different sample collection methods in SARS-CoV-2 viral load (e.g. nasopharyngeal swab, saliva) | • SARS-CoV-2 viral load measured by qRT-PCR |
| Evaluate the effect of sotrovimab on the development of SARS-CoV-2 antibodies | • SARS-CoV-2 anti-N antibody at Day 1 and Day 29 |

| Objectives | Endpoints |
|---|---|
| Describe the effect of sotrovimab on Long COVID symptoms | • Incidence and severity of Long COVID symptoms at Week 12, Week 24 and Week 36 |

### 4. STUDY DESIGN

#### 4.1. Overall Design

This study is a Phase 3 randomized, multi-center, open label, non-inferiority study of intramuscular (IM) versus intravenous (IV) administration of sotrovimab, a monoclonal antibody (mAb) against SARS-CoV-2 for the treatment of mild/moderate COVID-19 in participants aged 12 years and older at high risk of disease progression. The study will randomize participants 1:1:1 to receive a single dose IV infusion of sotrovimab or a single IM injection of sotrovimab at one of two dose levels.

Participants with early, mild/moderate COVID-19 who are not on supplementary oxygen and at risk for disease progression will receive an IV infusion or IM injection of sotrovimab. The IV sotrovimab dose will be 500 mg and will be infused over 15 minutes. The IM sotrovimab dose will be either 250 mg or 500 mg given at a single timepoint. The 250 mg IM dose will be given either as a single 250 mg (4ml) injection in the dorsogluteal muscle or as two 2ml injections in each deltoid muscle. The 500 mg IM dose will be administered as two 4ml injections in each dorsogluteal muscle.

After IV infusion or IM injection, participants will be monitored for 30 minutes with vital signs assessments performed every 15 minutes. Injection site reaction, for IM arms only, will be evaluated at 15 and 30 minutes after dosing.

All participants will be actively monitored on an outpatient basis with frequent collection of nasopharyngeal swabs for virology and blood draws for PK sampling, as well as weekly in-clinic evaluations at Weeks 1, 2, 3, and 4 as detailed in the Schedule of Activities.

Starting at Week 8, participants will be monitored monthly via phone call or in-clinic evaluation to assess for the incidence and severity of subsequent COVID-19 illness, if any, for a total of 36 weeks from dosing.

**Number of Participants:**

Approximately 1020 participants were to be enrolled and randomized 1:1:1 to receive a single 500 mg IV infusion of sotrovimab, a single 500 mg IM injection of sotrovimab, or a single 250 mg IM injection of sotrovimab.

**Study Oversight and Rationale for Change in Study Design in Amendment 3**

An interim analysis (IA) was originally planned for an IDMC to evaluate each dose level of IM sotrovimab versus IV sotrovimab for safety, futility and efficacy when approximately 50% of participants enrolled and reached Day 29. As described in Section 10.5.1, a Joint Safety Review Team was also in place to review in-stream safety data for identification of any safety concerns. During the conduct of the study, the JSRT noted a discrepancy in the rate of progression to hospitalization occurring in the 250 mg IM arm compared with the 500 mg IM and IV arms, and an ad hoc interim dataset was escalated to the IDMC for review. As a result of this review, enrollment into the 250 mg IM arm was discontinued at N=195, and the study design was changed to a 2-arm study with randomization 1:1 into the remaining 2 arms of 500 mg IM and 500 mg IV. The formal planned interim analysis was also eliminated as described in Section 9.5.

**Intervention Groups and Duration:**

Screening assessments will be performed within 48 hours prior to randomization. Screening can occur on the same day as randomization. Eligible participants will be treated with a single IM or IV dose of sotrovimab on Day 1 and followed for up to 36 weeks.

Participants will be stratified based on:

1. Age: 12-17 years old, 18-64 years old, and ≥65 years old
2. COVID-19 Vaccination History: receipt of any COVID-19 vaccine
3. Region of the world due to potential differences in standard of care for COVID-19 management and different circulating SARS-CoV-2 variants.

**Saliva SARS-CoV-2 qRT-PCR substudy**

To evaluate the comparability of SARS-CoV-2 quantitative RT-PCR when performed on saliva versus nasopharyngeal (NP) swabs, an optional sub-study may be performed. Up to 200 subjects will have paired saliva and NP collected at 4 timepoints during the study (D1, D8, D15, D22).

#### 4.2. Scientific Rationale for Study Design

This study is a Phase 3 randomized, multi-center, open label, non-inferiority study of intramuscular (IM) versus intravenous (IV) administration of sotrovimab, a monoclonal antibody (mAb) against SARS-CoV-2 for the prevention of progression of mild/moderate COVID19 in participants 12 years old and older at high risk of medical complications from COVID-19. High-risk adults include participants ≥55 years old regardless of co-morbidities and individuals age 18 or older with comorbidities associated with worse outcomes in adults, including diabetes, obesity (BMI ≥30), chronic kidney disease, congestive heart failure, chronic lung diseases (i.e. chronic obstructive pulmonary disease, moderate to severe asthma requiring steroids, interstitial lung disease, cystic fibrosis, and pulmonary hypertension), sickle cell disease, neurodevelopmental disorders, immunosuppressive disease or receiving immunosuppressive medications, or chronic liver disease. (CDC 2021) Adolescents, aged 12-17 years of age, at high-risk for COVID-19 progression include those with obesity (BMI ≥85th percentile for age/gender), sickle-cell disease, congenital heart disease, neurodevelopmental disorders, medically-related technology dependence such as tracheostomy, ventilator-support, or a gastrostomy tube, or chronic lung diseases (i.e. chronic obstructive pulmonary disease, moderate to severe asthma requiring steroids, interstitial lung disease, cystic fibrosis, and pulmonary hypertension), immunosuppressive disease or immunosuppressive medications, or chronic liver disease. (CDC 2021; Shekerdemian 2020) The study will include outpatients infected with SARS-CoV-2, the virus that causes COVID-19, as confirmed by local laboratory tests and/or point of care tests.

Although children appear to have more mild disease when infected with SARS-CoV-2 and only 0.1%-1.9% of COVID-19 cases result in hospitalization (AAP 2021), a number of risk factors have been identified that are associated with increased risk of severe disease. These include children with complex medical history due to genetic, neurologic, or metabolic conditions or with congenital heart disease. Children with obesity, sickle-cell disease, and chronic lung disease may also be at higher risk of hospitalization. Of note, approximately one-third of hospitalized children are admitted to the intensive care unit. (CDC 2021). Given the potential for severe disease in high-risk children, there is a significant unmet need for a medication that could prevent COVID-19 progression. Since hospitalization rates are somewhat lower in children, the adolescent population in this study will be limited to <10% of the total population.

Sotrovimab is currently being studied in a Phase 3 placebo-controlled trial evaluating the efficacy of sotrovimab in preventing progression of COVID-19 in high-risk patients with mild/moderate COVID-19 (VIR-7831-50001; COMET-ICE). On 10 Mar 2021, the independent data monitoring committee for COMET-ICE recommended that the study be stopped for enrollment due to evidence of clinical efficacy. This was based on an interim analysis of data from 583 patients which demonstrated an 85% (p=0.002) reduction in hospitalization or death in patients receiving sotrovimab compared to placebo (Vir 2021). Sotrovimab was well-tolerated with no safety signals detected. In addition, recently both the NIH COVID-19 Treatment Guidelines and the Infectious Diseases Society of America have both recommended monoclonal antibody therapy treatment for high-risk individuals with mild/moderate COVID-19 (NIH 2021).

In light of recent positive data from COMET-ICE demonstrating overwhelming clinical efficacy of sotrovimab in preventing COVID-19 disease progression and the changes in treatment guidelines noted above, enrollment to placebo is not ethically favorable for participants who are at risk of disease progression as it would unnecessarily subject participants to potential morbidity and mortality despite the availability of an otherwise favorable intervention.

Given the urgent unmet medical need and the ethical considerations of a placebo-controlled design in the setting of already demonstrated clinical efficacy, the design for this current trial is a non-inferiority study comparing the clinical efficacy of IV sotrovimab and two dose levels of IM sotrovimab. The primary endpoint to assess efficacy of treatment is progression of mild/moderate COVID-19 defined by hospitalization >24 hours for acute management of illness or death due to any cause within 29 days of randomization. A 3.5% non-inferiority margin on the risk difference scale will be used. These criteria were selected based on feedback from the FDA after taking into consideration multiple monoclonal antibody programs and their efficacy in preventing progression of COVID-19 in high-risk individuals. The study will first evaluate non-inferiority between 500 mg of sotrovimab given IM versus 500mg of sotrovimab given IV. If non-inferiority is demonstrated, an additional non-inferiority evaluation will be performed between 250 mg IM of sotrovimab and 500 mg IV of sotrovimab.

The transition from outpatient status without hypoxemia to hospitalization for acute care or death is a clinically significant metric. A 29-day period to assess the primary endpoint is appropriate as the median time for COVID-19 related hospital admission in patients who are hospitalised has been reported to be 7-11 days and the median time to clinical deterioration, 9-12 days (Huang 2020; Zhou 2020). Key secondary endpoints include evaluations of safety and tolerability for IM administration of sotrovimab, comparability of anti-viral activity of IV versus IM sotrovimab, and pharmacokinetics.

#### 4.3. Justification for Dose

##### 4.3.1. Sotrovimab

The single 500 mg IV dose of sotrovimab to be evaluated was selected based on extensive nonclinical data and expected human PK extrapolated from cynomolgus monkeys. This IV dose is currently being evaluated in ongoing clinical trials COMET-ICE (NCT04545060), ACTIV-3-TICO (NCT04501978) and BLAZE-4 (NCT04634409), with approximately 700 patients dosed to date.

The 250 and 500 mg IM doses were selected based on in vitro neutralization data, in vitro resistance data, and simulated IM PK based on preliminary IV PK from the 500 mg dose being evaluated in ongoing clinical studies (COMET-ICE, ACTIV-3-TICO, BLAZE-4).

Sotrovimab neutralized SARS-CoV-2 live virus with an average $EC_{90}$ value of 186.3 ng/mL (range: 125.8 – 329.5 ng/mL) (PC-7831-0105). In resistance analyses, no viral breakthrough was observed through 10 passages at fixed concentrations of antibody, indicating the potential for sotrovimab to have a high barrier to resistance (PC-7831-0109). Using an increasing concentration selection method to force resistance emergence, E340A was identified as a monoclonal antibody-resistant mutant (MARM) conferring a >100-fold reduction in susceptibility to sotrovimab. Notably, E340 is 99.9% conserved among available SARS-CoV-2 sequences. Due to the binary nature of the resistance selection results, a specific inhibitory quotient (IQ) was not informed by the resistance profiling.

Based on preliminary IV PK data from the Lead-in phase of an ongoing clinical study evaluating sotrovimab in the early treatment of COVID-19 (COMET-ICE; NCT04545060), the mean serum concentration of sotrovimab following a single 500 mg IV dose is 37.3µg/mL (N=9). Based on the PK data available to date, >20% of the AUC is being extrapolated so final CL, V, AUC and $t_{1/2}$ have not been determined; however a preliminary estimate of the median half-life of sotrovimab is approximately 47 days.

IM doses of 250 and 500 mg were selected to ensure that sotrovimab concentrations in lung are maintained at or above levels anticipated to be neutralizing for the duration of the treatment window. Based on the $EC_{90}$ (0.33 µg/mL) from the highest end of the $EC_{90}$ range (PC-7831-0105), and accounting for the lung:serum ratio for IgG (assumed conservative value of 0.25; reported range 0.25-0.68 for whole lung and interstitial fluid, respectively; Baxter 1994, Covel1 1986, Datta-Mannan 2019, Lobo 2004), and assuming 70% bioavailability following intramuscular administration, IM doses of 250 and 500 mg are expected to maintain serum levels at or above 5x and 10x lung tissue adjusted $EC_{90}$ from 1 day post-dose through the Day 29 primary endpoint, respectively.

Prior clinical experience with a 500 mg IV dose of sotrovimab has been gained in the setting of the early treatment of COVID-19 (COMET-ICE; NCT04545060). Available preliminary blinded safety data from this study has demonstrated an acceptable safety profile in individuals actively infected with SARS-CoV-2. An interim analysis of data from 583 high-risk patients demonstrated an 85% (p=0.002) reduction in hospitalization or death in patients receiving sotrovimab compared to placebo. (Vir 2021)

**Dosing in Adolescents**

The recommended doses in patients aged 12 years and above are the same as adults. Assuming conventional allometric scaling of exposure with bodyweight, and data from the National Examination Nutritional Health Survey (NHANES) on weight for age distribution), the expected overlap in exposure with adults is at least 60%, although the lightest adolescents may have exposures marginally above adults. Any risk of under-dosing adolescents is small. The sponsor therefore proposes the same dosing recommendation for adolescents as adults.

#### 4.4. End of Study Definition

A participant is considered to have completed the study if he/she has completed the Week 36 visit. The end of the study is defined as the date of the last contact of the last participant in the study.

### 5. STUDY POPULATION

Prospective approval of protocol deviations to recruitment and enrollment criteria, also known as protocol waivers or exemptions, is not permitted.

#### 5.1. Inclusion Criteria

Participants are eligible to be included in the study only if all of the following criteria apply:

##### 5.1.1. Age and Risk Factors

1. Participant must be aged 12 years or older at time of consent AND at high risk of progression of COVID-19 based on the presence of one or more of the following risk factors:

    a. For 12-17 years old: diabetes, obesity (BMI ≥85th percentile for age/gender based on CDC growth charts), chronic kidney disease (e.g. eGFR <60), sickle-cell disease, congenital heart disease, neurodevelopmental disorders, chronic lung diseases (i.e. chronic obstructive pulmonary disease, moderate to severe asthma requiring steroids, interstitial lung disease, cystic fibrosis, and pulmonary hypertension), immunosuppressive disease or immunosuppressive medications, or chronic liver disease
    NOTE: Participants age 12-17 years old should not exceed 10% of the total study population

    b. For 18-54 years old: diabetes (requiring medication), obesity (BMI ≥ 30, chronic kidney disease (i.e., eGFR <60 by MDRD), congenital heart disease, congestive heart failure (NYHA class II or more), chronic lung diseases (i.e. chronic obstructive pulmonary disease, moderate to severe asthma requiring steroids, interstitial lung disease, cystic fibrosis, and pulmonary hypertension), sickle cell disease, neurodevelopmental disorders, immunosuppressive disease or receiving immunosuppressive medications, or chronic liver disease

OR

2. Participant ≥ 55 years old, irrespective of co-morbidities

    a. NOTE: Target approximately 20% of participants of the study population ≥ 65 years old

##### 5.1.2. Type of Participant and Disease Characteristics

1. Participants who have a positive SARS-CoV-2 test result within 7 days of randomization (by any validated diagnostic test e.g. RT-PCR, antigen based testing on any specimen type)

AND

2. Oxygen saturation ≥94% on room air

AND

3. Have symptoms of COVID-19 defined by one or more of the following: fever, chills, cough, sore throat, malaise, headache, joint or muscle pain, change in smell or taste, vomiting, diarrhea, shortness of breath on exertion

AND

4. Participant to be dosed less than or equal to 7 days from onset of symptoms to dosing day (D1)

##### 5.1.3. Sex and Contraceptive/Barrier Requirements

1. No gender restrictions
2. Female participants must meet and agree to abide by the following contraceptive criteria. Contraception use by women should be consistent with local regulations regarding the methods of contraception for those participating in clinical studies.

A female participant is eligible to participate if she is not pregnant or breastfeeding, and one of the following conditions applies:

a. Is a woman of non-childbearing potential (WONCBP) as defined in Section 10.4.

OR

b. Is a WOCBP and using a contraceptive method that is highly effective, with a failure rate of <1%, as described in Section 10.4 of the protocol during the study intervention period and for up to 36 weeks after the last dose of study intervention. The investigator should evaluate potential for contraceptive method failure (e.g., noncompliance, recently initiated) in relationship to the first dose of study intervention.

A WOCBP must have a negative highly sensitive pregnancy test (urine or serum as required by local regulations) before the first dose of study intervention. See Section 8.3.8 Pregnancy Testing of the protocol.

- If a urine test cannot be confirmed as negative (e.g., an ambiguous result), a serum pregnancy test is required. In such cases, the participant must be excluded from participation if the serum pregnancy result is positive.
- Additional requirements for pregnancy testing during and after study intervention are located in Section 1.3 of the protocol.

The investigator is responsible for review of medical history, menstrual history, and recent sexual activity to decrease the risk for inclusion of a woman with an early undetected pregnancy

##### 5.1.4. Informed Consent

1. Capable of giving signed informed consent as described in Section 10.1 which includes compliance with the requirements and restrictions listed in the informed consent form (ICF) and in this protocol.

2. OR

3. If participants are not capable of giving written informed consent, alternative consent procedures will be followed as described in Section 10.1.3.
4. Participants <18 years old will be required to sign an assent form in addition to a parent or guardian signing the informed consent.

#### 5.2. Exclusion Criteria

Participants are excluded from the study if any of the following criteria apply:

##### 5.2.1. Medical Conditions

1. Currently hospitalized or judged by the investigator as likely to require hospitalization in the next 24 hours
2. Symptoms consistent with severe COVID-19 as defined by shortness of breath at rest or respiratory distress or requiring supplemental oxygen
3. Participants who, in the judgment of the investigator are likely to die in the next 7 days.
4. Known hypersensitivity to any constituent present in the investigational product

##### 5.2.2. Prior/Concurrent Clinical Study Experience

5. Enrollment in any investigational vaccine study within the last 180 days or any other investigational drug study within 30 days prior to Day 1 or within 5 half-lives of the investigational compound, whichever is longer
6. Enrollment in any trial of an investigational drug, vaccine or device study for SARS-CoV-2/COVID-19 within 90 days prior to Day 1 or within 5 half-lives of the investigational compound, whichever is longer

##### 5.2.3. Other Exclusions

7. Immunocompetent individuals who are fully vaccinated against SARS-CoV-2. A person is considered fully vaccinated if at least 14 days have passed since receiving the final dose in a COVID-19 vaccine series.
8. Receipt of convalescent plasma from a recovered COVID-19 patient or anti-SARS-CoV-2 mAb within the last 3 months.
9. Participants who, in the judgment of the investigator, will be unlikely or unable to comply with the requirements of the protocol through Day 29
10. (France Only) Adult patient protected by the law (under legal guardianship, curatorship or judicial protection or incapable of giving consent personally as defined by Articles L. 1121-5 to L.1121-8, L.1122-1-2 and L. 1122-2 of the French public health code)

#### 5.3. Lifestyle Considerations

Lifestyle considerations are not applicable to this study.

#### 5.4. Pre-Screening

Sites may have the option to pre-screen patients to participate in the clinical study. Sites may pre-screen and administer a SARS-CoV-2 test to patients who meet the Age and Risk Factor in Section 5.1.1, and display symptoms of COVID. Pre-screening is optional and not required by any site.

#### 5.5. Screen Failures

Screen failures are defined as participants who consent to participate in the clinical study but are not subsequently randomized. A minimal set of screen failure information is required to ensure transparent reporting of screen failure participants to meet the Consolidated Standards of Reporting Trials (CONSORT) publishing requirements and to respond to queries from regulatory authorities. Minimal information includes demography, screen failure details, eligibility criteria, any protocol deviations and any serious adverse events (SAEs).

Re-screening may be performed as long as the participant is able to be dosed within 7 days after the onset of COVID-19 symptoms.

### 6. STUDY INTERVENTION(S) AND CONCOMITANT THERAPY

Study intervention is defined as any investigational intervention(s), marketed product(s), placebo, or medical device(s) intended to be administered to a study participant according to the study protocol.

#### 6.1. Study Intervention(s) Administered

Overview of study interventions is provided in Table 3. Detailed instructions for the administration of study drug will be provided in a separate pharmacy manual.

**Table 2: Overview of Study Intervention**

| **Arm Name** | **Sotrovimab IV 500mg** | **Sotrovimab IM 500mg** | **Sotrovimab IM 250mg** |
|---|---|---|---|
| **Intervention Name** | VIR-7831 | VIR-7831 | VIR-7831 |
| **Type** | Biologic | Biologic | Biologic |
| **Dose Formulation** | Solution in single-use vial | Solution in single-use vial | Solution in single-use vial |
| **Unit Dose Strength(s)** | 500 mg/vial (62.5 mg/mL) | 500 mg/vial (62.5 mg/mL) | 500 mg/vial (62.5 mg/mL) |
| **Dosage Level(s)** | 500 mg once | 500 mg once | 250 mg once |
| **Route of Administration** | IV infusion | IM injection | IM injection |
| **IMP and NIMP** | IMP | IMP | IMP |
| **Sourcing** | VIR-7831 will be provided centrally by the Sponsor. | VIR-7831 will be provided centrally by the Sponsor. | VIR-7831 will be provided centrally by the Sponsor. |
| **Dosing instructions** | See Pharmacy Manual | See Pharmacy Manual | See Pharmacy Manual |
| **Special instructions** | Gently mix VIR-7831 prior to withdrawing from vial | Gently mix VIR-7831 prior to withdrawing from vial | Gently mix VIR-7831 prior to withdrawing from vial |
| **Packaging and Labelling** | Study intervention will be provided in a single-use vial in an individual carton and labelled as required per country requirement. | Study intervention will be provided in a single-use vial in an individual carton and labelled as required per country requirement. | Study intervention will be provided in a single-use vial in an individual carton and labelled as required per country requirement. |
| **Current/Former Name(s) or Alias(es)** | VIR-7831, GSK4182136 | VIR-7831, GSK4182136 | VIR-7831, GSK4182136 |

#### 6.2. Preparation/Handling/Storage/Accountability

Instructions for the preparation of study drug will be provided in a separate pharmacy manual.

1. The investigator or designee must confirm appropriate temperature conditions have been maintained during transit for all study intervention received and any discrepancies are reported and resolved before use of the study intervention.
2. Only participants enrolled in the study may receive study intervention and only authorized site staff may supply or administer study intervention. All study interventions must be stored in a secure, environmentally controlled, and monitored (manual or automated) area in accordance with the labeled storage conditions with access limited to the investigator and authorized site staff.
3. The investigator, institution, or the head of the medical institution (where applicable) is responsible for study intervention accountability, reconciliation, and record maintenance (i.e., receipt, reconciliation, and final disposition records).
4. Further guidance and information for the final disposition of unused study intervention are provided in the pharmacy manual.

Under normal conditions of handling and administration, study intervention is not expected to pose significant safety risks to site staff. Take adequate precautions to avoid direct eye or skin contact and the generation of aerosols or mists. In the case of unintentional occupational exposure notify the monitor, medical monitor and/or Sponsor study contact.

#### 6.3. Randomization

All participants will be centrally randomized using an Interactive Web Response System (IWRS). Before the study is initiated, the log in information and directions for the IWRS will be provided to each site.

Participants will be randomized in a 1:1:1 ratio to receive sotrovimab 500 mg IV, sotrovimab 500 mg IM, or sotrovimab 250 mg IM.

Participants will be stratified based on:

1. Age: 12-17 years old, 18-64 years old, and ≥65 years old
2. COVID-19 Vaccination History: receipt of any COVID-19 vaccine
3. Region of the world due to potential differences in standard of care for COVID-19 management and different circulating SARS-CoV-2 variants.

Following the discontinuation of sotrovimab 250 mg IM arm (See Section 9.1), no participants were enrolled and randomized into this arm. The remaining participants were randomized in a 1:1 ratio to receive sotrovimab 500 mg IV or sotrovimab 500 mg IM.

#### 6.4. Study Intervention Compliance

Participants will receive sotrovimab either by IM or IV directly from the investigator or designee, under medical supervision. The date and start and stop times of the dose administered will be recorded in the source documents. The dose of study intervention and study participant identification will be confirmed at the time of dosing by a member of the study site staff other than the person administering the study intervention.

#### 6.5. Dose Modification

There are two potential doses in this study and dose modifications are not applicable. See Section 7.1 for instructions to discontinue study treatment for safety reasons.

#### 6.6. Continued Access to Study Intervention after the End of the Study

COVID-19 is an acute illness and participants are not expected to need continued access to sotrovimab after the end of the study.

#### 6.7. Treatment of Overdose

No specific treatment is recommended for an overdose. The treatment physician may provide supportive measures depending on the symptoms.

In the event of an overdose, the treating physician should:

4. Contact the medical monitor immediately.
5. Closely monitor the participant for AE/SAE and laboratory abnormalities.
6. Document the quantity of the excess dose as well as the duration of the overdosing in the CRF.

#### 6.8. Concomitant Therapy

Any medication or vaccine (including over-the-counter or prescription medicines, recreational drugs, vitamins, and/or herbal supplements) or other specific categories of interest that the participant is receiving at the time of enrollment or receives during the study must be recorded along with:

7. reason for use
8. dates of administration including start and end dates
9. dosage information including dose and frequency

The medical monitor should be contacted if there are any questions regarding concomitant or prior therapy.

Fully vaccinated immunocompetent patients are not eligible for the study. However, partially vaccinated participants are allowed to be enrolled and randomised. A person is considered fully vaccinated if at least 14 days have passed since receiving the final dose in a COVID-19 vaccine series. The specific COVID-19 vaccine received should be recorded along with the date of the vaccine.

##### 6.8.1. Medication Not Permitted During the Study

Receipt of convalescent plasma from a recovered COVID-19 patient or anti-SARS-CoV-2 mAb is not permitted during the study unless these are given as local standard-of-care if a participant is hospitalized.

Receipt of any SARS-CoV-2 vaccine is not permitted within 90 days after dosing per CDC guidelines (https://www.cdc.gov/vaccines/covid-19/info-by-product/clinical-considerations.html).

##### 6.8.2. Permitted Concomitant Medication

All medication that the participant is receiving as local, established standard of care for acute COVID-19 is permitted.

Any concerns regarding the acceptability of potential treatments should be discussed with the medical monitor(s).

### 7. DISCONTINUATION OF STUDY INTERVENTION AND PARTICIPANT DISCONTINUATION/WITHDRAWAL

#### 7.1. Discontinuation of Study Intervention

A participant will be permanently discontinued from completion of drug injection if they experience life-threatening, injection-related reaction including severe allergic or hypersensitivity reactions, severe cytokine release syndrome, or Grade 4 injection site reaction (Section 10.4).

In rare instances, it may be necessary for a participant to permanently discontinue study intervention. If study intervention is permanently discontinued, the participant will remain in the study to be evaluated for follow-up assessments. See the Schedule of Activities in Section 1.3 for data to be collected at the time of discontinuation of study intervention and follow-up and for any further evaluations that need to be completed.

##### 7.1.1. Temporary Discontinuation

If a participant experiences a mild allergic or hypersensitivity reaction after the first injection, investigators will be instructed to pause prior to the second injection. A subsequent injection may be administered at the investigator's discretion, and/or after symptomatic treatment (e.g. anti-histamines, IV fluids).

#### 7.2. Participant Discontinuation/Withdrawal from the Study

- A participant may withdraw from the study at any time at his/her own request, at the request of their LAR (legally authorized representative) or may be withdrawn at any time at the discretion of the investigator for safety, behavioral, or compliance reasons. This is expected to be uncommon.
- At the time of withdrawal from the study, if possible, an early withdrawal (EW) visit should be conducted, as shown in the Schedule of Activities (Section 1.3). Participants may be contacted by phone. See Schedule of Activities for data to be collected at the time of study discontinuation and follow-up and for any further evaluations that need to be completed.
- If the participant withdraws consent or the LAR requests that the participant is withdrawn for disclosure of future information, the sponsor/designee may retain and continue to use any data collected before such a withdrawal of consent.
- If the participant withdraws from the study, he/she or the LAR may request destruction of any samples taken and not tested, and the investigator must document this in the site study records.

#### 7.3. Lost to Follow Up

A participant will be considered lost to follow-up if he or she repeatedly fails to return for scheduled visits and is unable to be contacted by the study site.

The following actions must be taken if a participant fails to return to the clinic for a required study visit:

- The site must attempt to contact the participant and reschedule the missed visit as soon as possible and counsel the participant on the importance of maintaining the assigned visit schedule and ascertain whether or not the participant wishes to and/or should continue in the study.
- Before a participant is deemed lost to follow up, the investigator or designee must make every effort to regain contact with the participant (where possible, 3 telephone calls and, if necessary, a certified letter to the participant's last known mailing address or local equivalent methods). These contact attempts should be documented in the participant's medical record.
- If participants cannot be reached after 3 telephone calls at least 24 hours apart, their listed secondary contact person(s) or health care provider will be contacted.
- Should the participant continue to be unreachable, he/she will be considered to have withdrawn from the study.

Discontinuation of specific sites or of the study as a whole are described in Section 10.1.9.

### 8. STUDY ASSESSMENTS AND PROCEDURES

This section lists the parameters of each planned study assessment.

- Study procedures and their timing are summarized in Section 1.3 (Schedule of Activities).
- Select follow-up visits as noted in the Schedule of Activities are planned to be phone call from site to participants. Follow-up visits may be performed at the clinic or as a home visit.
- Protocol waivers or exemptions are not allowed.
- Immediate safety concerns should be discussed with the medical monitor immediately upon occurrence or awareness to determine if the participant should continue or discontinue study intervention.
- Adherence to the study design requirements, including those specified in the Schedule of Activities, is essential and required for study conduct.
- All screening evaluations must be completed and reviewed to confirm that potential participants meet all eligibility criteria. The investigator will maintain a screening log to record details of all participants screened and to confirm eligibility or record reasons for screening failure, as applicable. Re-screening may be performed as long as the participant is able to be dosed within 7 days after the onset of COVID-19 symptoms.
- Procedures conducted as part of the participant's routine clinical management (e.g., blood count and COVID-19 testing) and obtained before signing of ICF may be utilized for screening or baseline purposes provided the procedure met the protocol-specified criteria and was performed within the time frame defined in the Schedule of Activities.
- Repeat or unscheduled samples may be taken for safety reasons or for technical issues with the samples.
- Clinical Research Associates will review the participants' records to verify that prioritized information in the records matches the information entered in the Electronic Data Capture (EDC) system. If required, in the event of a Quality Assurance audit, auditor(s) may be granted access to records.

#### 8.1. Screening Period

Informed consent must be obtained before conducting any study procedures. Screening will be performed within 48 hours prior to randomization and include the assessments outlined in the Schedule of Activities (Section 1.3).

Screening visit and Day 1 visit may occur on the same day.

##### 8.1.1. Medical History

Relevant medical history within the last three years, as determined by the Investigator, should be reported. Details regarding illnesses and allergies, date(s) of onset, and whether condition(s) is currently ongoing will be collected for all participants and should be updated prior to dosing.

##### 8.1.2. SARS-CoV-2 Diagnostic Testing

Documentation of laboratory-confirmed SARS-CoV-2 infection via a validated molecular diagnostic test or antigen test from any respiratory specimen collected ≤ 7 days prior to study entry must be confirmed for eligibility. This can include tests conducted in a Clinical Laboratory Improvement Amendments (CLIA) certified laboratory or equivalent or from a diagnostic test that has received an Emergency Use Authorization from the FDA (FDA 2021).

Participants with a negative test prior to screening, who are tested again at screening and are positive for SARS-CoV-2 can be included as long as the participant has had symptoms ≤ 7 days from dosing.

##### 8.1.3. Secondary Contact Information

In order to minimize the potential for missing data related to the primary endpoint of mortality or need for hospitalization, sites should collect participant contact information for two secondary contacts (e.g., caregiver, family member, friend). The site may also request health care provider contact information and medical care facilities the participant is likely to go to if they get sick.

Contact information for secondary contacts or health care provider will not be recorded in any eCRF. Contact information should be reviewed and updated at each clinic visit, home visit, and during site phone calls.

#### 8.2. Efficacy Assessments

##### 8.2.1. Hospitalization and Death Data Collection

A hospitalization event and the clinical care that is received during a hospitalization as well as death are components of primary, secondary, and exploratory endpoints. Data from the hospitalization and/or death should be captured in the electronic data capture (EDC) system including but not limited to:

1. Serious Adverse Event (SAE) form
2. Dates that the participant is hospitalized and discharged
3. Dates that the participant is admitted to an intensive care unit
4. Details on the amount of and type of supplemental oxygen and/or ventilatory support that the participant received
5. Date, time, and cause of death

##### 8.2.2. SARS-CoV-2 Serology Analysis

In order to evaluate the effect of sotrovimab treatment on seroconversion and the development of host anti-SARS-CoV-2 antibodies, blood for testing of anti-N SARS-CoV-2 antibodies will be collected at baseline and at Week 4. Since sotrovimab may interfere with anti-spike SARS-CoV-2 tests, antibodies directed against the nucleocapsid protein will be measured.

##### 8.2.3. Phone Call for Subsequent COVID-19 Illness

To monitor participants for subsequent COVID-19 illness after Day 29, participants will be called at Week 8, Week 16, Week 28, Week 32 and Week 36. This phone call will assess whether the participant was diagnosed again with COVID-19 and whether this illness resulted in any healthcare encounters. Any medications given as a result of this illness will also be recorded.

##### 8.2.4. Long COVID Assessment

After first being infected with SARS-CoV-2, some people experience a wide range of symptoms, e.g., fatigue, difficulty thinking or concentrating, cough, depression, etc., that can last weeks or months, known as Long COVID or Post-Acute Sequelae of SARS-CoV-2 infection (CDC 2021; NIH 2021). To monitor for development of Long COVID, participants will be asked a series of questions during the Week 12, Week 24 and Week 36/EOS visit to evaluate a range of symptoms in the past 30 days. These questions will assess whether or not the participant has experienced persistent symptoms related to their COVID-19 diagnosis, types of symptoms, and severity of symptoms. The questions can be asked and completed via phone call or during the in-clinic visit (See Section 10.6).

#### 8.3. Safety Assessments

Planned time points for all safety assessments are provided in the Schedule of Activities (Section 1.3).

##### 8.3.1. Physical Examinations

A complete physical examination will be performed on Day 1. For all other visits the physical examination will be symptom-directed as outlined in the Schedule of Activities.

- A complete physical examination will include, at a minimum, assessments of the Skin, Cardiovascular, Respiratory, and Abdominal systems.
- Height and weight will also be measured and recorded at Screening. Body mass index (BMI) will be calculated from these measurements.
- Investigators should pay special attention to clinical signs related to previous serious illnesses.

##### 8.3.2. Vital Signs

On Day 1, vital signs should be recorded prior to dose administration and at 15 mins and 30 mins after injection or infusion.

- Vital sign measurements will include blood pressure, pulse rate, temperature (oral preferred), respiratory rate, and oxygen saturation.

- Blood pressure and pulse measurements will be assessed while participant is semi-supine or sitting with a completely automated device. Manual techniques will be used only if an automated device is not available.
- Blood pressure and pulse measurements should be preceded by at least 5 minutes of rest for the participant in a quiet setting without distractions (e.g., television, cell phones).

##### 8.3.3. Electrocardiograms

At Screening, a single 12-lead ECG is required ONLY for participants with past medical history of a cardiovascular condition such as an arrhythmia, coronary artery disease, congestive heart failure, valvular disease OR diabetes mellitus.

Electrocardiograms will be performed locally as outlined in the Schedule of Activities (see Section 1.3). The review of the ECG printed at the time of collection must be documented. Any new clinically relevant finding on ECGs should be reported as an AE.

Before each ECG test, the participant should be at rest for approximately 10 minutes. The participant should be in the semi-recumbent or supine position.

##### 8.3.4. Local Tolerability Assessment

A local tolerability assessment will be performed for participants who received an IM injection of sotrovimab per the Schedule of Activities. Injection site(s) should be monitored for pain, tenderness, redness, and swelling. A local tolerability assessment tool is provided in (Section 10.5). This is not applicable for IV sotrovimab recipients.

The injection site(s) will be marked and mapped for later observation and should be documented. The tolerability assessment will be performed at approximately 15 minutes and 30 minutes following study drug administration and at Days 3, 5, and 8 per the Schedule of Activities (Section 1.3).

At the discretion of the investigator, unscheduled visits are permitted as needed for follow up of any unresolved local tolerability symptoms.

##### 8.3.5. Virologic Measures

Samples for virological analysis will be collected in accordance with the laboratory manual and Schedule of Activities (Section 1.3).

- Nasopharyngeal swabs will be collected for SARS-CoV-2 RT-PCR
- Saliva will also be collected in an optional substudy for SARS-CoV-2 RT-PCR
- Samples may also be used for resistance surveillance analysis
- Day 1 samples may be used to evaluate for other respiratory infections

##### 8.3.6. Resistance Analyses

In order to monitor for resistance, resistance surveillance will be conducted for all participants who are randomized and dosed. Deep sequencing analysis of the SARS-CoV-2 spike gene will be attempted to identify substitutions in the mAb epitope or substitutions outside the epitope that arise during treatment. NP samples from baseline and the last evaluable post-baseline visit at Day 8 or later will be subjected to deep sequencing analysis, including participants who do not experience a decline in viral load or who experience virologic rebound. Virologic rebound will be defined as participants who experience an increase of >1 $log_{10}$ copies/mL in viral load for 2 consecutive visits, or viral load becomes quantifiable for 2 consecutive visits after having been below the limit of quantification. For identified substitutions that qualify for phenotypic analysis, in vitro phenotypic analysis of the antiviral activity of sotrovimab using a SARS-CoV-2 spike pseudovirus system will be attempted and analyzed for reduced susceptibility to sotrovimab.

##### 8.3.7. Clinical Safety Laboratory Assessments

- See Section 10.2 for the list of clinical laboratory tests to be performed and to the Schedule of Activities (Section 1.3) for the timing and frequency.
- Laboratory assessments at Screening visit will be performed locally at the clinical site as determined necessary by the investigator or as required by local regulations.
- All protocol-required laboratory test performed locally and/or centrally, as defined in Section 10.2, must be conducted in accordance with the laboratory manual and Schedule of Activities (Section 1.3)
  - The investigator must review the laboratory report, document this review, and record any clinically significant changes occurring during the study as an AE. The laboratory reports must be filed with the source documents.
  - Abnormal laboratory findings associated with the underlying disease are not considered clinically significant, unless judged by the investigator to be more severe than expected for the participant’s condition.
  - All laboratory tests with values considered by the investigator to be clinically significantly abnormal during participation in the study should be repeated until the values return to normal or baseline or are no longer considered significantly abnormal by the investigator or medical monitor.
  - If clinically significant values do not return to normal/baseline within a period of time judged reasonable by the investigator, the etiology should be identified, and the sponsor notified.
- If laboratory values from non-protocol specified laboratory tests performed at the institution’s local laboratory require a change in participant management or are considered clinically significant by the investigator (e.g., SAE or AE), then the results must be recorded.

##### 8.3.8. Pregnancy Testing

- Refer to Section 5.1 Inclusion Criteria for pregnancy testing entry criteria.
- Pregnancy testing (urine or serum as required by local regulations) should be conducted at Screening to confirm eligibility and at Week 24 or Early Withdrawal visit. If serum pregnancy test is required, it will be performed locally.
- Additional serum or urine pregnancy tests may be performed, as determined necessary by the investigator or required by local regulation, to establish the absence of pregnancy at any time during the participant's participation in the study.

#### 8.4. Adverse Events (AEs), Serious Adverse Events (SAEs) and Other Safety Reporting

The definitions of adverse events (AE) or serious adverse events (SAEs) can be found in Section 10.3.

The definitions of unsolicited and solicited adverse events can be found in Section 10.3.

AEs will be reported by the participant (or, when appropriate, by a caregiver, surrogate, or the participant's legally authorized representative).

Hospitalization or death are typically considered SAEs and they are included in the primary endpoint. However, if a hospitalization or adverse event is related to expected progression, signs, or symptoms of COVID-19 (as detailed in Section 8.4.8) or if hospitalization is due to elective treatment of a pre-existing condition that did not worsen from baseline as noted in Section 10.3 it will not be considered as an SAE. These events will be collected and reported as SAEs only if the event is more severe than expected for the participant's current clinical status and medical history or if the investigator feels that it is related to study drug. These will be collected and reported as SAEs as delineated in Section 8.4.1 below. Since it will not be possible to delineate in a single participant whether the hospitalization is directly related to COVID-19 complications or could be related to sotrovimab causing more severe disease due to ADE, all hospitalizations for management of acute illness, regardless of cause, will be included in the primary endpoint.

The investigator and any qualified designees are responsible for detecting, documenting, and reporting events that meet the definition of an AE or SAE and remain responsible for following up AEs that are serious, considered related to the study intervention or the study, or that caused the participant to discontinue the study (see Section 7). As noted in Section 8.2.1 data on hospitalization or death should additionally be recorded in the eCRF for all relevant sections.

The method of recording, evaluating, and assessing causality of AEs and SAEs and the procedures for completing and transmitting SAE reports are provided in Section 10.3.

##### 8.4.1. Time Period and Frequency for Collecting AE and SAE Information

- All AEs will be collected from dose administration through Week 12 post-dose. SAEs will be collected from dose administration through the Week 36 follow-up visit at the time points specified in the Schedule of Activities (Section 1.3). However, any SAEs assessed as related to study participation (e.g., study intervention, protocol-mandated procedure, invasive tests or change in existing therapy) will be recorded from the time participant consents to participate in the study.
- Medical occurrences that begin before the start of study intervention but after obtaining informed consent will be recorded as Medical History/Current Medical Conditions not as AEs.
- All SAEs will be recorded and reported to the sponsor or designee immediately and under no circumstance should this exceed 24 hours, as indicated in Section 10.3. The investigator will submit any updated SAE data to the sponsor or designee within 24 hours of it being available.
- Investigators are not obligated to actively seek information on AEs or SAEs after the conclusion of the study participation. However, if the investigator learns of any SAE, including a death, at any time after a participant has been discharged from the study, and he/she considers the event to be reasonably related to the study intervention or study participation, the investigator must promptly notify the sponsor or designee.

##### 8.4.2. Assessment of Severity

Standard toxicity grading according to the *DAIDS Table for Grading the Severity of Adult and Pediatric Adverse Events*, version 2.1 (July 2017) will be used to grade all AEs.

##### 8.4.3. Method of Detecting AEs and SAEs

Care will be taken not to introduce bias when detecting AE and/or SAE. Open-ended and non-leading verbal questioning of the participant is the preferred method to inquire about AE occurrence.

##### 8.4.4. Follow-up of AEs and SAEs

After the initial AE/SAE report, the investigator is required to proactively follow each participant at subsequent visits/contacts. All SAEs and non-serious AEs of special interest (as defined in Section 8.4.8), will be followed until the event is resolved, stabilized, otherwise explained, or the participant is lost to follow-up (as defined in Section 7.3). Further information on follow-up procedures is given in Section 10.3.

##### 8.4.5. Regulatory Reporting Requirements for SAEs

PPD and GlaxoSmithKline (GSK) are acting on behalf of Vir for the purposes of global safety reporting for this study as outlined in the PPD/Vir Safety Medical Management Plan and Expedited Safety Reporting Documents.

- Prompt notification by the investigator to PPD of an SAE is essential so that legal obligations and ethical responsibilities towards the safety of participants and the safety of a study intervention under clinical investigation are met.
- The sponsor or designee has a legal responsibility to notify both the local regulatory authorities and other regulatory agencies about the safety of a study intervention under clinical investigation. The sponsor or designee will comply with country-specific regulatory requirements relating to safety reporting to the regulatory authority, Institutional Review Boards (IRB)/Independent Ethics Committees (IEC), and investigators. Details for notification of expedited and periodic safety reporting can be found in the Expedited and Periodic Safety Reporting Plan (ESRP).
- An investigator who receives an investigator safety report describing an SAE or other specific safety information (e.g., summary or listing of SAEs) as outlined in the ESRP will review and then file it along with the Investigator’s Brochure and will notify the IRB/IEC, if appropriate according to local requirements.
- Investigator safety reports must be prepared for suspected unexpected serious adverse reactions (SUSAR) according to local regulatory requirements and GSK policy and forwarded to investigators as necessary.

##### 8.4.6. Pregnancy

- Details of all pregnancies in female participants will be collected after the start of study intervention and until Week 36.
- If a pregnancy is reported, the investigator will record pregnancy information on the appropriate form and submit it to sponsor or designee within 24 hours of learning of the female participant pregnancy. While pregnancy itself is not considered to be an AE or SAE, any pregnancy complication or elective termination of a pregnancy for medical reasons will be reported as an AE or SAE.
- Abnormal pregnancy outcomes (e.g., spontaneous abortion, fetal death, stillbirth, congenital anomalies, ectopic pregnancy) are considered SAEs and will be reported as such.
- The participant will be followed to determine the outcome of the pregnancy and for up to 1 year after birth. The investigator will collect follow-up information on the participant and the neonate/child and the information will be forwarded to the sponsor or designee.
- Any post-study pregnancy-related SAE considered reasonably related to the study intervention by the investigator will be reported to the sponsor or designee as described in Section 8.4.1. While the investigator is not obligated

to actively seek this information in former study participants, he or she may learn of an SAE through spontaneous reporting.

##### 8.4.7. Cardiovascular and Death Events

For any cardiovascular events detailed in Section 10.3 and all deaths, whether or not they are considered SAEs, specific Cardiovascular (CV) and Death sections of the CRF will be required to be completed. These sections include questions regarding cardiovascular (including sudden cardiac death) and non-cardiovascular death.

The CV CRFs are presented as queries in response to reporting of certain CV MedDRA terms. The CV information should be recorded in the specific cardiovascular section of the CRF within one week of receipt of a CV Event data query prompting its completion.

The Death CRF is provided immediately after the occurrence or outcome of death is reported. Initial and follow-up reports regarding death must be completed within one week of when the death is reported.

##### 8.4.8. Disease-Related Events and/or Disease-Related Outcomes Not Qualifying as AEs or SAEs

Adverse events related to expected progression, signs, or symptoms of COVID-19, unless more severe than expected for the participant's current clinical status and medical history, or resulting in hospitalization as noted in Section 8.4 should not be reported as an AE or SAE.

However, if the underlying disease (i.e., progression) is greater than that which would normally be expected for the clinical course of the disease and/or the patient's clinical status, or if the investigator considers that there was a causal relationship between treatment with study treatment(s) or protocol design/procedures and the disease progression, then this must be reported as an AE or SAE.

For example, the following constitute events NOT meeting the AE definition and that should be considered as expected progression, signs, or symptoms of COVID-19:

- hypoxemia due to COVID-19 requiring supplemental oxygen
- hypoxemia due to COVID-19 requiring non-invasive ventilation or high flow oxygen devices
- respiratory failure due to COVID-19 requiring invasive mechanical ventilation or extracorporeal membrane oxygenation (ECMO)

**NOTE:** If either of the following conditions apply, then the event must be recorded and reported as an AE or SAE (instead of a disease-related event):

- The event is, in the investigator's opinion, of greater intensity, frequency, or duration than expected for the natural history of the disease, or
- The investigator considers that there is a reasonable possibility that the event was related to treatment with study treatment(s).

##### 8.4.9. Adverse Events of Special Interest

Adverse events of special interest are defined in the study protocol as relevant known toxicities of other therapeutic mAbs or as a result of signals observed from previous studies in the nonclinical programs of sotrovimab that will be monitored by the Sponsor either during or at the end of the study. These will be updated during the course of the study based on accumulating safety data.

AESI include:

- Systemic Infusion and Injection-related reactions including hypersensitivity reactions occurring on same day as infusions
- Local infusion/injection site reactions
- Immunogenicity related adverse drug reactions
- Adverse events potentially related to antibody dependent enhancement of disease

###### 8.4.9.1. Systemic Reactions and Hypersensitivity Reactions

Please refer to local or institutional guidelines for monitoring relevant adverse events encompassing hypersensitivity, angioedema, anaphylaxis, acute anaphylactic shock and minor allergic episodes. Pre-medications will be permitted at the investigator’s discretion and will be appropriately documented.

###### 8.4.9.2. Local infusion/injection site reactions (LI/ISR)

Injections may be associated with local reactions (e.g. swelling, induration, pain). Local tolerability assessment is discussed in Section 8.3.3.

###### 8.4.9.3. Immunogenicity

Unwanted immunogenicity is an immune response by an organism against a therapeutic protein. This reaction leads to production of anti-drug-antibodies (ADA) which may inactivate the therapeutic effects of the treatment and, in rare cases, induce adverse events. This study will include participant follow-up for a period of 5 half-lives to assess for the development of ADA and potential impacts on safety, PK and/or efficacy.

###### 8.4.9.4. Antibody Dependent Enhancement

Antibody-dependent enhancement (ADE) of disease theoretically can occur via one of three previously described mechanisms:

6. By facilitating viral entry into host cells and enhancing viral replication in these cells;
7. By increasing viral fusion with target host cells, enhancing viral replication in these cells.
8. By enhancing disease pathology from viral antigen-antibody related immune complex deposition or complement activation and immune cell recruitment in target organs;

The first two mechanisms are hypothesized to occur at sub-neutralizing antibody concentrations (Arvin 2020). This study will include participant follow-up for a period of 5 half-lives to assess for the potential of enhanced disease in the context of waning sotrovimab levels, which may manifest as an increased incidence of re-infection or increased severity of re-infections after recovery from initial illness. The third mechanism is hypothesized to occur at high levels of antigen (i.e., viral load) and antibody potentially leading to immune complex deposition and complement activation in tissue sites of high viral replication. This may manifest as acute deterioration temporally associated with sotrovimab dosing or as increased severity or duration of illness in sotrovimab-treated participants vs. placebo-treated participants.

As of 10 March 2021, enrollment into the COMET ICE study, evaluating sotrovimab for the treatment of non-hospitalized individuals with mild to moderate COVID-19, has been stopped based on the recommendation of IDMC. The total of 1057 participants have been randomized to either the IV infusion of sotrovimab Gen1 (500 mg dose) or placebo. Based on the Joint SRT review of blinded data, there has been no confirmed events of ADE. The ACTIV-3 TICO study of sotrovimab treatment in hospitalized patients with COVID-19 symptoms, which was terminated due to futility, did not reveal any safety concerns overall and no ADE.

As described in Section 2.3.1, AEs potentially related to ADE of the disease will be reviewed by the JSRT to see if there are unusually severe manifestations of COVID-19 in treated individuals.

#### 8.5. Pharmacokinetics

Blood samples for serum PK will be collected as detailed in the Schedule of Activities (Section 1.3). Instructions for the collection and handling of biological samples will be provided by the sponsor or designee in the laboratory manual.

- The actual date and time (24-hour clock time) of each sample will be recorded.
- Samples collected for analyses of sotrovimab serum concentration may also be used to evaluate safety or efficacy aspects related to concerns arising during or after the study.
- At visits during which whole blood samples are collected to obtain serum endpoints other than PK for sotrovimab, one sample of sufficient volume can be used.

Serum concentrations may be combined with data from other studies evaluating sotrovimab for the purpose of population PK model development. Pharmacokinetic analyses may be conducted to explore exposure-response relationships between PK parameters and selected antiviral variables. These analyses may include graphical plots, tabular summaries, and various linear and/or nonlinear analyses. Details of the PK analyses will be provided in the analysis plan.

#### 8.6. Immunogenicity Assessments

Antibodies to sotrovimab will be evaluated in serum samples collected from all participants according to the Schedule of Activities (Section 1.3). Additionally, serum samples should also be collected at the final visit from participants who discontinued study intervention or were withdrawn from the study. These samples will be tested by the sponsor or designee.

Serum samples will be screened for antibodies binding to sotrovimab and the titer of confirmed positive samples will be reported. Other analyses may be performed to verify the stability of antibodies to sotrovimab and/or further characterize the immunogenicity of sotrovimab.

The detection and characterization of antibodies to sotrovimab will be performed using a validated assay method by or under the supervision of the sponsor or designee. All samples collected for detection of antibodies to study intervention will also be evaluated for sotrovimab serum concentration to enable interpretation of the antibody data. Antibodies may be further characterized and/or evaluated for their ability to neutralize the activity of the study intervention(s). Samples may be stored for a maximum of 15 years (or according to local regulations) following the last participant's last visit for the study at a facility selected by the sponsor/designee.

Samples will be collected in accordance with the laboratory manual and Schedule of Activities (Section 1.3).

### 9. STATISTICAL CONSIDERATIONS

#### 9.1. Statistical Hypotheses

The primary objective of this study is to evaluate the efficacy of IM sotrovimab versus IV sotrovimab in preventing the progression of mild/moderate COVID-19.

The primary endpoint is progression of COVID-19 through Day 29 as defined by hospitalization > 24 hours for acute management of illness OR death.

Denoting the proportions of participants with progression of COVID-19 by Day 29 in the sotrovimab 500 mg IM arm, sotrovimab 250 mg IM arm and sotrovimab 500 mg IV arm as $P_{IM500}$, $P_{IM250}$ and $P_{IV500}$, respectively, then the null ($H_0$) and alternative ($H_A$) hypotheses are as follows using a non-inferiority margin of 3.5% (per feedback from the FDA, as mentioned in Section 4.2) on the risk difference scale:

1) Sotrovimab 500 mg IM vs sotrovimab 500 mg IV:

$$H_0: P_{IM500} – P_{IV500} \geq 3.5\%. \ H_A: P_{IM500} – P_{IV500} < 3.5\%$$

2) Sotrovimab 250 mg IM vs sotrovimab 500 mg IV:

$$H_0: P_{IM250} – P_{IV500} \geq 3.5\%. \ H_A: P_{IM250} – P_{IV500} < 3.5\%$$

The above two hypotheses will be tested according to the hierarchical testing principal, with more details described in Section 9.6.

An *ad hoc* meeting of the IDMC was called on on 11-August-2021. The IDMC concurred that recruitment to the sotrovimab 250 mg IM arm be permanently discontinued. The enrollment for sotrovimab 250 mg IM arm was terminated afterwards. Therefore hypothesis 2) will not be tested. (See Section 9.5 and Section 9.6).

#### 9.2. Sample Size Determination

Sotrovimab IM participants and sotrovimab IV participants were planned to be enrolled and randomized in a 1:1:1 ratio to receive a single dose IV infusion of sotrovimab or a single IM injection of sotrovimab at one of two dose levels, in an open-label manner.

Approximately 1020 (340 per arm) participants were planned to be randomly assigned to study intervention. This sample size would provide approximately 90% power to demonstrate that IM injection of sotrovimab is non-inferior to IV infusion of sotrovimab in terms of the proportion of participants with progression of COVID-19 through Day 29 with a one-sided 2.5% type I error rate, assuming COVID-19 progression rates of 2% in the sotrovimab IM and IV arms and 3.5% non-inferiority margin on the risk difference scale.

Following the discontinuation of sotrovimab 250 mg IM arm (See Section 9.1), no more participants were enrolled and randomized into this arm. The remaining participants were randomized in a 1:1 ratio to receive sotrovimab 500mg IV or sotrovimab 500 mg IM.

**Sample Size/Power Sensitivity**

The table below shows the study power for showing non-inferiority for different true progression rates on the sotrovimab IV arm and when the progression rate in sotrovimab IM arm (either sotrovimab 500 mg IM or sotrovimab 250 mg IM) is equal to or higher than that on IV, given a sample size of N=340 per arm.

| | **Progression rate in sotrovimab IV arm** | | |
|---|---|---|---|
| Progression rate in sotrovimab IM arm minus progression rate in sotrovimab IV arm | 1.0% | 1.5% | 2.0% |
| 0% | 99% | 95% | 90% |
| +0.1% | 98% | 94% | 87% |
| +0.2% | 97% | 91% | 85% |
| +0.3% | 95% | 90% | 82% |

#### 9.3. Analysis Sets

For the purposes of analysis, the following mainanalysis sets are defined:

| **Participant Analysis Set** | **Description** |
|---|---|
| Intent-to-Treat (ITT) | All participants who were randomized. Participants who were randomized under protocol amendment Version 1 and were immunocompetent and fully vaccinated will be excluded from ITT population. Participants will be analyzed according to the route of administration and dose level they were randomized to: sotrovimab IM (500 mg), sotrovimab IM (250 mg) or sotrovimab IV (500 mg). |
| Safety | All participants who are randomized and exposed to study intervention. Participants will be analyzed according to the route of administration and dose level they actually received: sotrovimab IM (500 mg), sotrovimab IM (250 mg) or sotrovimab IV (500 mg). |
| Pharmacokinetic | All participants in the Safety analysis set who had at least 1 non-missing PK assessment (post-dose non-quantifiable [NQ] values will be considered as non-missing values) |
| Virology | All participants who were randomized and with a lab confirmed quantifiable nasopharyngeal swab at Day 1. Participants who were randomized under protocol amendment Version 1 and were immunocompetent and fully vaccinated will be excluded from the Virology analysis set. Participants will be analysed according to the route of administration and dose level they were randomized to: sotrovimab IM (500mg), sotrovimab IM (250 mg) or sotrovimab IV (500mg). This will be the primary analysis set for virology. |

#### 9.4. Statistical Analyses

The statistical analysis plan will be finalized prior to first subject first visit (FSFV) and it will include a more technical and detailed description of the statistical analyses described in this section. This section is a summary of the planned statistical analyses of the most important endpoints including primary and key secondary endpoints.

Any change to the data analysis methods described in the protocol will require an amendment only if it changes a principal feature of the protocol. Any other change to the data analysis methods described in the protocol and the justification for making the change will be described in the statistical analysis plan. Additional exploratory analyses of the data will be conducted as deemed appropriate.

##### 9.4.1. General Considerations

In the case of a difference between the stratification assigned at the time of randomization and the data collected in the eCRF, the analyses will be performed based on the data collected in the eCRF, not the stratum assigned at randomization.

Two-sided 95% confidence intervals will be used unless otherwise specified.

##### 9.4.2. Primary Endpoint(s)

###### Primary Estimand

The primary efficacy endpoint will primarily be assessed using an estimand that uses a hypothetical strategy to deal with the main intercurrent events as detailed below.

| | |
|---|---|
| **Target Participant Population** | All participants in the ITT population based on sotrovimab 500 mg IV and 500 mg IM arms, excluding participants who did not meet key inclusion/exclusion criteria. Key inclusion/exclusion criteria used to define the primary analysis population will be defined in the statistical analysis plan. |
| **Primary Endpoint** | Progression of COVID-19 through Day 29 as defined by hospitalization > 24 hours for acute management of illness or death |
| **Intercurrent events** | The anticipated intercurrent events are:<br>• Not receiving randomized treatment (e.g. treatment misallocation) or an alternative study treatment<br>• Discontinuation of study intervention as described in Section 7.1<br>• Use of medication not permitted during the study as listed in Section 6.8.1 up to Day 29<br>All intercurrent events will be handled using a hypothetical strategy, i.e. assuming the intercurrent event had not occurred. Further details for the intercurrent events will be provided in the statistical analysis plan. |
| **Summary measure** | Absolute difference in the proportion of participants meeting the primary endpoint |
| **Analysis Method** | The proportion of participants meeting the primary endpoint will be compared between treatments using a generalized linear model (GLM) with identity link, adjusted for treatment group (sotrovimab 500 mg IV and 500mg IM), COVID-19 vaccination history (yes/no), age (12-18 years old, 19 to 64 years old, ≥65 years old), region and gender (female vs. male). If the model does not converge then some covariate(s) may be combined or removed. Further details will be provided in the statistical analysis plan. |
| **Handling of missing data and intercurrent events leading to exclusion of data** | Data observed in the period following an intercurrent event will be excluded from the analysis. Data from this period will be assumed "missing at random" (MAR).<br>For participants that withdraw from the study and do not meet the primary endpoint at the time of withdrawal, data in the post-withdrawal period will also be assumed MAR.<br>Further details will be provided in the statistical analysis plan. |

###### Supplementary Estimands

A supplementary estimand will be conducted in the ITT population by handling all intercurrent events with a treatment policy strategy (i.e. regardless of the intercurrent events occurring), with all the other attributes being the same as in the primary estimand. To estimate this estimand, all data collected in the population of interest up to Day 29 will be included in the analysis. Missing data due to participant withdrawal from study will be handled in the same manner as the primary estimand.

Other sensitivity/supplementary estimands for the primary efficacy endpoint analysis will be considered if deemed needed. Further details will be provided in the statistical analysis plan.

##### 9.4.3. Secondary Endpoint(s)

Full details of all analysis methods for the secondary endpoints will be provided in the statistical analysis plan.

##### 9.4.4. Exploratory Endpoint(s)

Full details of all analysis methods for the exploratory endpoints will be provided in the statistical analysis plan.

##### 9.4.5. Safety Analysis

For safety data, no formal hypotheses will be tested. Occurrence of AEs, non-COVID-19 -related AEs, SAEs, non-COVID 19 related SAEs, and AESIs, including laboratory tests and vital signs will be displayed in the form of listings, frequencies, summary statistics, graphs, and statistical analyses where appropriate. Interpretation will be aided by clinical expertise.

As noted in Section 8.4, since it will not be possible to delineate in a single participant whether the hospitalization is directly related to COVID-19 complications or could be related to sotrovimab causing more severe disease due to ADE, all hospitalizations regardless of cause will be included in the primary endpoint and will also be counted as serious adverse events. To inform on the number and nature of non-COVID-19 adverse events and serious adverse events, additional safety analyses will be performed in which select, pre-specified terms consistent with known progression of COVID-19 disease will be excluded. Details of these and all analyses, including example outputs, will be documented in the statistical analysis plan.

##### 9.4.6. Other Analysis

Full details of all analysis methods of immunogenicity and population pharmacokinetics will be provided in the statistical analysis plan.

#### 9.5. Interim Analysis

The Joint Safety Review Team (JSRT) comprising individuals from Vir and GSK will review safety data at regular intervals throughout the conduct of the study. Details of the JSRT process is recorded in relevant SRT documents.

A single interim analysis (IA) was initially planned to be performed for evaluation of safety, efficacy and futility when approximately 50% of participants are enrolled and reach Day 29. The IA data was to be evaluated by an IDMC. However, prior to the time of the planned analysis, the JSRT noted a discrepancy in the rate of progression to hospitalization occurring in the 250mg IM arm compared with the 500mg IM and IV arms. Upon review of the cumulative data, the JSRT made the decision to pause enrollment into the 250mg IM arm on 04-August-2021 and escalated the issue to the IDMC. An ad hoc meeting of the IDMC was called on 11-August-2021 and based upon their review of the data, the IDMC concurred that enrollment in the 250mg IM arm should be discontinued. As this ad hoc IDMC meeting occurred prior to the timepoint when the formal interim analysis would have occurred and concluded with discontinuing one arm of the study combined with an unexpected rapid increase in the rate of enrollment, it was not feasible to conduct the formal interim analysis.

#### 9.6. Multiple Comparisons and Multiplicity

The full details of the multiplicity strategy of this study will be provided in the in the statistical analysis plan.

There were 2 hypotheses for the primary endpoint:

(1) non-inferiority of sotrovimab 500 mg IM versus sotrovimab 500 mg IV

(2) non-inferiority of sotrovimab 250 mg IM versus sotrovimab 500 mg IV

Hypotheses (1) and (2) will be tested using the primary estimand according to the hierarchical testing principal as follows:

- If the null hypothesis for (1) is rejected, i.e., the upper bound of the two-sided 95% confidence interval for the risk difference between sotrovimab 500 mg IM and sotrovimab 500 mg IV is less than the pre-specified 3.5% non-inferiority margin, then hypothesis for (2) will be tested.
- If the null hypothesis for (1) is not rejected, then hypothesis for (2) will not be tested.

Below is the summary of possible hypothesis testing results regarding non-inferiorities of (1) and (2):

| Which Null Hypothesis is Rejected | Non-inferiority Declared For |
|---|---|
| Hypotheses (1) and (2) | sotrovimab 500 mg IM vs. sotrovimab 500 mg IV<br>and<br>sotrovimab 250 mg IM vs. sotrovimab 500 mg IV |
| Hypothesis (1) but Not (2) | Only sotrovimab 500 mg IM vs. sotrovimab 500 mg IV |
| Hypothesis (1) is NOT rejected[a] | None |

[a] Null hypothesis for (2) won't be tested in this case

As described in Section 9.5, the sotrovimab 250 mg IM arm was discontinued. Therefore, only hypothesis (1) above in Section 9.6 will be tested.

### 10. SUPPORTING DOCUMENTATION AND OPERATIONAL CONSIDERATIONS

#### 10.1. Appendix 1: Regulatory, Ethical, and Study Oversight Considerations

##### 10.1.1. Regulatory and Ethical Considerations

- This study will be conducted in accordance with the protocol and with:
  - Consensus ethical principles derived from international guidelines including the Declaration of Helsinki and Council for International Organizations of Medical Sciences (CIOMS) International Ethical Guidelines
  - Applicable ICH Good Clinical Practice (GCP) Guidelines
  - Applicable laws and regulations
- The protocol, protocol amendments, ICF, IB, and other relevant documents (e.g., advertisements) must be submitted to an IRB/IEC by the investigator and reviewed and approved by the IRB/IEC before the study is initiated.
- Any amendments to the protocol will require IEC/IRB approval before implementation of changes made to the study design, except for changes necessary to eliminate an immediate hazard to study participants.
- Protocols and any substantial amendments to the protocol will require health authority approval prior to initiation except for changes necessary to eliminate an immediate hazard to study participants.
- The investigator will be responsible for the following:
  - Providing written summaries of the status of the study to the IRB/IEC annually or more frequently in accordance with the requirements, policies, and procedures established by the IRB/EC
  - Notifying the IRB/IEC of SAE or other significant safety findings as required by IRB/IEC procedures
  - Providing oversight of the conduct of the study at the site and adherence to requirements of 21 CFR, ICH guidelines, the IRB/IEC, EU Clinical Trials Directive 2001/20/EC or Regulation (EU) No. 536/2014 (if applicable), and all other applicable local regulations

##### 10.1.2. Financial Disclosure

Investigators and sub-investigators will provide the sponsor or designee with sufficient, accurate financial information as requested to allow the sponsor/designee to submit complete and accurate financial certification or disclosure statements to the appropriate regulatory authorities. Investigators and sub-investigators are responsible for providing information on financial interests during the course of the study and for 1 year after completion of the study.

##### 10.1.3. Informed Consent Process

- The investigator or his/her representative will explain the nature of the study to the participant or their legally authorized representative and answer all questions regarding the study.
- Participants must be informed that their participation is voluntary. Participants or their legally authorized representative will be required to sign a statement of informed consent that meets the requirements of 21 CFR 50, local regulations, ICH guidelines, Health Insurance Portability and Accountability Act (HIPAA) requirements, where applicable, and the IRB/IEC or study center.
- The medical record must include a statement that written informed consent was obtained before the participant was enrolled in the study and the date the written consent was obtained. The authorized person obtaining the informed consent must also sign the ICF.
- Participants must be re-consented to the most current version of the ICF(s) during their participation in the study.
- A signed copy of the ICF(s) must be provided to the participant or their legally authorized representative.

Vir (alone or working with others) may use the participant's coded study data and samples and other information to carry out this study; understand the results of this study; learn more about sotrovimab or about the study disease; fulfill legal and regulatory obligations, including reporting safety information about sotrovimab, this study, and the results of this study to regulatory authorities; provide information about the safety and use of sotrovimab to investigators and institutions that plan to administer sotrovimab to patients; publish the results of these research efforts; work with government agencies or insurers to have sotrovimab approved for medical use or approved for payment coverage.

##### 10.1.4. Data Protection

- Participants will be assigned a unique identifier by the sponsor/designee. Any participant records or datasets that are transferred to the sponsor/designee will contain the identifier only; participant names or any information which would make the participant identifiable will not be transferred.
- The participant must be informed that his/her personal study-related data will be used by the sponsor/designee in accordance with local data protection law. The level of disclosure must also be explained to the participant, who will be required to give consent for their data to be used as described in the informed consent.
- The participant must be informed that his/her medical records may be examined by Clinical Quality Assurance auditors or other authorized personnel appointed by the sponsor/designee, by appropriate IRB/IEC members, and by inspectors from regulatory authorities.

##### 10.1.5. Committees Structure

**Independent Data Monitoring Committee (IDMC)**

An IDMC will perform an interim analysis to make recommendations to Vir/GSK as detailed in the IDMC Charter. Details regarding the IDMC process, structure and function will be available in the IDMC charter, which is available upon request.

**Joint Safety Review Team**

A Joint Safety Review Team (JSRT) comprised of team members from clinical research, pharmacovigilance and statistics from Vir and GSK will meet at regular intervals throughout the conduct of the study for review of instream safety data for identification of safety concerns. If a potential safety issue is identified, the JSRT may escalate this to an ad hoc IDMC meeting for further evaluation. The JSRT will monitor injection site reaction data in both of the sotrovimab IM arms to assess local tolerability issues.

##### 10.1.6. Dissemination of Clinical Study Data

- Where required by applicable regulatory requirements, an investigator signatory will be identified for the approval of the clinical study report. The investigator will be provided reasonable access to statistical tables, figures, and relevant reports and will have the opportunity to review the complete study results at a sponsor site or other mutually-agreeable location.
- Sponsor or designee will also provide all investigators who participated in the study with a summary of the study results and will tell the investigators what treatment their study participants received. The investigator(s) is/are encouraged to share the summary results with the study participants, as appropriate.
- Under the framework of the SHARE initiative, the sponsor intends to make anonymized participant-level data from this trial available to external researchers for scientific analyses or to conduct research that can help advance medical science or improve patient care. This helps ensure the data provided by trial participants are used to maximum effect in the creation of knowledge and understanding. Requests for access may be made through www.clinicalstudydatarequest.com.
- Sponsor or its designee will provide the investigator with the randomization codes for their site only after completion of the full statistical analysis.
- The procedures and timing for public disclosure of the protocol and results summary and for development of a manuscript for publication for this study will be in accordance with GSK Policy.
- A manuscript will be progressed for publication in the scientific literature if the results provide important scientific or medical knowledge.

##### 10.1.7. Data Quality Assurance

- All participant data relating to the study will be recorded on printed or electronic CRFs unless transmitted to the sponsor or designee electronically (e.g., laboratory data). The investigator is responsible for verifying that data entries are accurate and correct by physically or electronically signing the CRF.
- Guidance on completion of CRFs will be provided in the data entry guidelines.
- Quality Tolerance limits (QTLs) will be pre-defined in the Integrated Quality Risk Management Plan to identify systematic issues that can impact participant safety and/or reliability of study results. These pre-defined parameters will be monitored during and at the end of the study, and all deviations from the QTLs and remedial actions taken will be summarized in the clinical study report.
- The investigator must permit study-related monitoring, audits, IRB/IEC review, and regulatory agency inspections and provide direct access to source data documents.
- Monitoring details describing strategy including definition of study critical data items and processes (e.g., risk-based initiatives in operations and quality such as Risk Management and Mitigation Strategies and Analytical Risk-Based Monitoring), methods, responsibilities and requirements, including handling of noncompliance issues and monitoring techniques (central, remote, or on-site monitoring) are provided in the Monitoring Plan.
- The sponsor or designee is responsible for the data management of this study including quality checking of the data.
- The sponsor assumes accountability for actions delegated to other individuals (e.g., Contract Research Organizations).
- Records and documents, including signed ICFs, pertaining to the conduct of this study must be retained by the investigator for 25 years from the issue of the final Clinical Study Report (CSR)/equivalent summary unless local regulations or institutional policies require a longer retention period. No records may be destroyed during the retention period without the written approval of the sponsor. No records may be transferred to another location or party without written notification to the sponsor.
- As hospitalization and mortality are both primary study endpoints and serious adverse events, instream data cleaning including updating and reconciliation of hospitalization events between the safety dataset and efficacy dataset will be performed throughout the study duration and any changes or updates to the SAEs of hospitalization or mortality will be reflected in the efficacy dataset.

##### 10.1.8. Source Documents

- Source documents provide evidence for the existence of the participant and substantiate the integrity of the data collected. Source documents are filed at the investigator's site.
- Data reported on the printed CRFs or entered in the electronic eCRFs that are transcribed from source documents must be consistent with the source documents or the discrepancies must be explained. The investigator may need to request previous medical records or transfer records, depending on the study. Also, current medical records must be available.
- Definition of what constitutes source data and its origin can be found in [Investigator Source Data Agreement].
- The investigator must maintain accurate documentation (source data) that supports the information entered in the CRF.
- Study monitors will perform ongoing source data verification to confirm that data entered into the CRF by authorized site personnel are accurate, complete, and verifiable from source documents; that the safety and rights of participants are being protected; and that the study is being conducted in accordance with the currently approved protocol and any other study agreements, ICH GCP, and all applicable regulatory requirements.

##### 10.1.9. Study and Site Start and Closure

###### First Act of Recruitment

The study start date is the date on which the clinical study will be open for recruitment of participants.

###### Study/Site Termination

Vir reserves the right to close a study site or terminate the study at any time for any reason at the sole discretion of Vir. Study sites will be closed upon study completion. A study site is considered closed when all required documents and study supplies have been collected and a study-site closure visit has been performed.

The investigator may initiate study-site closure at any time, provided there is reasonable cause and sufficient notice is given in advance of the intended termination.

Reasons for the early closure of a study site by the sponsor or investigator may include but are not limited to:

For study termination:

- Discontinuation of further study intervention development

For site termination:

- Failure of the investigator to comply with the protocol, the requirements of the IRB/IEC or local health authorities, the sponsor's procedures, or GCP guidelines
- Inadequate or no recruitment of participants (evaluated after a reasonable amount of time) by the investigator
- If the study is prematurely terminated or suspended, the sponsor shall promptly inform the investigators, the IECs/IRBs, the regulatory authorities, and any contract research organization(s) used in the study of the reason for termination or suspension, as specified by the applicable regulatory requirements. The investigator shall promptly inform the participant(s) and should assure appropriate participant therapy and/or follow-up

###### 10.1.10. Publication Policy

- The results of this study may be published or presented at scientific meetings. If this is foreseen, the investigator agrees to submit all manuscripts or abstracts to the sponsor before submission. This allows the sponsor to protect proprietary information and to provide comments.
- The sponsor will comply with the requirements for publication of study results. In accordance with standard editorial and ethical practice, the sponsor will generally support publication of multicenter studies only in their entirety and not as individual site data. In this case, a coordinating investigator will be designated by mutual agreement.
- Authorship will be determined by mutual agreement and in line with International Committee of Medical Journal Editors authorship requirements.

#### 10.2. Appendix 2: Clinical Laboratory Tests

The tests detailed in Table 3 will be performed by the central laboratory and/or the site local laboratory

Additional tests may be performed at any time during the study as determined necessary by the investigator or required by local regulations.

**Table 3: Protocol-Required Safety Laboratory Tests**

| **Laboratory Assessments** | **Parameters** | | | |
|---|---|---|---|---|
| Hematology | Platelet Count<br>RBC Count<br>Hemoglobin<br>Hematocrit | RBC Indices:<br>MCV<br>MCH | | <u>WBC count with Differential</u>:<br>Neutrophils<br>Lymphocytes<br>Monocytes<br>Eosinophils<br>Basophils |
| Clinical Chemistry | BUN | Potassium | Aspartate Aminotransferase (AST)/ Serum Glutamic-Oxaloacetic Transaminase (SGOT) | Total and direct bilirubin |
| | Creatinine [1] | Sodium | Alanine Aminotransferase (ALT)/ Serum Glutamic-Pyruvic Transaminase (SGPT) | Total Protein |
| | Glucose (non-fasting) | Calcium | Alkaline phosphatase | Gamma-glutamyl transferase (GGT) |
| | Carbon dioxide/bicarbonate | Chloride | Lactate dehydrogenase (LDH) | Albumin |
| | Amylase | Lipase | | |
| Coagulation parameters | International Normalized Ratio (INR) time | Prothrombin time (PT) | Partial thromboplastin time (PTT) / Activated PTT (aPTT) | |
| Pregnancy testing | Highly sensitive (Serum/plasma or urine) human chorionic gonadotropin (hCG) pregnancy test (as needed for women of childbearing potential) | | | |

[1] Repeat within 1 week if above the normal range

### 10.3. Appendix 3: AEs and SAEs: Definitions and Procedures for Recording, Evaluating, Follow-up, and Reporting

#### 10.3.1. Definition of AE

| AE Definition |
|---|
| • An AE is any untoward medical occurrence in a clinical study participant, temporally associated with the use of a study intervention, whether or not considered related to the study intervention.<br>**NOTE:** An AE can therefore be any unfavorable and unintended sign (including an abnormal laboratory finding), symptom, or disease (new or exacerbated) temporally associated with the use of a study intervention. |

| Definition of Unsolicited and Solicited AE |
|---|
| • An unsolicited adverse event is an adverse event that was not solicited using a Participant Diary and that is communicated by a participant who has signed the informed consent. Unsolicited AEs include serious and non-serious AEs.<br>• Potential unsolicited AEs may be medically attended (i.e., symptoms or illnesses requiring a hospitalization, or emergency room visit, or visit to/by a health care provider). The participants will be instructed to contact the site as soon as possible to report medically attended event(s), as well as any events that, though not medically attended, are of participant concern. Detailed information about reported unsolicited AEs will be collected by qualified site personnel and documented in the participant's records.<br>• Unsolicited AEs that are not medically attended nor perceived as a concern by participant will be collected during interview with the participants and by review of available medical records at the next visit.<br>• Solicited AEs are predefined such as injection site reactions and systemic events for which the participant is specifically questioned. |

| Events <u>Meeting</u> the AE Definition |
|---|
| • Exacerbation of a chronic or intermittent pre-existing condition including either an increase in frequency and/or intensity of the condition.<br>• New conditions detected or diagnosed after study intervention administration even though it may have been present before the start of the study.<br>• Signs, symptoms, or the clinical sequelae of a suspected intervention- intervention interaction.<br>• Signs, symptoms, or the clinical sequelae of a suspected overdose of either study intervention or a concomitant medication. Overdose per se will not be reported as an AE/SAE unless it is an intentional overdose taken with possible suicidal/self-harming intent. Such overdoses should be reported regardless of sequelae.<br>• "Lack of efficacy" or "failure of expected pharmacological action" per se will not be reported as an AE or SAE. Such instances will be captured in the efficacy assessments. However, the signs, symptoms, and/or clinical sequelae resulting from lack of efficacy will be reported as AE or SAE if they fulfill the definition of an AE or SAE. |

| **Events Meeting the AE Definition** |
|---|
| • Clinically significant changes in laboratory assessments. |

| **Events NOT Meeting the AE Definition** |
|---|
| • Any clinically significant abnormal laboratory findings or other abnormal safety assessments which are associated with the underlying disease, unless judged by the investigator to be more severe than expected for the participant's condition.<br>• Elective treatment of a pre-existing condition that did not worsen from baseline.<br>• The disease/disorder being studied or expected progression, signs, or symptoms of the disease/disorder being studied, unless more severe than expected for the participant's condition.<br>• Medical or surgical procedure (e.g., endoscopy, appendectomy): the condition that leads to the procedure is the AE.<br>• Situations in which an untoward medical occurrence did not occur (social and/or convenience admission to a hospital).<br>• Anticipated day-to-day fluctuations of pre-existing disease(s) or condition(s) present or detected at the start of the study that do not worsen. |

##### 10.3.2. Definition of SAE

| **An SAE is defined as any serious adverse event that, at any dose:** |
|---|
| **a. Results in death** |
| **b. Is life-threatening**<br>The term 'life-threatening' in the definition of 'serious' refers to an event in which the participant was at risk of death at the time of the event. It does not refer to an event, which hypothetically might have caused death, if it were more severe. |
| **c. Requires inpatient hospitalization or prolongation of existing hospitalization**<br>• In general, hospitalization signifies that the participant has been admitted (usually involving at least an overnight stay) at the hospital or emergency ward for observation and/or treatment that would not have been appropriate in the physician's office or outpatient setting. Complications that occur during hospitalization are AE. If a complication prolongs hospitalization or fulfills any other serious criteria, the event is serious. When in doubt as to whether "hospitalization" occurred or was necessary, the AE should be considered serious.<br>• Hospitalization for elective treatment of a pre-existing condition that did not worsen from baseline is not considered an AE. |

| An SAE is defined as any serious adverse event that, at any dose: |
|---|
| **d. Results in persistent or significant disability/incapacity**<br>• The term disability means a substantial disruption of a person's ability to conduct normal life functions.<br>• This definition is not intended to include experiences of relatively minor medical significance such as uncomplicated headache, nausea, vomiting, diarrhea, influenza, and accidental trauma (e.g. sprained ankle) which may interfere with or prevent everyday life functions but do not constitute a substantial disruption. |
| **e. Is a congenital anomaly/birth defect** |
| **f. Other medically significant situations:**<br>• Possible Hy's Law case: ALT≥3xULN AND total bilirubin ≥2xULN (>35% direct bilirubin) or international normalized ratio (INR) >1.5 must be reported as SAE<br>• Medical or scientific judgment should be exercised by the investigator in deciding whether SAE reporting is appropriate in other situations such as significant medical events that may jeopardize the participant or may require medical or surgical intervention to prevent one of the other outcomes listed in the above definition. These events should usually be considered serious.<br>o Examples of such events include invasive or malignant cancers, intensive treatment for allergic bronchospasm, blood dyscrasias, convulsions, or development of intervention dependency or intervention abuse. |

##### 10.3.3. Definition of Cardiovascular Events

| Cardiovascular Events (CV) Definition: |
|---|
| Investigators will be required to fill out the specific CV event page of the CRF for the following AEs and SAEs:<br>• Myocardial infarction/unstable angina<br>• Congestive heart failure<br>• Arrhythmias<br>• Valvulopathy<br>• Pulmonary hypertension<br>• Cerebrovascular events/stroke and transient ischemic attack<br>• Peripheral arterial thromboembolism<br>• Deep venous thrombosis/pulmonary embolism<br>• Revascularization |

##### 10.3.4. Recording and Follow-Up of AE and SAE

| **AE and SAE Recording** |
|---|
| • When an AE/SAE occurs, it is the responsibility of the investigator to review all documentation (e.g. hospital progress notes, laboratory, and diagnostics reports) related to the event.<br>• The investigator will then record all relevant AE/SAE information.<br>• It is **not** acceptable for the investigator to send photocopies of the participant's medical records to sponsor in lieu of completion of the required form.<br>• There may be instances when copies of medical records for certain cases are requested by pharmacovigilance staff. In this case, all participant identifiers, with the exception of the participant number, will be redacted on the copies of the medical records before submission.<br>• The investigator will attempt to establish a diagnosis of the event based on signs, symptoms, and/or other clinical information. Whenever possible, the diagnosis (not the individual signs/symptoms) will be documented as the AE/SAE. |
| **Assessment of Intensity** |
| • The investigator will make an assessment of intensity for each AE and SAE reported during the study and assign it to 1 of the following categories:<br>• Mild: An event that is easily tolerated by the participant, causing minimal discomfort and not interfering with everyday activities.<br>• Moderate: An event that causes sufficient discomfort and interferes with normal everyday activities.<br>• Severe: An event that prevents normal everyday activities. An AE that is assessed as severe should not be confused with an SAE. Severe is a category utilized for rating the intensity of an event; and both AE and SAE can be assessed as severe.<br>• An event is defined as 'serious' when it meets at least 1 of the predefined outcomes as described in the definition of an SAE, NOT when it is rated as severe. |

| **Assessment of Causality** |
|---|
| The investigator is obligated to assess the relationship between study intervention and each occurrence of each AE/SAE.<br>• A "reasonable possibility" of a relationship conveys that there are facts, evidence, and/or arguments to suggest a causal relationship, rather than a relationship cannot be ruled out.<br>• The investigator will use clinical judgment to determine the relationship.<br>• Alternative causes, such as underlying disease(s), concomitant therapy, and other risk factors, as well as the temporal relationship of the event to study intervention administration will be considered and investigated.<br>• The investigator will also consult the Investigator's Brochure (IB) and/or Product Information, for marketed products, in his/her assessment.<br>• For each AE/SAE, the investigator **must** document in the medical notes that he/she has reviewed the AE/SAE and has provided an assessment of causality.<br>• There may be situations in which an SAE has occurred and the investigator has minimal information to include in the initial report to sponsor. However, **it is very important that** |

| Assessment of Causality |
| --- |
| **the investigator always make an assessment of causality for every event before the initial transmission of the SAE data to sponsor.**<br>• The investigator may change his/her opinion of causality in light of follow-up information and send an SAE follow-up report with the updated causality assessment.<br>• The causality assessment is one of the criteria used when determining regulatory reporting requirements. |

| Follow-up of AE and SAE |
| --- |
| • The investigator is obligated to perform or arrange for the conduct of supplemental measurements and/or evaluations as medically indicated or as requested by pharmacovigilance to elucidate the nature and/or causality of the AE or SAE as fully as possible. This may include additional laboratory tests or investigations, histopathological examinations, or consultation with other health care professionals.<br>• If a participant dies during participation in the study or during a recognized follow-up period, the investigator will provide sponsor with a copy of any post-mortem findings including histopathology.<br>• New or updated information will be recorded in the originally submitted documents.<br>• The investigator will submit any updated SAE data to the sponsor or designee within 24 hours of receipt of the information. |

##### 10.3.5. Reporting of SAEs

| SAE Reporting via Electronic Data Collection Tool |
| --- |
| • The primary mechanism for reporting SAEs will be the electronic data collection (EDC) tool.<br>• If the electronic system is unavailable, then the site will use the paper SAE data collection tool to report the event within 24 hours. Details to be provided in the Study Specific SAE Reporting Guidelines.<br>• The site will enter the SAE data into the electronic system as soon as it becomes available.<br>• The investigator or medically-qualified sub-investigator must show evidence within the eCRF (e.g., check review box, signature, etc.) of review and verification of the relationship of each SAE to IP/study participation (causality) within 24 hours of SAE entry into the eCRF.<br>• After the study is completed at a given site, the electronic data collection tool will be taken off-line to prevent the entry of new data or changes to existing data.<br>• If a site receives a report of a new SAE from a study participant or receives updated data on a previously reported SAE after the electronic data collection tool has been taken off-line, then the site can report this information on a paper SAE form (see next section) or to the SAE coordinator by telephone.<br>• Contacts for SAE reporting will be included in the Study Specific SAE Reporting Guidelines |

###### 10.3.6. Expedited Reporting to Health Authorities, Investigators and Ethics Committees

| Expedited Reporting |
|---|
| The Sponsor or their third party delegates will promptly evaluate all serious adverse events and non-serious adverse events to identify and expeditiously communicate new safety findings to investigators, IRBs, ECs, and health authorities based on applicable legislation as per the Expedited and Periodic Safety Reporting Plan.<br><br>To determine reporting requirements for single adverse event cases, the Sponsor or their third party delegate will assess the expectedness of these events using the sotrovimab Investigator's Brochure. |

#### 10.4. Appendix 4: Contraceptive and Barrier Guidance

##### 10.4.1. Definitions:

**Woman of Childbearing Potential (WOCBP)**

Women in the following categories are considered WOCBP (fertile):

1. Following menarche
2. From the time of menarche until becoming post-menopausal unless permanently sterile (see below)

**Notes:**

- A postmenopausal state is defined as no menses for 12 months without an alternative medical cause.
  - A high follicle-stimulating hormone (FSH) level in the postmenopausal range may be used to confirm a postmenopausal state in women not using hormonal contraception or hormonal replacement therapy (HRT). However, in the absence of 12 months of amenorrhea, confirmation with more than one FSH measurement is required.
  - Females on HRT and whose menopausal status is in doubt will be required to use one of the non-estrogen hormonal highly effective contraception methods if they wish to continue their HRT during the study. Otherwise, they must discontinue HRT to allow confirmation of postmenopausal status before study enrollment.
- Permanent sterilization methods (for the purpose of this study) include:
  - Documented hysterectomy
  - Documented bilateral salpingectomy
  - Documented bilateral oophorectomy

For individuals with permanent infertility due to an alternate medical cause other than the above, (e.g., Mullerian agenesis, androgen insensitivity, gonadal dysgenesis), investigator discretion should be applied to determining study entry.

**Note:** Documentation can come from the site personnel’s review of the participant’s medical records, medical examination, or medical history interview.

If fertility is unclear (e.g., amenorrhea in adolescents or athletes) and a menstrual cycle cannot be confirmed before first dose of study intervention, additional evaluation should be considered.

**Woman of Nonchildbearing Potential (WONCBP)**

Women in the following categories are considered WONCBP:

1. Premenopausal female with permanent infertility due to one of the following (for the purpose of this study):
    a. Documented hysterectomy
    b. Documented bilateral salpingectomy
    c. Documented bilateral oophorectomy

For individuals with permanent infertility due to an alternate medical cause other than the above, (e.g., Mullerian agenesis, androgen insensitivity, gonadal dysgenesis), investigator discretion should be applied to determining study entry.

**Note**: Documentation can come from the site personnel’s review of the participant’s medical records, medical examination, or medical history interview.

2. Postmenopausal female

A postmenopausal state is defined as no menses for 12 months without an alternative medical cause.

- A high follicle stimulating hormone (FSH) level in the postmenopausal range may be used to confirm a postmenopausal state in women not using hormonal contraception or hormonal replacement therapy (HRT). However, in the absence of 12 months of amenorrhea, confirmation with more than one FSH measurement is required.
- Females on HRT and whose menopausal status is in doubt must discontinue HRT to allow confirmation of postmenopausal status before study enrollment.

##### 10.4.2. Contraception Guidance:

| **CONTRACEPTIVES ALLOWED DURING THE STUDY INCLUDE:** |
|---|
| **Highly Effective Methods That Have Low User Dependency** *Failure rate of <1% per year when used consistently and correctly.* |
| • Implantable progestogen-only hormone contraception associated with inhibition of ovulation |
| • Intrauterine device (IUD) |
| • Intrauterine hormone-releasing system (IUS) |
| • Bilateral tubal occlusion |
| • Azoospermic partner (vasectomized or due to a medical cause)<br>Azoospermia is a highly effective contraceptive method provided that the partner is the sole sexual partner of the woman of childbearing potential and the absence of sperm has been confirmed. If not, an additional highly effective method of contraception should be used. Spermatogenesis cycle is approximately 90 days.<br>Note: documentation of azoospermia for a male participant can come from the site personnel's review of the participant's medical records, medical examination, or medical history interview.) |
| **Highly Effective Methods That Are User Dependent** *Failure rate of <1% per year when used consistently and correctly.* |
| • Combined (estrogen- and progestogen-containing) hormonal contraception associated with inhibition of ovulation<br>○ oral<br>○ intravaginal<br>○ transdermal<br>○ injectable |
| • Progestogen-only hormone contraception associated with inhibition of ovulation<br>○ oral<br>○ injectable |
| • Sexual abstinence<br>*Sexual abstinence is considered a highly effective method only if defined as refraining from heterosexual intercourse during the entire period of risk associated with the study intervention. The reliability of sexual abstinence needs to be evaluated in relation to the duration of the study and the preferred and usual lifestyle of the participant.* |

#### 10.5. Appendix 5: Local Tolerability Assessment[1]

- Study Visit Day: ____________
- Timepoint: ____________
- Time Performed: ____________
- Injection Location (check ONE):

☐LEFT DORSOGLUTEAL ☐RIGHT DORSOGLUTEAL

☐LEFT DELTOID ☐RIGHT DELTOID

| Local Reaction to Injectable Product | Absent (Grade 0) | Mild (Grade 1) | Moderate (Grade 2) | Severe (Grade 3) | Potentially Life Threatening (Grade 4) |
|---|---|---|---|---|---|
| **Pain** | ☐ Absent | ☐ Does not interfere with activity | ☐ Repeated use of non-narcotic pain reliever > 24 hr or interferes with activity | ☐ Any use of narcotic pain reliever or prevents daily activity | ☐ ED visit or hospitalisation |
| **Tenderness** | ☐ Absent | ☐ Mild discomfort to touch | ☐ Discomfort with movement | ☐ Significant discomfort at rest | ☐ ED visit or hospitalisation |
| **Erythema/ Redness (Length) cm** | ☐ Absent | ☐ 2.5 to 5.0 cm ________ cm | ☐ 5.1 to 10.0 cm ________ cm | ☐ > 10.0 cm ________ cm | ☐ Necrosis or exfoliative dermatitis |
| **Induration/ Swelling** | ☐ Absent | ☐ 2.5 to 5.0 cm and does not interfere with activity ________ cm | ☐ 5.1 to 10.0 cm or interferes with daily activity ________ cm | ☐ > 10.0 cm or prevents daily activity ________ cm | ☐ Necrosis |

_________________________________
First Initial & Surname (print)

______________________________ _______/_______/_______
Signature Date

[1] Toxicity Grading Scale for Healthy Adult and Adolescent Volunteers Enrolled in Preventive Vaccine Clinical Trials 2007, FDA

#### 10.6. Appendix 6: Long COVID Questionnaire

| **In the past 30 days, has the subject experienced any of the following long COVID symptoms? Yes/No** | | | |
|---|---|---|---|
| 1. | Did the subject have a fever? | Yes/No | If Yes,<br><br>What was the grade or severity of the symptom? (Grade 1/2/3; mild/moderate/severe) |
| 2. | Did the subject have a cough? | Yes/No | If Yes,<br><br>What was the grade or severity of the symptom? (Grade 1/2/3; mild/moderate/severe) |
| 3 | Did the subject have a sore throat? | Yes/No | If Yes,<br><br>What was the grade or severity of the symptom? (Grade 1/2/3; mild/moderate/severe) |
| 4 | Did the subject have muscle aches (myalgia)? | Yes/No | If Yes,<br><br>What was the grade or severity of the symptom? (Grade 1/2/3; mild/moderate/severe) |
| 5 | Did the subject have joint pain (arthralgia)? | Yes/No | If Yes,<br><br>What was the grade or severity of the symptom? (Grade 1/2/3; mild/moderate/severe) |
| 6 | Did the subject have fatigue? | Yes/No | If Yes,<br><br>What was the grade or severity of the symptom? (Grade 1/2/3; mild/moderate/severe) |
| 7 | Did the subject have malaise? | Yes/No | If Yes,<br><br>What was the grade or severity of the symptom? (Grade 1/2/3; mild/moderate/severe) |
| 8 | Did the subject have shortness of breath? | Yes/No | If Yes,<br><br>What was the grade or severity of the symptom? (Grade 1/2/3; mild/moderate/severe) |
| 9 | Did the subject have a headache? | Yes/No | If Yes,<br><br>What was the grade or severity of the symptom? (Grade 1/2/3; mild/moderate/severe) |
| 10 | Did the subject have vomiting? | Yes/No | If Yes,<br><br>What was the grade or severity of the symptom? (Grade 1/2/3; mild/moderate/severe) |

| **In the past 30 days, has the subject experienced any of the following long COVID symptoms? Yes/No** | | | |
|---|---|---|---|
| 11 | Did the subject have nausea? | Yes/No | If Yes,<br><br>What was the grade or severity of the symptom? (Grade 1/2/3; mild/moderate/severe) |
| 12 | Did the subject have diarrhea? | Yes/No | If Yes,<br><br>What was the grade or severity of the symptom? (Grade 1/2/3; mild/moderate/severe) |
| 13 | Did the subject have loss of smell? | Yes/No | If Yes,<br><br>What was the grade or severity of the symptom? (Grade 1/2/3; mild/moderate/severe). |
| 14 | Did the subject have loss of taste? | Yes/No | If Yes,<br><br>What was the grade or severity of the symptom? (Grade 1/2/3; mild/moderate/severe) |
| 15 | Did the subject have chills? | Yes/No | If Yes,<br><br>What was the grade or severity of the symptom? (Grade 1/2/3; mild/moderate/severe) |
| 16 | Did the subject have a skin rash? | Yes/No | If Yes,<br><br>What was the grade or severity of the symptom? (Grade 1/2/3; mild/moderate/severe) |
| 17 | Did the subject have difficulty thinking or concentrating/ brain fog? | Yes/No | If Yes,<br><br>What was the grade or severity of the symptom? (Grade 1/2/3; mild/moderate/severe) |
| 18 | Does the subject have chest pain? | Yes/No | If Yes,<br><br>What was the grade or severity of the symptom? (Grade 1/2/3; mild/moderate/severe) |
| 19 | Does the subject have palpitations? | Yes/No | If Yes,<br><br>What was the grade or severity of the symptom? (Grade 1/2/3; mild/moderate/severe) |
| 20 | Does the subject have trouble sleeping? | Yes/No | If Yes,<br><br>What was the grade or severity of the symptom? (Grade 1/2/3; mild/moderate/severe) |

| In the past 30 days, has the subject experienced any of the following long COVID symptoms? Yes/No | | | |
|---|---|---|---|
| 21 | Does the subject have hair loss? | Yes/No | If Yes,<br><br>What was the grade or severity of the symptom? (Grade 1/2/3; mild/moderate/severe) |
| 22 | Has the subject experienced depression? | Yes/No | If Yes,<br><br>What was the grade or severity of the symptom? (Grade 1/2/3; mild/moderate/severe) |
| 23 | Has the subject experienced anxiety? | Yes/No | If Yes,<br><br>What was the grade or severity of the symptom? (Grade 1/2/3; mild/moderate/severe) |
| 24 | Does the subject have impaired mobility? | Yes/No | If Yes,<br><br>What was the grade or severity of the symptom? (Grade 1/2/3; mild/moderate/severe) |
| 25 | Does the subject have other symptoms? | Yes/No | If Yes, specify<br><br>[enter free text] |

Eli Lilly, bamlanivimab EUA Letter of Authorization 090, 09 February, 2021, accessed 11 February 2021

Eli Lilly, Bamlanivimab and Etesevimab EUA Letter of Authorization 094, 09 February 2021, Accessed 11 Feburary 2021

Garg S, Kim L, Whitaker M, et al. Hospitalization rates and characteristics of patients hospitalized with laboratory-confirmed coronavirus disease 2019 - COVID-NET, 14 states, March 1-30, 2020. MMWR Morb Mortal Wkly Rep. 2020;69(15):458-464. https://www.cdc.gov/mmwr/volumes/69/wr/mm6915e3.htm

Gold JAW, Wong KK, Szablewski CM, Patel PR, Rossow J, da Silva J, Natarajan P, Morris SB, Fanfair RN, Rogers-Brown J, Bruce BB, Browning SD, Hernandez-Romieu AC, Furukawa NW, Kang M, Evans ME, Oosmanally N, Tobin-D'Angelo M, Drenzek C, Murphy DJ, Hollberg J, Blum JM, Jansen R, Wright DW, Sewell WM 3rd, Owens JD, Lefkove B, Brown FW, Burton DC, Uyeki TM, Bialek SR, Jackson BR. Characteristics and Clinical Outcomes of Adult Patients Hospitalized with COVID-19 - Georgia, March 2020. MMWR Morb Mortal Wkly Rep. 2020 May 8;69(18):545-550. doi: 10.15585/mmwr.mm6918e1. PMID: 32379729; PMCID: PMC7737948.

Goldstein RH, Walensky RP. The Challenges Ahead With Monoclonal Antibodies: From Authorization to Access. JAMA. 2020 Dec 1;324(21):2151-2152. doi: 10.1001/jama.2020.21872. PMID: 33175110.

Guan WJ, Ni ZY, Hu Y, et al. Characteristics of coronavirus disease 2019 in China. N Engl J Med. 2020. https://www.nejm.org/doi/full/10.1056/NEJMoa2002032

Hamming I, Timens W, Bulthuis ML, Lely AT, Navis G, van Goor H. Tissue distribution of ACE2 protein, the functional receptor for SARS coronavirus. A first step in understanding SARS pathogenesis. J Pathol. 2004 Jun;203(2):631-7. doi: 10.1002/path.1570. PMID: 15141377; PMCID: PMC7167720.

Hershberger E, Sloan S, Narayan K, Hay CA, Smith P, Engler F, Jeeninga R, Smits S, Trevejo J, Shriver Z, Oldach D. Safety and efficacy of monoclonal antibody VIS410 in adults with uncomplicated influenza A infection: Results from a randomized, double-blind, phase-2, placebo-controlled study. EBioMedicine. 2019 Feb;40:574-582. doi: 10.1016/j.ebiom.2018.12.051. Epub 2019 Jan 9. PubMed PMID: 30638863; PubMed Central PMCID: PMC6412085.

Hope T. Multiscale imaging of therapeutic antibody distribution and localization. Presented at: 2019 HIV & HBV Cure Forum; Jul 20-21; Mexico City, Mexico. Accessed 2020 Apr 14. https://www.iasociety.org/Web/WebContent/File/HIV_Cure_Forum_2019/Session5/FT5-2Hope.pdf.

Huang C, Wang Y, Li X, Ren L, Zhao J, Hu Y, Zhang L, Fan G, Xu J, Gu X, Cheng Z, Yu T, Xia J, Wei Y, Wu W, Xie X, Yin W, Li H, Liu M, Xiao Y, Gao H, Guo L, Xie J, Wang G, Jiang R, Gao Z, Jin Q, Wang J, Cao B. Clinical features of patients infected with 2019 novel coronavirus in Wuhan, China. Lancet. 2020 Feb 15;395(10223):497-506. doi: 10.1016/S0140-6736(20)30183-5. Epub 2020 Jan 24. Erratum in: Lancet. 2020 Jan 30;: PMID: 31986264; PMCID: PMC7159299.

Hui KPY, Cheung MC, Perera RAPM, Ng KC, Bui CHT, Ho JCW, Ng MMT, Kuok DIT, Shih KC, Tsao SW, Poon LLM, Peiris M, Nicholls JM, Chan MCW. Tropism, replication competence, and innate immune responses of the coronavirus SARS-CoV-2 in human respiratory tract and conjunctiva: an analysis in ex-vivo and in-vitro cultures. Lancet. 2020 May 07; https://doi.org/10.1016/S2213-2600(20)30193-4.

Hwang IK, Shih WJ, DeCani JS, Statistics In Medicine. Dec 1990 (Volume9, Issue 12, 1439-1445) Group sequential designs using a family of type I error probability spending functions.

Jordan RE, Adab P, Cheng KK. Covid-19: risk factors for severe disease and death. BMJ. 2020 Mar 26;368:m1198. doi: 10.1136/bmj.m1198. PMID: 32217618.

Joyner MJ, Bruno KA, Klassen SA, Kunze KL, et al. Safety Update: COVID-19 Convalescent Plasma in 20,000 Hospitalized Patients. Mayo Clin Proc. 2020; 95. https://mayoclinicproceedings.org/pb/assets/raw/Health%20Advance/journals/jmcp/jmcp_ft95_6_8.pdf.

Klok FA, Kruip MJHA, van der Meer NJM, Arbous MS, Gommers DAMPJ, Kant KM, Kaptein FHJ, van Paassen J, Stals MAM, Huisman MV, Endeman H Thromb Res. 2020; PMID 32291094 Clinical characteristics of 113 deceased patients with coronavirus disease 2019: retrospective study.

Ko S-Y, Pegu A, Rudicell RS, Yang Z-Y, Joyce MG, Chen X, Wang K, Bao S, Kraemer TD, Rath T, Zeng M, Schmidt SD, Todd J-P, Penzak SR, Saunders KO, Nason MC, Haase AT, Rao SS, Blumberg RS, Mascola JR, Nabel GJ. 2014. Enhanced neonatal Fc receptor function improves protection against primate SHIV infection. Nature. 2014 Oct 30; 514(7524):642-5. doi: 10.1038/nature13612.

Lan KKG, DeMets DL. Discrete sequential boundaries for clinical trials. Biometrika 1983;70:659-63.

Li Q, Guan X, Wu P, Wang X, Zhou L, Tong Y, Ren R, Leung KSM, Lau EHY, Wong JY, Xing X, Xiang N, Wu Y, Li C, Chen Q, Li D, Liu T, Zhao J, Liu M, Tu W, Chen C, Jin L, Yang R, Wang Q, Zhou S, Wang R, Liu H, Luo Y, Liu Y, Shao G, Li H, Tao Z, Yang Y, Deng Z, Liu B, Ma Z, Zhang Y, Shi G, Lam TTY, Wu JT, Gao GF, CowlingBJ, Yang B, Leung GM, Feng Z. Early Transmission Dynamics in Wuhan, China, ofNovel Coronavirus-Infected Pneumonia. N Engl J Med. 2020 Mar 26;382(13):1199-1207. doi: 10.1056/NEJMoa2001316. Epub 2020 Jan 29. PubMed PMID: 31995857.

Lim JJ, Derby MA, Zhang Y, Deng R, Larouche R, Anderson M, Maia M, Carrier S, Pelletier I, Girard J, Kulkarni P, Newton E, Tavel JA. A Phase 1, Randomized, Double-Blind, Placebo-Controlled, Single-Ascending-Dose Study To Investigate the Safety, Tolerability, and Pharmacokinetics of an Anti-Influenza B Virus Monoclonal Antibody, MHAB5553A, in Healthy Volunteers. Antimicrob Agents Chemother. 2017 Jul 25;61(8):e00279-17. doi: 10.1128/AAC.00279-17. PMID: 28559255; PMCID: PMC5527589.

Liu Y, Yan LM, Wan L, Xiang TX, Le A, Liu JM, Peiris M, Poon LLM, Zhang W.. Viral dynamics in mild and severe cases of COVID-19. Lancet Infect Dis 2020;doi: 10.1016/S1473-3099(20)30232-2.

Lobo ED, Hansen RJ, Balthasar JP. Antibody pharmacokinetics and pharmacodynamics. J Pharm Sci. 2004 Nov;93(11):2645-68.

McCallum M, De Marco A, Lempp F, Tortorici M, Pinto D, Walls A, Beltramello M, Chen A, Liu Z, Zatta F, Zepeda S, di Iulio S, Bowen J, Montiel-Ruiz M, Zhou J, Rosen L, Bianchi S, Guarino B, Fregni C, Abdelnabi R, Foo S, Rothlauf P, Bloyet L, Benigni F, Cameroni E, Neyts J, Riva A, Snell G, Telenti A, Whelan S, Virgin H, Corti D, Pizzuto M, Veesler D. N-terminal domain antigenic mapping reveals a site of vulnerability for SARS-CoV-2. Biorxiv. 2021 Jan 14. doi: https://doi.org/10.1101/2021.01.14.426475

NICE. COVID-19 rapid guideline: critical care in adults (NG159). https://www.nice.org.uk/guidance/ng159, 2020

NIH. COVID-19 Treatment Guidelines https://www.covid19treatmentguidelines.nih.gov/. Accessed 2020 May, 4

NIH. NIH launches new initiative to study "Long COVID". https://www.nih.gov/about-nih/who-we-are/nih-director/statements/nih-launches-new-initiative-study-long-covid. Accessed 04 Oct 2021

Pablo N Perez-Guzman, Anna Daunt, Sujit Mukherjee et al. Clinical characteristics and predictors of outcomes of hospitalised patients with COVID-19 in a London NHS Trust: a retrospective cohort study (2020), doi: https://doi.org/10.25561/78613

Petrilli et al, medrxiv N=1999 (hosp) OpenSAFELY (NHS data) N=5683 Deaths REF: COVID-19 dashboard, Johns Hopkins https://gisanddata.maps.arcgis.com/apps/opsdashboard/index.html

Pinto D, Park Y, Beltramello M et al. Cross-neutralization of SARS-CoV-2 by a human monoclonal SARS-CoV antibody. Nature. Published online May 18, 2020. https://doi.org/10.1038/s41586-020-2349-y

Polack FP, Teng MN, Collins PL, Prince GA, Exner M, Regele H, Lirman DD, Rabold R, Hoffman SJ, Karp CL, Kleeberger SR, Wills-Karp M, Karron RA. A role for immune complexes in enhanced respiratory syncytial virus disease. J Exp Med. 2002 Sep 16;196(6):859-65. doi: 10.1084/jem.20020781. PMID: 12235218; PMCID: PMC2194058.

Poon A, Powers JH 3rd, Leidy NK, Yu R, Memoli MJ. Using the Influenza Patient-reported Outcome (FLU-PRO) Diary to Evaluate Symptoms of Influenza Viral Infection in a Healthy Human Challenge Model. BMC Infect Dis. 2018 Jul 28;18(1):353. doi: 10.1186/s12879-018-3220-8.

Powers JH, Bacci ED, Leidy NK, Poon JL, Stringer S, Memoli MJ, et al. Performance of the inFLUenza Patient-Reported Outcome (FLU-PRO) Diary in Patients with Influenza-Like Illness (ILI). PLoS One. 2018 Mar 22;13(3):e0194180. doi: 10.1371/journal.pone.0194180. eCollection 2018.

Powers JH, Bacci ED, Guerrero ML, Leidy NK, Stringer S, Kim K, Memoli MJ, et al. Reliability, Validity, and Responsiveness of InFLUenza Patient-Reported Outcome (FLU-PRO©) Scores in Influenza-Positive Patients. Value Health. 2018 Feb;21(2):210-218. doi: 10.1016/j.jval.2017.04.014. Epub 2017 Jun 7.

Regeneron, casirivimab and imdevimab EUA Letter of Authorization, Accessed 11 February 2021

Richardson S, Hirsch JS, Narasimhan M, et al. Presenting Characteristics, Comorbidities, and Outcomes Among 5700 Patients Hospitalized With COVID-19 in the New York City Area. JAMA. Published online April 22, 2020. doi:10.1001/jama.2020.6775.

Sager J, D. Hong, A. Bonavia, L. Connolly, D. Cebrik, M. Fanget, E. Mogalian, P. Griffin. Preliminary safety and pharmacokinetic profile of VIR-2482: a monoclonal antibody for the prevention of influenza A illness. IDWeek Chasing the Sun Poster. Accessed 11 February, 2021

Shekerdemian LS, Mahmood NR, Wolfe KK, Riggs BJ, Ross CE, McKiernan CA, Heidemann SM, Kleinman LC, Sen AI, Hall MW, Priestley MA, McGuire JK, Boukas K, Sharron MP, Burns JP; International COVID-19 PICU Collaborative. Characteristics and Outcomes of Children With Coronavirus Disease 2019 (COVID-19) Infection Admitted to US and Canadian Pediatric Intensive Care Units. JAMA Pediatr. 2020 Sep 1;174(9):868-873. doi: 10.1001/jamapediatrics.2020.1948. PMID: 32392288; PMCID: PMC7489842.

Vir. Press Release. Vir Biotechnology and GSK Announce VIR-7831 Reduces Hospitalization and Risk of Death in Early Treatment of Adults with COVID-19. Issued 2021 March 21.

Wang P, Liu L, Iketani S, Luo Y, Guo Y, Wang M, Yu J, Zhang B, Kwong PD, Graham BS, Mascola JR, Chang JY, Yin MT, Sobieszczyk M, Kyratsous CA, Shapiro L, Sheng Z, Nair MS, Huang Y, Ho DD. Increased Resistance of SARS-CoV-2 Variants B.1.351 and B.1.1.7 to Antibody Neutralization. bioRxiv [Preprint]. 2021 Jan 26:2021.01.25.428137. doi: 10.1101/2021.01.25.428137. PMID: 33532778; PMCID: PMC7852271.

Weinreich DM, Sivapalasingam S, Norton T, Ali S, Gao H, Bhore R, Musser BJ, Soo Y, Rofail D, Im J, Perry C, Pan C, Hosain R, Mahmood A, Davis JD, Turner KC, Hooper AT, Hamilton JD, Baum A, Kyratsous CA, Kim Y, Cook A, Kampman W, Kohli A, Sachdeva Y, Graber X, Kowal B, DiCioccio T, Stahl N, Lipsich L, Braunstein N, Herman G, Yancopoulos GD; Trial Investigators. REGN-COV2, a Neutralizing Antibody Cocktail, in Outpatients with Covid-19. N Engl J Med. 2021 Jan 21;384(3):238-251. doi: 10.1056/NEJMoa2035002. Epub 2020 Dec 17. PMID: 33332778; PMCID: PMC7781102.

Weiss P, Murdoch DR. Clinical course and mortality risk of severe COVID-19. Lancet. 2020 Mar 28;395(10229):1014-1015. doi: 10.1016/S0140-6736(20)30633-4. Epub 2020 Mar 17. PMID: 32197108; PMCID: PMC7138151.

WHO. World Health Organization Incidence of thrombotic complications in critically ill ICU patients with COVID-19. https://www.who.int/health-topics/coronavirus#tab=tab_3 (Carolina WHO checklist reference) 2021. Accessed 11 February, 2021

Wölfel R, Corman VM, Guggemos W, et al. Virological assessment of hospitalized patients with COVID-2019. Nature. 2020 Apr 01. https://doi.org/10.1038/s41586-020-2196-x

Wu JT, Ma ES, Lee CK, Chu DK, Ho PL, Shen AL, Ho A, Hung IF, Riley S, Ho LM, Lin CK, Tsang T, Lo SV, Lau YL, Leung GM, Cowling BJ, Malik Peiris JS. The infection attack rate and severity of 2009 pandemic H1N1 influenza in Hong Kong. Clin Infect Dis. 2010 Nov 15;51(10):1184-91. doi: 10.1086/656740. PMID: 20964521; PMCID: PMC3034199.

Yang H, et al. O0814 Phase IIb study evaluating the efficacy and safety of anti-influenza A monoclonal antibody, CT-P27, in subjects with acute uncomplicated Influenza A Infection. 29th European Congress of Clinical Microbiology and Infectious Diseases. Amsterdam, Netherlands. 2019 Apr 13-16. Poster presentation.

Yu J, Powers JH 3rd, Vallo D, Falloon J. Evaluation of Efficacy Endpoints for a Phase IIb Study of a Respiratory Syncytial Virus Vaccine in Older Adults Using Patient-Reported Outcomes with Laboratory Confirmation. Value Health. 2020 Feb;23(2):227-235. doi: 10.1016/j.jval.2019.09.2747. Epub 2019 Nov 21.

Zheng S, Fan J, Yu F, Feng B, Lou B, Zou Q, Xie G, Lin S, Wang R, Yang X, Chen W, Wang Q, Zhang D, Liu Y, Gong R, Ma Z, Lu S, Xiao Y, Gu Y, Zhang J, Yao H, Xu K, Lu X, Wei G, Zhou J, Fang Q, Cai H, Qiu Y, Sheng J, Chen Y, Liang T. Viral load dynamics and disease severity in patients infected with SARS-CoV-2 in Zhejiang province, China, January-March 2020: retrospective cohort study. BMJ. 2020 Apr 21;369:m1443. doi: 10.1136/bmj.m1443. PMID: 32317267; PMCID: PMC7190077.

Zhou F, Yu T, Du R, Fan G, Liu Y, Liu Z, Xiang J, Wang Y, Song B, Gu X, Guan L, Wei Y, Li H, Wu X, Xu J, Tu S, Zhang Y, Chen H, Cao B. Clinical course and risk factors for mortality of adult inpatients with COVID-19 in Wuhan, China: a retrospective cohort study. Lancet. 2020 Mar 28;395(10229):1054-1062. doi: 10.1016/S0140-6736(20)30566-3. Epub 2020 Mar 11. Erratum in: Lancet. 2020 Mar 28;395(10229):1038. Lancet. 2020 Mar 28;395(10229):1038. PubMed PMID: 32171076.

Zou L, Ruan F, Huang M, Liang L, Huang H, Hong Z, Yu J, Kang M, Song Y, Xia J, Guo Q, Song T, He J, Yen HL, Peiris M, Wu J. SARS-CoV-2 Viral Load in Upper Respiratory Specimens of Infected Patients. N Engl J Med. 2020 Mar 19;382(12):1177-1179. doi: 10.1056/NEJMc2001737. Epub 2020 Feb 19. PubMed PMID: 32074444.

Signature Page for VV-CLIN-000295 v5.0

| | |
|---|---|
| Approval | PPD |

Signature Page for VV-CLIN-000295 v5.0

| **Information Type:** | Statistical Analysis Plan (SAP) |
|---|---|

### TITLE PAGE

**Protocol Title:** A Phase 3 randomized, multi-center, open label study to assess the efficacy, safety, and tolerability of monoclonal antibody VIR-7831 (sotrovimab) given intramuscularly versus intravenously for the treatment of mild/moderate coronavirus disease 2019 (COVID-19) in high-risk non-hospitalized patients

**Study Number:** VIR-7831-5008 (GSK Study 217114)

**Compound Number:** VIR-7831 (sotrovimab; GSK4182136)

**Abbreviated Title:** Intramuscular VIR-7831 for mild/moderate COVID-19

**Sponsor Name:** This study is sponsored by Vir Biotechnology, Inc.
GlaxoSmithKline is supporting Vir Biotechnology, Inc. in the conduct of this study.

Vir Biotechnology, Inc.
499 Illinois Street, Suite 500
San Francisco, CA 94158
USA

**Regulatory Agency Identifier Number(s)**

| **Registry** | **ID** |
|---|---|
| **IND:** | IND-149315 |
| **EudraCT:** | 2021-000623-13 |
| **NCT:** | NCT04913675 |

Copyright 2021 the Vir Biotechnology, Inc and GlaxoSmithKline group of companies. All rights reserved. Unauthorised copying or use of this information is prohibited

#### SAP Authors:

| Author | Date |
|---|---|
| PPD<br>Statistics Leader, Biostatistics, GlaxoSmithKline | 19-Oct-2021 |

#### SAP Team Review Confirmations (E-mail Confirmation):

| Approver | Date of E-mail Confirmation |
|---|---|
| PPD<br>PPD Clinical Research, Vir Biotechnology Inc. | 19-Oct-2021 |
| PPD<br>PPD, SMG Pharma Safety, GlaxoSmithKline | 25-Oct-2021 |
| PPD<br>PPD, Biometrics, Vir Biotechnology Inc. | 19-Oct-2021 |
| PPD<br>PPD, Clinical Pharmacology, Vir Biotechnology Inc. | 22-Oct-2021 |
| PPD<br>Programming Leader, Biostatistics, GlaxoSmithKline | 19-Oct-2021 |

#### Clinical Statistics Line Approval (E-Mail):

| Approver | Date |
|---|---|
| PPD<br>PPD, Biostatistics, GlaxoSmithKline | 22-Oct-2021 |

#### Version history

| SAP Version<br><br>(Approval Date) | Protocol Version (Date) on which SAP is Based | Change | Rationale |
|---|---|---|---|
| 1.0<br><br>(08-Jun-2021) | Amendment 1<br><br>(05-May-2021) | Not Applicable | Original version |
| 2.0<br><br>(25-Oct-2021) | Amendment 3<br><br>(04-Oct-2021) | • Definition of ITT analysis set changed to match that in the protocol<br>• Update to hypothetical estimand to detail key inclusion/exclusion criteria that lead to exclusion from population<br>• Details of planned and ad hoc interim analyses added<br>• Removal of 250mg IM dose from analyses and testing hierarchy<br>• Region and vaccination status removed as covariates from analysis models, and the 12-17 years and 18-64 years age groupings combined in the age group covariate.<br>• Methods for imputing COVID-19 progression status and viral load data corrected and clarified<br>• Details added for cut-point used to define persistently high viral load<br>• Inclusion of long COVID exploratory endpoint<br>• Amend Week 24 timepoint to Week 36 due to increased length of follow-up in the study | Clarifications made to language in SAP to better align to language used in protocol amendments.<br><br>250mg IM arm removed from analyses and testing hierarchy due to being permanently discontinued from study at ad hoc interim analysis.<br><br>Model covariates amended due to small numbers of patients in non-US regions, small proportion of fully/partially vaccinated participants, and small number of participants <18 years. |

| SAP Version<br><br>(Approval Date) | Protocol Version (Date) on which SAP is Based | Change | Rationale |
|---|---|---|---|
| | | • Addition of Kaplan-Meier plot summarizing time to undetectable viral load<br>• Addition of new subgroup and PK-PD analyses. | |

### 1. INTRODUCTION

The purpose of this SAP is to describe the planned analyses to be included in the Clinical Study Report for Study VIR-7831-5008. Details of the final Day 29 efficacy and safety analyses and 36-Week Safety Follow-Up analyses are provided.

Additional detail with regards to data handling conventions and the specification of data displays will be provided in the Output and Programming Specification (OPS) document.

#### 1.1. Objectives, Estimands and Endpoints

| Objectives | Endpoints |
|---|---|
| Primary | |
| • Evaluate the efficacy of two dose levels of intramuscular sotrovimab versus intravenous sotrovimab in preventing the progression of mild/moderate COVID-19 | Progression of COVID-19 through Day 29 as defined by:<br>• Hospitalization > 24 hours for acute management of illness due to any cause<br>OR<br>• Death |
| Secondary | |
| **Safety**<br>• Describe the safety and tolerability of intramuscular and intravenous sotrovimab | • Occurrence of adverse events (AEs)<br>• Occurrence of serious adverse events (SAEs)<br>• Occurrence of adverse events of special interest (AESI)<br>• Occurrence of disease-related events |
| • Assess the immunogenicity of sotrovimab | • Incidence and titers (if applicable) of serum anti-drug antibody (ADA) to sotrovimab |
| **Efficacy**<br>• Evaluate the efficacy of two dose levels of intramuscular versus intravenous sotrovimab on the progression of mild/moderate COVID-19 | Progression of COVID-19 through Day 29 as defined by:<br>• Visit to a hospital emergency room for management of illness<br>OR<br>• Hospitalization for acute management of illness for any duration and for any cause<br>OR<br>• Death |

| Objectives | Endpoints |
|---|---|
| • Evaluate the efficacy of two dose levels of intramuscular versus intravenous sotrovimab in preventing COVID-19 respiratory disease progression | • Development of severe and/or critical respiratory COVID-19 as manifested by requirement for respiratory support (including oxygen) at Day 8, Day 15, Day 22, and Day 29 |
| • Compare the virologic activity of sotrovimab given IM (two dose levels) or IV in reducing SARS-CoV-2 viral load | • Mean area under the curve of SARS-CoV-2 viral load in nasal secretions as measured by qRT-PCR from Day 1 to Day 8 ($AUC_{D1-8}$)<br>• Change from baseline in viral load by qRT-PCR at Day 8<br>• Proportion of participants with a persistently high SARS-CoV-2 viral load at Day 8 by qRT-PCR |
| **Pharmacokinetics**<br>• Assess the pharmacokinetics (PK) of sotrovimab in serum following IV and IM (two dose levels) administration | • IV and IM sotrovimab pharmacokinetics (PK) in serum |
| Exploratory | |
| • Describe the effect of two doses levels of intramuscular versus intravenous sotrovimab on incidence and duration of time on total hospital length of stay (LOS), incidence and duration of time on a ventilator, and ICU length of stay | • Incidence of hospitalization through Day 29<br>• Total hospital LOS<br>• Proportion of participants requiring ICU stay or mechanical ventilation through Day 29<br>• Total ICU LOS |
| • Monitor SARS-CoV-2 resistant mutants against sotrovimab | • SARS-CoV-2 resistance mutants to sotrovimab at baseline<br>• Emergence of viral resistance mutants to mAb by SARS-CoV-2 |
| • Compare the virologic activity of sotrovimab given IM (two dose levels) or IV in reducing SARS-CoV-2 viral load | • Change from baseline in viral load in nasal secretions by qRT-PCR during follow-up period at Day 5, Day 11, Day 15, Day 22 and Day 29 |

| Objectives | Endpoints |
|---|---|
| | • Undetectable SARS-CoV-2 in nasal secretions by qRT-PCR at Day 3, Day 5, Day 8, Day 11, Day 15, Day 22 and Day 29<br>• Mean area under the curve of SARS-CoV-2 viral load as measured by qRT-PCR from Day 1 to Day 5 ($AUC_{D1-5}$) and Day 1 to 11 ($AUC_{D1-11}$) |
| • Compare the effect of different sample collection methods in SARS-CoV-2 viral load (e.g. nasopharyngeal swab, saliva) | • SARS-CoV-2 viral load measured by qRT-PCR |
| • Evaluate the effect of sotrovimab on the development of SARS-CoV-2 antibodies | • SARS-CoV-2 anti-N antibody at Day 1 and Day 29 |
| • Describe the effect of sotrovimab on Long COVID symptoms | • Incidence and severity of Long COVID symptoms at Week 12, Week 24 and Week 36 |

**Primary estimand**

The primary efficacy endpoint will primarily be assessed using an estimand that uses a hypothetical strategy to deal with the main intercurrent events. The estimand is described by the following attributes:

- Population: All participants in the Intent-to-Treat (ITT) analysis set excluding participants who did not meet key inclusion/exclusion criteria (see section 4.2.2.1).
- Endpoint: Progression of COVID-19 through Day 29 as defined by hospitalization > 24 hours for acute management of illness due to any cause or death
- Summary measure: Absolute difference in the proportion of participants meeting the primary endpoint
- The anticipated intercurrent events are:
  - Not receiving randomized treatment or an alternative study treatment (i.e. treatment misallocation)
  - Discontinuation of study intervention as described in Protocol Section 7.1.
  - Use of medication not permitted during the study as listed in Protocol Section 6.8.1 up to Day 29

All intercurrent events will be handled using a hypothetical strategy where data collected after the occurrence of an intercurrent event will be excluded from the analysis, i.e. assuming the intercurrent event had not occurred.

Rationale for estimand: A hypothetical estimand is used as this is considered more conservative than treatment policy and therefore more appropriate for a non-inferiority comparison.

**Supplementary Estimand for the Primary Endpoint**

A supplementary estimand will be conducted based on all participants in the ITT analysis set by handling all intercurrent events with a treatment policy strategy (i.e. regardless of the intercurrent events occurring), with all the other attributes being the same as in the primary estimand. To estimate this estimand, all data collected in the ITT analysis set up to Day 29 will be included in the analysis.

**Estimands for Key Secondary Endpoints**

The estimand for the secondary endpoint 'Mean area under the curve of SARS-CoV-2 viral load in nasal secretions as measured by qRT-PCR from Day 1 to Day 8 ($AUC_{D1-8}$)' is described by the following attributes:

- Population: All participants in the Virology analysis set, as defined in Section 3
- Summary measure: Treatment ratio (IM/IV) of mean $\log_{10}$ $AUC_{D1-8}$
- Intercurrent events: Death and use of medication not permitted during the study as listed in Protocol Section 6.8.1 up to day 8
  - Strategy for Death: Composite strategy, where for participants who die prior to Day 8, the $AUC_{D1-8}$ viral load value will be imputed using the worst $AUC_{D1-8}$ viral load value from other participants in the same treatment arm.
  - The strategy for all the other intercurrent events (as applicable for this endpoint) will be treatment policy where data will be reported as captured.

The secondary endpoint 'Progression of COVID-19 through Day 29 as defined by visit to a hospital emergency room for management of illness OR Hospitalization for acute management of illness for any duration and for any cause OR Death' will be assessed using a hypothetical estimand as described for the primary endpoint. A supplementary treatment policy estimand will also be assessed using the same approach as the treatment policy estimand for the primary endpoint.

#### 1.2. Study Design

**Overview of Study Design and Key Features**

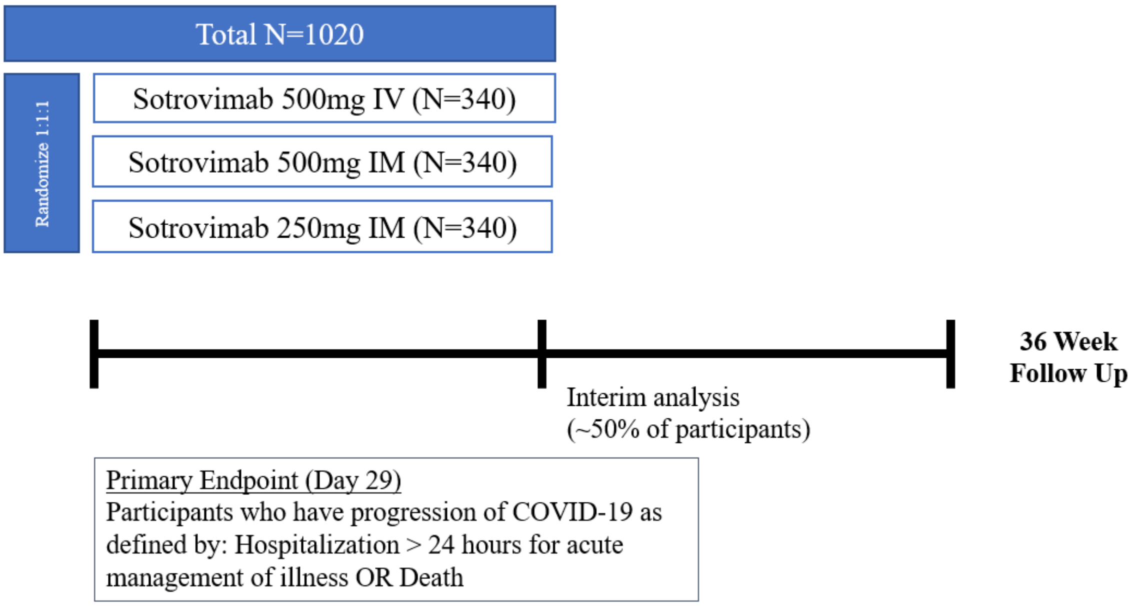

| | |
|---|---|
| **Design Features** | • This study is a Phase 3 randomized, multi-center, open label, non-inferiority study of intramuscular (IM) versus intravenous (IV) administration of sotrovimab, a monoclonal antibody (mAb) against SARS-CoV-2 for the treatment of mild/moderate COVID-19 in participants aged 12 years and older at high risk of disease progression.<br>• Approximately 1020 participants will be enrolled and randomized 1:1:1 to receive a single 500 mg IV infusion of sotrovimab, a single 500 mg IM injection of sotrovimab, or a single 250 mg IM injection of sotrovimab.<br>• Participants with early, mild/moderate COVID-19 who are not on supplementary oxygen and are at risk for disease progression will receive an IV infusion or IM injection of sotrovimab. The intravenous sotrovimab dose will be 500 mg and will be infused over 15 minutes. The intramuscular sotrovimab dose will be either 250 mg or 500 mg given at a single timepoint. The 250 mg IM dose will be given either as a single 250 mg (4ml) injection in the dorsogluteal muscle or as two 2ml injections in each deltoid muscle. The 500 mg IM dose will be administered as two 4ml injections in each dorsogluteal muscle.<br>• After IV infusion or IM injection, participants will be monitored for 30 minutes with vital signs assessments performed every 15 minutes. Injection site reaction assessments, for IM arms only, will be performed at 15 and 30 minutes after injection. |

<table>
<tr><th colspan="2">Overview of Study Design and Key Features</th></tr>
<tr><td></td><td><ul><li>All participants will be actively monitored on an outpatient basis with frequent collection of nasopharyngeal swabs for virology and blood draws for PK sampling and safety labs, as well as in-clinic evaluations at Weeks 1, 2, 3, 4, 12, 20 and 24 as detailed in the Schedule of Activities.</li><li>Starting at Week 8, participants will be monitored monthly via phone call or in-clinic evaluation to assess for the incidence and severity of subsequent COVID-19 illness, if any, for a total of 36 weeks from dosing.</li></ul></td></tr>
<tr><td>Study intervention</td><td><ul><li>VIR-7831 500mg IV or VIR-7831 500mg IM or VIR-7831 250mg IM</li><li>All participants will receive SoC as per institutional protocols, in addition to the study intervention.</li></ul></td></tr>
<tr><td>Study intervention Assignment</td><td><ul><li>Participants with early, mild/moderate COVID-19 who are not on supplementary oxygen and at risk for disease progression will be randomised 1:1:1 to receive a single dose of VIR-7831 500mg IV, VIR-7831 500mg IM or VIR-7831 250mg IM.</li></ul>Participants will be stratified based on:<ol><li>Age: 12-17 years old, 18-64 years old, and ≥65 years old</li><li>COVID-19 Vaccination History: receipt of any COVID-19 vaccine (yes, no)</li><li>Region of the world (North America, South America, Europe, South Asia, Rest of Asia, Rest of the World).</li></ol><ul><li>All participants will be centrally randomized using an Interactive Web Response System (IWRS).</li><li>Participants will receive sotrovimab either by IM or IV directly from the investigator or designee, under medical supervision. The date and start and stop times of the dose administered will be recorded in the source documents. The dose of study intervention and study participant identification will be confirmed at the time of dosing by a member of the study site staff other than the person administering the study intervention.</li></ul></td></tr>
<tr><td>Interim Analysis</td><td><ul><li>The Joint Safety Review Team (JSRT) comprising individuals from Vir and GSK will review safety data at regular intervals throughout the conduct of the study. Details of the JSRT process is recorded in relevant SRT documents.</li><li>If, during its regular scheduled review of study data, the JSRT identifies a potential safety signal, Vir may request an ad hoc meeting of the IDMC to review data for further evaluation.</li><li>An interim analysis was planned (see Protocol Amendment Version 2) once 50% of the planned number of participants had completed the Day 29 assessment. However, prior to the time of the planned analysis, the JSRT noted a discrepancy in the rate of progression to hospitalization occurring in</li></ul></td></tr>
</table>

| Overview of Study Design and Key Features | |
|---|---|
| | the 250mg IM arm compared with the 500mg IM and IV arms. Upon review of the cumulative data, the JSRT made the decision to pause enrollment into the 250mg IM arm on 04-August-2021 and escalated the issue to the IDMC.<br>• An ad hoc meeting of the IDMC was called on 11-August-2021 and based upon their review of the data, the IDMC concurred that enrollment in the 250mg IM arm should be discontinued. As this ad hoc IDMC meeting occurred prior to the timepoint when the formal interim analysis would have occurred and concluded with discontinuing one arm of the study combined with an unexpected rapid increase in the rate of enrollment, it was not feasible to conduct the formal interim analysis.<br>• Therefore, data for the 250mg IM arm will be summarised only and not used for any statistical hypotheses tests or analyses. |

### 2. STATISTICAL HYPOTHESES

The primary objective of this study is to evaluate the efficacy of IM sotrovimab versus IV sotrovimab in preventing the progression of mild/moderate COVID-19 in a non-inferiority comparison.

The primary endpoint is progression of COVID-19 through Day 29 as defined by hospitalization > 24 hours for acute management of illness OR death.

Denoting the proportions of participants with progression of COVID-19 by Day 29 in the sotrovimab 500 mg IM arm, sotrovimab 250 mg IM arm and sotrovimab 500 mg IV arm as $P_{IM500}$, $P_{IM250}$ and $P_{IV500}$, respectively, then the null ($H_0$) and alternative ($H_A$) hypotheses are as follows using a non-inferiority margin of 3.5% (per feedback from the FDA, as mentioned in protocol section 4.2) on the risk difference scale:

(1) Sotrovimab 500 mg IM vs sotrovimab 500 mg IV:

- $H_0$: $P_{IM500} – P_{IV500} \geq 3.5\%$. $H_A$: $P_{IM500} – P_{IV500} < 3.5\%$

(2) Sotrovimab 250 mg IM vs sotrovimab 500 mg IV:

- $H_0$: $P_{IM250} – P_{IV500} \geq 3.5\%$. $H_A$: $P_{IM250} – P_{IV500} < 3.5\%$

#### 2.1. Multiplicity Adjustment

Hypotheses (1) and (2) above will be tested using the primary estimand according to the hierarchical testing principal as follows:

- If the null hypothesis for (1) is rejected, i.e., the upper bound of the two-sided 95% confidence interval for the risk difference between sotrovimab 500 mg IM and sotrovimab 500 mg IV is less than the pre-specified 3.5% non-inferiority margin, then hypothesis for (2) will be tested.

- If the null hypothesis for (1) is not rejected, then hypothesis for (2) will not be tested.

Below is a summary of possible outcomes of the hierarchical testing procedure:

| Which Null Hypothesis is Rejected | Non-inferiority Declared For |
|---|---|
| Hypotheses (1) and (2) | Both sotrovimab 500 mg IM and sotrovimab 250 mg IM (vs. sotrovimab 500 mg IV) |
| Hypothesis (1) but Not (2) | Only sotrovimab 500 mg IM vs. sotrovimab 500 mg IV |
| Hypothesis (1) is NOT rejected (Null hypothesis for (2) won't be tested in this case) | Neither sotrovimab 500mg IM nor sotrovimab 250mg IM (vs sotrovimab 500mg IV) |

Two secondary endpoints will be formally analysed:

1) Mean area under the curve of SARS-CoV-2 viral load in nasal secretions as measured by qRT-PCR from Day 1 to Day 8 ($AUC_{D1-8}$). Each IM dose will be assessed for equivalence to IV based on the two-sided 90% confidence interval for the treatment ratio falling within equivalence bounds of 0.5 to 2.0.
2) Progression of COVID-19 through Day 29 as defined by visit to a hospital emergency room for management of illness OR Hospitalization for acute management of illness for any duration and for any cause OR Death. Each IM dose will be assessed for non-inferiority vs IV using a non-inferiority margin of 3.5% on the risk difference scale.

The testing of the two secondary endpoints above is adjusted for multiplicity by using the following hierarchy:

- Secondary endpoint (1) will only be tested if non-inferiority is achieved for sotrovimab 500 mg IM vs. sotrovimab 500 mg IV and sotrovimab 250 mg IM vs. sotrovimab 500 mg IV comparisons for the primary endpoint.
- Secondary endpoint (1) will be tested in the same sequential way as the primary endpoint i.e If the null hypothesis for sotrovimab 500 mg IM vs. sotrovimab 500 mg IV is rejected, then the null hypothesis for sotrovimab 250 mg IM vs. sotrovimab 500 mg IV will be tested. If the null hypothesis for sotrovimab 250 mg IM vs sotrovimab 500mg IV for secondary endpoint (1) is rejected, then secondary endpoint (2) will be tested in the same sequential manner as secondary endpoint (1).

Given that recruitment to the sotrovimab 250 mg IM arm was permanently discontinued following the *ad hoc* interim analysis on 11-Aug-2021, the sotrovimab 250 mg IM arm will not be formally compared to the sotrovimab 500 mg IV arm. Therefore only hypotheses for primary and secondary endpoints related to the 500 mg IM arm will be formally tested in the above framework and hypotheses related to the 250 mg IM arm are removed from the testing hierarchy.

### 3. ANALYSIS SETS

| Analysis Set | Definition / Criteria | Analyses Evaluated |
|---|---|---|
| Screened | All participants who were screened for eligibility | Study Population |
| Enrolled | All participants who entered the study<br>Note screening failures (who never passed screening even if rescreened) are excluded from the Enrolled analysis set as they did not enter the study | Study Population |
| Randomized | All participants who were randomized. Participants will be analyzed according to the route of administration and dose level they were randomized to: sotrovimab IM (500 mg), sotrovimab IM (250 mg) or sotrovimab IV (500 mg). | Study Population |
| Intent-to-Treat (ITT) | All participants who were randomized. Participants who were randomized under Protocol Amendment Version 1 and were immunocompetent and fully vaccinated at randomisation will be excluded.<br>Participants will be analyzed according to the route of administration and dose level they were randomized to: sotrovimab IM (500 mg), sotrovimab IM (250 mg) or sotrovimab IV (500 mg). | Study Population<br>Efficacy |
| Safety | All randomized participants who are exposed to study intervention.<br>Participants will be analyzed according to the route of administration and dose level they actually received: sotrovimab IM (500 mg), sotrovimab IM (250 mg) or sotrovimab IV (500 mg). | Study Population<br>Safety |
| Pharmacokinetic (PK) | All participants in the Safety analysis set who had at least 1 non-missing PK assessment (Non-quantifiable [NQ] values will be considered as non-missing values).<br>Participants will be analyzed according to the route of administration and dose level they actually received. | PK |

| Analysis Set | Definition / Criteria | Analyses Evaluated |
|---|---|---|
| Virology | All participants in the ITT analysis set with a lab confirmed quantifiable baseline nasopharyngeal swab at Day 1.<br><br>Participants will be analyzed according to the route of administration and dose level they were randomized to: sotrovimab IM (500 mg), sotrovimab IM (250 mg) or sotrovimab IV (500 mg).<br><br>This will be the primary analysis set for virology. | Efficacy (Virology) |

### 4. STATISTICAL ANALYSES

#### 4.1. General Considerations

##### 4.1.1. General Methodology

The ITT Analysis Set will be used for all Study Population analyses and Efficacy analyses, unless otherwise specified  A subset of the ITT analysis set will be used for the primary estimand, excluding participants who did not meet key inclusion/exclusion criteria (see 4.2.2). The Safety Analysis Set will be used for selected Study Population summaries and all safety analyses.  The PK analysis Set will be used for all PK analyses. Virology analysis set will be used for all Virology analyses.

In the case of a difference between the stratification assigned at the time of randomization and the data collected in the eCRF, the analyses will be performed based on the data collected in the eCRF, not the assigned stratum at randomization. All analyses will be adjusted for treatment group (sotrovimab 500mg IV, sotrovimab 500mg IM), derived age group (<65 years old, ≥65 years old, see Section 6.2.7), and gender (female, male). Stratification factors of region and vaccination status are not included as covariates due to the small number of non-US and partially/fully vaccinated participants in the ITT analysis set. In addition the 12-17 years old and 18-64 years old categories are combined due to the small number of participants in the 12-17 years old category.

In the case of model convergence issues, if one category dominates within a particular covariate then that covariate may be considered for removal from the model.

Confidence intervals will use two-sided 95% confidence levels unless otherwise specified.

Unless otherwise specified, continuous data will be summarized using descriptive statistics: n, mean, standard deviation (std), median, minimum and maximum. For log-transformed data (e.g. AUC) descriptive statistics will present the geometric mean, standard deviation (SD of log-transformed data) and coefficient of variation. Categorical data will be summarized as the number and percentage of participants in each category.

Efficacy data will be reported according to nominal time of clinical visits unless otherwise stated. Hospitalization status, oxygen supplementation and healthcare resource utilization use will be slotted against the clinical visit date for summaries and analyses by visit.

All data will be reported according to the nominal time of clinic visits and assessments as specified in the protocol unless stated otherwise:

- Day 8, Day 11, Day 15, Day 22, where data within ±1 days of the target day may be used if data is not recorded on the actual day
- Day 29, where data within ±2 days of the target day may be used if data is not recorded on the actual day
- Day 57, where data within ±4 days of the target day may be used if data is not recorded on the actual day
- Day 85, Day 113, Day 141, Day 169, Day 197, Day 224, Day 252 where data within ±7 days of the target day may be used if data is not recorded on the actual day

It is anticipated that patient accrual will be spread thinly across centers and summaries of data by center would unlikely be informative and will not, therefore, be provided.

##### 4.1.2. Baseline Definition

For all endpoints the baseline value will be the latest pre-dose assessment with a non-missing value, including those from unscheduled visits and the screening visit. If time is not collected, Day 1 assessments are assumed to be taken prior to first dose and used as baseline. For endpoints assessed using an analysis set other than the Safety analysis set, the baseline value for subjects who are randomised and not dosed will be the latest assessment with nominal visit on or prior to Day 1.

Unless otherwise stated, if baseline data is missing no derivation will be performed and baseline will be set to missing.

##### 4.1.3. Missing Data Handling Rules

- Missing data can occur due to study withdrawal or participants lost to follow-up before the completion of the study or due to intermittent missing values (i.e. data between two non-missing assessments).
- In addition, for the hypothetical estimands of the primary and secondary COVID-19 progression endpoints, data collected after the occurrence of an intercurrent event will be excluded from the analysis and will therefore be set to missing.
- For the primary and secondary COVID-19 progression endpoints, missing data will be imputed under a missing at random (MAR) assumption using a multiple imputation (MI) model. More details are provided in Section 6.2.8.

- For all other endpoints, a treatment policy strategy will be used for all intercurrent events where all data captured will be included in the summary and analysis, unless otherwise stated.

The following rules will be applied to viral load endpoints:

- The imputation logic when a sample is reported as 'NEG' (below lower-limit of detection) or '<2.08' (below lower-limit of quantification) is to impute as 0.5 x 120 copies/ml = 60 copies/ml = 1.78 $\log_{10}$ copies/ml
  - Subjects who die prior to Day 8 (or Day 5 or Day 11), their AUC viral load value will be imputed using the worst AUC viral load value from other subjects in the same treatment arm who have completed Day 8 (5/11) and have a valid AUC(D1-8)/(D1-5)/(D1-11).
  - If Day 8 assessment alone is missing but Day 5 and Day 11 assessments are available, the Day 8 will be imputed using linear interpolation between Day 5 and Day 11. AUC will be calculated then.
  - If more than 2 consecutive assessments are missing prior to and including Day X, then the AUC(D1-X) will not be calculated.
  - If a subject is lost to follow-up prior to Day X, then AUC(D1-X) will be set to missing.

#### 4.2. Primary Endpoint Analyses

##### 4.2.1. Definition of Primary Endpoint

The primary endpoint is the progression of COVID-19 through Day 29 as defined by hospitalization > 24 hours for acute management of illness due to any cause OR death.

The primary efficacy endpoint will primarily be assessed using an estimand that uses a hypothetical strategy to deal with the main intercurrent events as detailed in section 1.1.

##### 4.2.2. Main analytical approach

The primary analysis will be conducted on the ITT analysis set, excluding participants who did not meet key inclusion/exclusion criteria (see section 4.2.2.1). All intercurrent events (see below) will be handled using a hypothetical strategy for the primary estimand, i.e. assuming the intercurrent event had not occurred.

The primary endpoint will be summarized using counts and percentages of the number of participants who have progression of COVID-19. The proportion of participants meeting the primary endpoint will be compared for sotrovimab 500 mg IM vs sotrovimab 500 mg IV using a 3.5% non-inferiority margin on the risk difference scale as per section 2.

The proportion of participants meeting the primary endpoint will be compared between treatments using a generalized linear model (GLM) from the binomial family with identity link, adjusted for treatment group (sotrovimab 500mg IV, sotrovimab 500mg IM), derived age group (<65 years old, ≥65 years old, see Section 6.2.7), and gender (female, male).

The absolute risk difference and associated 95% CI will be computed to test for non-inferiority in hypothesis (1).

Covariates may be removed from the model if it fails to converge as described in section 4.1.1. If the model still does not converge a GLM assuming a normal distribution for the endpoint and with identity link will be used, adjusted for the covariates above. If no events are observed on either treatment arm, the Farrington-Manning test will be used to compare arms.

The anticipated intercurrent events are:

- Not receiving randomized treatment or an alternative study treatment (i.e. treatment misallocation)
- Discontinuation of study intervention as described in Protocol Section 7.1.
- Use of medication not permitted during the study as listed in Protocol Section 6.8.1 up to day 29

Events are only deemed to be intercurrent events if they occur before occurrence of the primary endpoint. Data observed in the period following an intercurrent event (i.e. from the study day following the intercurrent event) will be excluded from the analysis. Data from this period will be assumed “missing at random” (MAR).

For participants that withdraw from the study and do not meet the primary endpoint at the time of withdrawal, data in the post-withdrawal period will also be assumed MAR.

Details of the method for multiple imputation is described in section 6.2.8.

###### 4.2.2.1. Key Inclusion/Exclusion Criteria

| Type | Category | Criteria |
|---|---|---|
| Inclusion | 5.1.2. Type of Participant and Disease Characteristics | 1. Participants who have a positive SARS-CoV-2 test result within 7 days of randomization (by any validated diagnostic test e.g. RT-PCR, antigen based testing on any specimen type) |
| | | 4. Participant to be dosed less than or equal to 7 days from onset of symptoms to dosing day (D1)<br>Note: Participants will only be excluded if ≥9 days since dosing. |
| Exclusion | 5.2.1. Medical Conditions | 1. Currently hospitalized or judged by the investigator as likely to require hospitalization in the next 24 hours<br>Note: Participants will only be excluded if hospitalized at baseline per eCRF form. |
| | 5.2.2. Prior/Concurrent Clinical Study Experience | 5. Enrollment in any investigational vaccine study within the last 180 days or any other investigational drug study within 30 days prior to Day 1 or within 5 half-lives of the investigational compound, whichever is longer |

| | | |
|---|---|---|
| | | 6. Enrollment in any trial of an investigational drug, vaccine or device study for SARS-CoV-2/COVID-19 within 90 days prior to Day 1 or within 5 half-lives of the investigational compound, whichever is longer |
| | 5.2.3. Other Exclusions | 7. Immunocompetent individuals who are fully vaccinated against SARS-CoV-2. A person is considered fully vaccinated if at least 14 days have passed since receiving the final dose in a COVID-19 vaccine series. |
| | | 8. Receipt of convalescent plasma from a recovered COVID-19 patient or anti-SARS-CoV-2 mAb within the last 3 months. |

##### 4.2.3. Additional estimands

A supplementary estimand will be conducted in the ITT analysis set by handling all intercurrent events with a treatment policy strategy (i.e. regardless of the intercurrent events occurring), with all the other attributes being the same as in the primary estimand. To estimate this estimand, all data collected in the population of interest up to Day 29 will be included in the analysis. Missing data due to participant withdrawal from study will be handled in the same manner as the primary estimand.

##### 4.2.4. Sensitivity analyses

To assess the sensitivity of the results to the missing data, a tipping point analyses will be performed for the hypothetical and treatment policy estimands. The underlying response rate among those subjects with missing response status in each arm will be varied ranging between 0 and 1. This analysis will be two-dimensional, i.e. it will allow for assumptions about the assumed response rate (and thus missing outcomes) in the two treatment arms to vary independently. For combinations of the assumed response rates in the arms, the number of additional responders among subjects with missing response will be imputed multiple times by drawing from a binomial distribution. The absolute difference and associated standard error for each imputed dataset will be calculated using the primary analysis model and results combined using Rubin's rules to calculate the risk difference and the 95% CI for the treatment comparison. Results will be presented via a heatmap.

In addition, a summary of the number of COVID-19 progressions through Day 29 (as defined in section 4.2.1) for the hypothetical estimand will be provided for the split by those progressions that occur <3 days and ≥3 days post-dose.

#### 4.3. Secondary Endpoints Analyses

The secondary endpoints will be based on the ITT Analysis Set, unless otherwise specified.

##### 4.3.1. Definition of Secondary Endpoints

###### 4.3.1.1. Safety and tolerability

Adverse events analyses including the analysis of adverse events (AEs), serious AEs (SAEs) and other significant AEs will be based on GSK Core Data Standards.

An overview summary of AEs, including counts and percentages of participants with

- any AE
- any SAE
- any AESI

will be provided as outlined in Section 4.5.2. The Safety analysis set will be used.

Since it will not be possible to delineate in a single participant whether the hospitalization is directly related to COVID-19 complications or could be related to VIR-7831 causing more severe disease due to ADE, all hospitalizations regardless of cause will be included in the primary endpoint. However, if a hospitalization or adverse event is related to expected progression, signs, or symptoms of COVID-19 (as detailed in Section 8.4.8 of the Protocol) or if hospitalization is due to elective treatment of a pre-existing condition that did not worsen from baseline as noted in Section 10.3 of the Protocol it will not be considered as an SAE. These events will be collected and reported as SAEs only if the event is more severe than expected for the participant's current clinical status and medical history or if the investigator feels that it is related to study drug.

In addition, a summary of the number and percentage of participants with disease-related events, that is, events related to expected progression, signs, or symptoms of COVID-19, unless more severe than expected for the participant's current clinical status and medical history will be provided. These events will be captured on the disease related event CRF. This data will also be listed.

The following are examples of events NOT meeting the AE definition but will be classed and captured separately as disease progression events in the CRF:

- hypoxemia due to COVID-19 requiring supplemental oxygen
- hypoxemia due to COVID-19 requiring non-invasive ventilation or high flow oxygen devices
- respiratory failure due to COVID-19 requiring invasive mechanical ventilation or ECMO

###### 4.3.1.2. Immunogenicity of VIR-7831

The incidence and titers (if applicable) of serum ADA to VIR-7831 will be listed and summarized for the Week 36 safety summary in the Safety analysis set.

Additional immunologic analyses may be conducted and will be documented in a separate technical document.

###### 4.3.1.3. Progression of COVID-19 by Day 29 (emergency room, hospitalisation for any duration or cause, death)

Progression of COVID-19 through Day 29 as defined by:

1. Visit to a hospital emergency room for management of illness

OR

2. Hospitalization for acute management of illness for any duration and for any cause

OR

3. Death

###### 4.3.1.4. Proportion of Participants Who Progress to Develop Severe or Critical Respiratory COVID-19 at Day 8, 15, 22 and 29

Participants are defined as progressing to develop severe respiratory COVID-19 if they require supplemental oxygen either by nasal cannula, face mask, high-flow oxygen devices, or non-invasive ventilation.

Participants are defined as progression to critical respiratory COVID-19 if they require invasive mechanical ventilation or ECMO.

Participants who die prior to the timepoint of interest without first having received supplemental oxygen will be considered to have met the endpoint (composite estimand strategy).

Further detail on the severity of respiratory COVID-19 based on the proportion of participants meeting each tier of the Vir modified version of the NIAID Ordinal Scale for Clinical Improvement (Table 1) will be summarized.

##### Table 1 Ordinal Scale for Clinical Improvement

| Ordinal Scale (Vir modified version, adapted from the Adaptive COVID-19 Treatment Trial [ACTT], 2020; referred to as National Institute of Allergy and Infectious Disease (NIAID) scale hereafter) |
|---|
| 1) Not hospitalized, no limitations of activities |
| 2) Not hospitalized, limitation of activities and/or requiring home oxygen [1] |
| 3) Hospitalized, not requiring supplemental oxygen and no longer requiring ongoing medical care (used if hospitalization was extended for infection-control reasons) |
| 4) Hospitalized, not requiring supplemental oxygen but requiring ongoing medical care (COVID-19–related or other medical conditions) |
| 5) Hospitalized, requiring any supplemental oxygen |
| 6) Hospitalized, requiring non-invasive ventilation or use of high-flow oxygen devices |
| 7) Hospitalized, receiving invasive mechanical ventilation or ECMO [2] |
| 8) Death |

[1] report as category 5 if using oxygen at home (unless were receiving home oxygen pre-morbidly), [2] Participant in receipt of extracorporeal membrane oxygenation (ECMO) at the time of dosing on Day 1 are excluded from the study (participants who subsequently require ECMO may continue in the study).

###### 4.3.1.5. Reduction in SARS-CoV-2 Viral Load

Viral load will be log10-transformed prior to summary and analysis. The Virology analysis set will be used for all analyses.

Reduction of SARS-CoV-2 viral shedding will be determined by the change from baseline of SARS-CoV-2 nasal viral load at Day 8.

In addition, the SARS-CoV-2 viral load will also be assessed from the area under the curve (AUC as determined by the trapezoidal rule) of $log_{10}$-transformed SARs-CoV-2 viral load through Days 1 to 8. AUC will be log-transformed prior to analysis.

The proportion of participants with a persistently high SARS-CoV-2 viral load (PHVL) assessed by qRT-PCR at Day 8 will also be assessed. PHVL is defined as a viral load value >4.1 $log_{10}$ copies/ml at Day 8.

###### 4.3.1.6. Pharmacokinetics of VIR-7831

Serum pharmacokinetic concentrations will be listed and summarized by treatment and visit in the PK analysis set. Any concentration not included in summaries should be flagged in the individual listings with an explanation for the exclusion.

Additional PK analyses, including additional exposure-response analyses, will be summarized in a separate PopPK analysis plan.

##### 4.3.2. Main Analytical Approach

###### 4.3.2.1. Binary Endpoints

The following secondary endpoints will be summarised/analysed as binary endpoints:

- Participants who have progression of COVID-19 (emergency room, hospitalisation for any duration or cause, death) through Day 29
- Proportion of Participants Who Progress to Develop Severe or Critical Respiratory COVID-19 at Day 8, 15, 22 and 29
- Proportion of participants with a persistently high SARS-CoV-2 viral load at Day 8 by qRT-PCR

The secondary binary endpoints will be summarized using counts and percentages.

Participants who have progression of COVID-19 (emergency room, hospitalisation for any duration or cause, death) through Day 29 will be analysed as per the primary endpoint (hypothetical and treatment policy estimands).

###### 4.3.2.2. Continuous Endpoints

The following secondary endpoints will be summarised/analysed as continuous endpoints:

- Change from baseline in viral load in nasal secretions by qRT-PCR at Day 8
- Area under the curve (AUC) of SARS-CoV-2 viral load as measured by qRT-PCR from Day 1 to Day 8 ($AUC_{D1-8}$)
- Serum pharmacokinetic concentrations

Change from baseline in viral load at all visits including Day 8 will be summarised on the log10 scale. The Virology analysis set will be used.

$AUC_{D1-8}$ for log10-transformed SARS-CoV-2 viral load will be analyzed using analysis of covariance adjusted for treatment group (sotrovimab 500mg IV, sotrovimab 500mg IM), baseline viral load, derived age group (<65 years old, ≥65 years old, see Section 6.2.7), and gender (female, male). AUC will be log-transformed prior to analysis. The treatment ratio and two-sided 90% CI for the comparison of sotrovimab 500mg IM vs sotrovimab 500mg IV will be calculated by back-transforming the corresponding estimates on the log-scale.

In addition, the following will be produced:

- Box plot of serum pharmacokinetic concentrations at Day 8, 15 and 29 vs COVID-19 progression status through Day 29 (as defined in section 4.2.1) for the hypothetical estimand, overall and by treatment group

- Summary table of Day 8 serum pharmacokinetic concentrations (n, mean, SD, median, min, max) by COVID-19 progression status through Day 29 (as defined in section 4.2.1) for the hypothetical estimand, total and by treatment group.

#### 4.4. Exploratory Endpoints Analyses

The exploratory analyses will be based on the ITT Analysis Set, unless otherwise specified.

##### 4.4.1. Definition of Exploratory Endpoints

###### 4.4.1.1. Incidence of Hospitalisation through Day 29

A participant is defined as requiring hospital stay if they received any in-patient healthcare (A&E, general ward or ICU) encounter of any duration.

Data will be summarised as collected (while-alive estimand strategy) in addition to assuming participants who die prior to Day 29 to have met the endpoint (composite estimand strategy).

###### 4.4.1.2. Total Hospital Length of Stay

A participant is defined as requiring hospital stay if they received any in-patient healthcare (A&E, general ward or ICU) encounter of any duration. A participant may have more than one period of hospital stay included in the total hospital length of stay.

Data will be summarised as collected (while-alive estimand strategy) in addition to assuming participants who die prior to Day 29 to have been in hospital from the date of death to Day 29 (composite estimand strategy).

###### 4.4.1.3. Proportion of subjects requiring ICU stay or mechanical ventilation through Day 29

A participant is defined as being on a ventilator if they received invasive mechanical ventilation or ECMO as a form of oxygen therapy. A participant is defined as requiring intensive care if they received an intensive care unit in-patient healthcare encounter of any duration.

Data will be summarised as collected (while-alive estimand strategy) in addition to assuming participants who die prior to Day 29 to have met the endpoint (composite estimand strategy).

###### 4.4.1.4. Total Intensive Care Length of Stay

A participant is defined as requiring intensive care if they received an intensive care unit in-patient healthcare encounter of any duration. A participant may have more than one period of intensive care stays included in the total intensive care length of stay.

Data will be summarised as collected (treatment policy estimand strategy) in addition to assuming participants who die prior to Day 29 to have been in intensive care from the date of death to Day 29 (composite estimand strategy).

###### 4.4.1.5. SARS-CoV-2 Resistant Mutants Against VIR-7831 at Baseline

Summaries of viral resistance mutations at baseline will be provided based on frequency counts in the Safety analysis set.

###### 4.4.1.6. Emergence of SARS-CoV-2 Resistant Mutants Against VIR-7831

Summaries of viral resistance mutations will be provided based on frequency counts in the Safety analysis set.

###### 4.4.1.7. Reduction in SARS-CoV-2 Viral Load

Viral load will be log10-transformed prior to summary and analysis. The Virology analysis set will be used for all analyses.

Reduction of SARS-CoV-2 viral shedding will be determined by the change from baseline of SARS-CoV-2 nasal viral load at Day 3, Day 5, Day 11, Day 15, Day 22, and Day 29.

In addition, the SARS-CoV-2 viral load will also be assessed from the area under the curve (AUC as determined by the trapezoidal rule) of $log_{10}$-transformed SARs-CoV-2 viral load through Days 1 to 5 ($AUC_{D1-5}$) and Days1 to 11 ($AUC_{D1-11}$). AUC will be log-transformed prior to analysis.

The proportion of participants with undetectable, unquantifiable and positive SARS-CoV-2 in nasal secretions by qRT-PCR at Day 3, Day 5, Day 8, Day 11, Day 15, Day 22, and Day 29 will be summarised.

In addition a Kaplan-Meier plot of time to undetectable SARS-CoV-2 in nasal secretions by qRT-PCR by treatment group will be produced. For the purposes on the plot, a confirmed negative PCR is defined as first of two or more consecutive negative (no SARS-CoV2 detected) PCR tests.

###### 4.4.1.8. SARS-CoV-2 viral load measured by qRT-PCR

Summaries will be provided to compare different sample collection methods in SARS-CoV-2 viral load (e.g. nasopharyngeal swab, saliva) within participants. The Virology analysis set will be used for all analyses.

###### 4.4.1.9. SARS-CoV-2 anti-N antibody at Day 1 and Day 29

The incidence and titers (if applicable) of anti-N SARS CoV-2 antibodies at Day 1 and Day 29 will be summarized using frequency counts by treatment (including a total column) in the Safety analysis set.

###### 4.4.1.10. Incidence and severity of Long COVID symptoms at Week 12, Week 24 and Week 36

To monitor for subsequent development of long COVID at Week 12, Week 24 and Week 36, participants will be asked a series of questions to evaluate a range of symptoms after first being infected with the SARS-CoV-2. The questions to be asked are defined in Appendix 6 of the Protocol.

The participant responses to the questions will be summarised, based on ITT analysis set. The frequency of the response Grade categories (“*Grade 1”* to “*Grade 3*”) will be presented for each question.

##### 4.4.2. Main Analytical Approach

All exploratory endpoints will be summarised only.

Total hospital length and total intensive care length of stay will only be summarised if at least 10% of participants have one or more recorded event of interest. Length of stay will be summarised in all patients in the analysis set, in addition to those with the event of interest. Otherwise the data will be listed only.

#### 4.5. Safety Analyses

The safety analyses will be based on the Safety Analysis Set, unless otherwise specified.

##### 4.5.1. Extent of Exposure

Subjects will receive either an IV infusion of sotrovimab or a single IM injection of sotrovimab at one of two dose levels.

The intravenous sotrovimab dose will be 500 mg and will be infused over 15 minutes. The intramuscular sotrovimab dose will be either 250 mg or 500 mg given at a single timepoint. The 250 mg IM dose will be given either as a single 250 mg (4ml) injection in the dorsogluteal muscle or as two 2 ml injections in each deltoid muscle. The 500 mg IM dose will be administered as two 4 ml injections in each dorsogluteal muscle.

Summaries of exposure will be limited to the number of participants exposed and the number of participants with interruptions or infusions/injections stopped early and not completed.

##### 4.5.2. Adverse Events

Adverse events analyses including the analysis of adverse events (AEs), Serious AEs (SAEs) and other significant AEs will be based on GSK Core Data Standards. As per the protocol, AEs are collected through Week 12 and SAEs collected through Week 36.

An overview summary of AEs will be produced, including counts and percentages of participants with:

- AEs
- AEs related to study intervention
- AEs leading to permanent discontinuation of study intervention,
- AEs leading to temporary interruption of study intervention
- Grade 3 and 4 AEs
- Grade 3 and 4 AEs related to study intervention,
- Grade 3 and 4 AEs leading to permanent discontinuation of study intervention,
- Grade 3 and 4 AEs leading to temporary interruption of study intervention
- SAEs
- SAEs related to study intervention,
- Fatal SAEs
- Fatal SAEs related to study intervention.
- Local infusion/injection site reactions

Adverse events will be coded using the latest version of the standard Medical Dictionary for Regulatory Affairs (MedDRA dictionary) and graded by the investigator according to the DAIDS 2017 v2.1.

The following adverse events will be considered of special interest (AESI) and will be summarised using frequency and percentages separately.

- Systemic infusion/injection related reactions (IRR) including hypersensitivity reactions (HSR); reactions within 24 hours of start of infusion will be identified using a list of MedDRA preferred terms confirmed by the Safety team.
- Local infusion/injection site reactions (ISR) will be identified using a list of MedDRA preferred terms confirmed by the Safety team.
- Immunogenicity (Anti-Drug Antibodies (ADA)) related adverse drug reactions will be reported. These potential events of ADAs will be detected by reviewing AEs that indicate HSR in those subjects who have positive anti-drug antibodies.
- Adverse events potentially related to antibody-dependent enhancement of disease (ADE) will be reported. These potential events of ADEs will be detected by reviewing adverse event/serious adverse events for an increase in the incidence and severity of COVID complications for a participant that cannot be explained by underlying risk factors.

In addition to the AE overview summary,

- Most common AEs defined as any AE preferred term (PT) with an incidence of at least ≥1% (approx. 2 subjects) in any of the treatment group will be summarised using frequency and percentages separately.
- AEs by severity will be presented. The summary will use the following algorithms for counting the participant:
  - Preferred term row: Participants experiencing the same AE preferred term several times with different grades will only be counted once with the maximum grade.
  - Any event row: Each participant with at least one adverse event will be counted only once at the maximum grade no matter how many events they have.

The frequency and percentage of AEs (all grades) will be summarized and displayed in descending order by system organ class (SOC) and PT.

In the SOC row, the number of participants with multiple events under the same SOC will be counted once.

A separate summary will be provided for study intervention-related AEs. A study intervention-related AE is defined as an AE for which the investigator classifies the possible relationship to study intervention as “Yes”.

A worst-case scenario approach will be taken to handle missing relatedness data, i.e. the summary table will include events with the relationship to study intervention as ‘Yes’ or missing. The summary table will be displayed by PT only.

All SAEs will be tabulated based on the number and percentage of participants who experienced the event. Separate listings will also be provided for study intervention-related SAEs.

A study intervention-related SAE is defined as an SAE for which the investigator classifies the relationship to study intervention as “Yes”. A worst-case scenario approach will be taken to handle missing data, i.e. the summary table will include events with the relationship to study intervention as ‘Yes’ or missing.

###### 4.5.2.1. Disease-Related Events (not classified as AEs)

Disease-related events will be summarised by treatment group by overall frequency.

##### 4.5.3. Laboratory Data

Summaries by shift tables of worst-case grade increase from baseline grade will be provided for all the lab tests that are gradable by DAIDS 2017 v2.1. These summaries will display the number and percentage of participants with a maximum post-baseline grade increasing from their baseline grade. Any increase in grade from baseline will be summarized along with any increase to a maximum grade of 2, any increase to a maximum grade of 3 and any increase to a maximum grade of 4. Missing baseline grade will be assumed as grade 0.

For laboratory tests with both low and high graded values, summaries will be provided separately and labelled by direction, e.g., sodium will be summarized as hyponatremia and hypernatremia separately.

Any clinically abnormal laboratory results which are not identified as AEs will be summarised separately, identification of such values will be described in the OPS document.

For lab tests that are not gradable by DAIDS 2017 v2.1, summaries of worst-case changes from baseline with respect to normal range will be generated. Decreases to low, changes to normal or no changes from baseline, and increases to high will be summarized for the worst-case post-baseline. If a participant has a decrease to low and an increase to high during the same time interval, then the participant is counted in both the “Decrease to Low” categories and the “Increase to High” categories.

Summaries and listings for hematology, and chemistry laboratory tests will be produced separately. Liver function laboratory tests will be included with chemistry lab tests.

Separate summary of hepatobiliary laboratory events (if any) including possible Hy’s law cases will be provided in addition to what has been described above.

Possible Hy’s law cases are defined as elevated alanine aminotransferase (ALT) > 3×upper limit of normal (ULN), total bilirubin ≥ 2×ULN or international normalized ratio (INR) >1.5. Total bilirubin ≥ 2×ULN can be within 29 days following the ALT elevation and if direct bilirubin is available on the same day, it must be ≥ 35% of total bilirubin.

The summary will be produced for worst case post baseline only. A plot for maximum post-baseline Total Bilirubin against ALT will also be produced.

##### 4.5.4. Vital Signs

Summaries of grade increase in systolic blood pressure (SBP) and diastolic blood pressure (DBP) will be provided separately. These summaries will display the number and percentage of participants with any grade increase, increase to Grade 2 and increase to Grade 3 for worst case post-baseline only. The grade definition for SBP is: Grade 0 (<120), Grade 1 (120-139), Grade 2 (140-159), Grade 3 (>=160). The grade definition for DBP is: Grade 0 (<80), Grade 1 (80-89), Grade 2 (90-99), Grade 3 (>=100). The summaries will be produced for worst case post baseline only.

In addition, summaries of respiratory rate, heart rate and temperature will be provided.

A boxplot will be presented for blood pressures by treatment. A summary of temperature will also be presented by planned visit/timepoints and pre-defined categories i.e., <38°C, 38°C-<38.4°C, 38.4°C-<38.9°C, 38.9°C-<40°C and ≥40°C.

In addition, listings of vital signs parameters (blood pressure, respiratory rate, heart rate and temperature) by visit will be provided.

##### 4.5.5. Oxygen Saturation

Summaries of actual and changes in blood oxygen saturation ($SpO_2$) will be provided in addition a listing of participants administered oxygen.

Requirement for respiratory support is assessed according to the following categories:

1. Room air
2. Low flow nasal canulae/face mask
3. Non-re-breather mask or high flow nasal cannulae/ non-invasive ventilation (including continuous positive airway pressure support)
4. Mechanical ventilation / extra-corporeal membrane oxygenation
5. Other
6. Death

Changes in requirement for respiratory support (excluding room air) will be summarized using proportions of participants with change from baseline to higher respiratory support in each category by study day using a stacked bar chart.

#### 4.6. Other Analyses

##### 4.6.1. Subgroup analyses

The progression of COVID-19 through Day 29 will be summarised using counts and proportions for the following subgroups:

- Age (<65, ≥65 years)
- BMI (<30, ≥30kg/m$^2$)
- Duration of symptoms (≤5, >5 days)
- Sex (Female, Male)

Change from baseline in viral load will be summarised by visit in the following subgroups:

- Age (<65, ≥65 years)
- Baseline viral load (<4, ≥4-<5, ≥5-<6, ≥6-<7, ≥7 $\log_{10}$ copies/mL)
- Duration of symptoms (≤5, >5 days)
- Serological status (Positive, Negative)

#### 4.7. Interim Analyses

The Joint Safety Review Team (JSRT) comprising individuals from Vir and GSK will review safety data at regular intervals throughout the conduct of the study. Details of the JSRT process is recorded in relevant SRT documents.

A single interim analysis (IA) was initially planned to be performed for evaluation of safety, efficacy and futility when approximately 50% of participants are enrolled and reach Day 29. The IA data was to be evaluated by an IDMC. However, prior to the time of the planned analysis, the JSRT noted a discrepancy in the rate of progression to hospitalization occurring in the 250mg IM arm compared with the 500mg IM and IV arms. Upon review of the cumulative data, the JSRT made the decision to pause enrollment into the 250mg IM arm on 04-August-2021 and escalated the issue to the IDMC. An ad hoc meeting of the IDMC was called on 11-August-2021 and based upon their review of the data, the IDMC concurred that enrollment in the 250mg IM arm should be discontinued. As this ad hoc IDMC meeting occurred prior to the timepoint when the formal interim analysis would have occurred and concluded with discontinuing one arm of the study combined with an unexpected rapid increase in the rate of enrollment, it was not feasible to conduct the formal interim analysis.

There will be 2 reporting efforts: the primary Day 29 efficacy and safety analyses, and the 36-Week Safety Follow-Up analyses.

#### 4.8. Changes to Protocol Defined Analyses

Changes from the originally planned statistical analysis specified in the protocol are detailed in Table 2.

**Table 2 Changes to Protocol Defined Analysis Plan**

| Protocol Defined Analysis | SAP Defined Analysis | Rationale for Changes |
|---|---|---|
| • Section 9.4.2: Planned primary endpoint model adjusted for treatment group, COVID-19 vaccination status, age (12-18, 19-64, ≥65 years old), region and gender. | • Planned primary endpoint model adjusted for treatment group, age (<65, ≥65 years old) and gender only. | • Model covariates amended due to small numbers of patients in non-US regions, small proportion of fully/partially vaccinated participants, and small number of participants <18 years. |

### 5. SAMPLE SIZE DETERMINATION

Sotrovimab IM participants and sotrovimab IV participants were planned to be enrolled and randomized in a 1:1:1 ratio to receive a single dose IV infusion of sotrovimab or a single IM injection of sotrovimab at one of two dose levels, in an open-label manner.

Approximately 1020 (340 per arm) participants were planned to be randomly assigned to study intervention. This sample size will provide approximately 90% power to demonstrate that IM injection of sotrovimab is non-inferior to IV infusion of sotrovimab in terms of the proportion of participants with progression of COVID-19 through Day 29 at the one-sided 2.5% significance level assuming COVID-19 progression rates of 2% in the sotrovimab IM and IV arms and 3.5% non-inferiority margin on the risk difference scale.

Following the discontinuation of sotrovimab 250 mg IM arm (See Section 4.7), no more participants were enrolled and randomized into this arm. The remaining participants were randomized in a 1:1 ratio to receive sotrovimab 500mg IV or sotrovimab 500 mg IM

**Sample Size/Power Sensitivity**

The table below shows power for showing non-inferiority for different true progression rates on the sotrovimab IV arm and when the progression rate in the sotrovimab 500mg IM arm is equal to or higher than that on IV, given a sample size of N=340 per arm.

| | | Progression rate in sotrovimab IV arm | | |
|---|---|---|---|---|
| | | 1.0% | 1.5% | 2.0% |
| Progression rate in sotrovimab IM arm minus Progression rate in sotrovimab IV arm | 0% (equal) | >99% | 96% | 90% |
| | +0.1% | 99% | 95% | 88% |
| | +0.2% | 98% | 93% | 85% |
| | +0.3% | 97% | 91% | 82% |

### 6. SUPPORTING DOCUMENTATION

#### 6.1. Appendix 1 Study Population Analyses

The study population analyses will be based on the Intent-to-Treat Analysis Set, unless otherwise specified. Please see Section 3 of the SAP for more information on analysis sets.

##### 6.1.1. Participant Disposition

A summary of subject status and subject disposition for the study conclusion record will be provided. This display will show the number and percentage of subjects who completed the study and who withdrew from the study, including primary and secondary reasons for study withdrawal and presented in the order they are displayed on the collection form. A subject is considered to have completed the study if he/she has completed the Week 36 visit. The end of the study is defined as the date of the last contact of the last subject in the study.

A summary of the number and percentage of subjects who passed screening and entered the study or who failed screening and therefore were not entered into the study, will be summarized along with the reasons for failure will be summarized for those subjects who failed screening. This summary will be produced based on the Screened analysis set.

The number of subjects will be summarized by Country, Site Id. and Investigator Id. This summary will be produced based on the Enrolled analysis set

A summary of the duration in days in follow-up since infusion/injection will be summarized categorically for the Safety analysis set. The duration is calculated using the following formula:

min (Date of study withdrawal, Date of Data Cut, Date of Study Completion) – Date of Dosing +1

where the earlier date of either a subject withdrawal from the study, the current data cut date, or the date a subject completed the study are used. The summary will display duration overall and by mortality status: alive or deceased. The study withdrawal date for deceased subjects coincides with their death date. Duration categories post-dose include:

- Less than 5 days
- 5 to 10 days
- 11 to 14 days
- 15 to 29 days
- Greater than 29 days
- Greater than 85 days
- Greater than 141 days
- Greater than 169 days
- Greater than 224 days

A summary of treatment exposure will display the number of subjects exposed, the duration of treatment administration, and the number of subjects with infusion/injection interrupted or stopped early and not completed using the Safety set.

A summary of the number of subjects in each of the analysis sets described in the SAP Section 3 will be provided. Also, a listing will display subject exclusions from any analysis set using the Screened analysis set.

##### 6.1.2. Demographic and Baseline Characteristics

The demographic characteristics (e.g., age including age categories, race, ethnicity, sex, baseline height, baseline BMI and its categories) will be summarized by descriptive statistics. An additional summary of age ranges using the EMA clinical trial results disclosure requirement categories will be produced and is based on the Enrolled analysis set (see SAP section 3).

The high-level FDA race categories and detailed race subcategories collected on the CRF will be summarized along with categories for mixed race based on the Intent-to-treat analysis set.

Number of positive SARS-CoV-2 results, specimen type used for SARS-CoV-2 test, diagnostic method, risk factors for COVID-19 progression (as listed in the Protocol Inclusion Criteria), number of COVID-19 conditions met, types of symptoms present, and symptom duration will be summarized.

Additionally, the Baseline Disease Characteristics table will summarize subjects with a baseline obesity risk factor as (BMI ≥30kg/$m^2$ for adult participants and BMI ≥85th percentile for age/gender based on CDC growth charts for adolescents) and summarize COPD risk factor to include the following protocol-defined conditions: history of chronic bronchitis, chronic obstructive lung disease, or emphysema with dyspnea on physical exertion.

The summary of stratification factors will be provided:

- Age: 12-17 years old, 18-64 years old, and ≥65 years old.
- COVID-19 Vaccination History: receipt of any COVID-19 vaccine.
- Region of the world (Europe, North America, South America, South Asia, Rest of Asia, Rest of the World).

Medical conditions collected at screening will be summarized separately by past and current. These factors will be summarized as collected for current medical conditions at screening.

##### 6.1.3. Protocol Deviations

Important protocol deviations are a subset of protocol deviations that may significantly impact the completeness, accuracy, and/or reliability of the study data or that may significantly affect a subject's rights, safety, or well-being. For example, important protocol deviations may include enrolling subjects in violation of key eligibility criteria designed to ensure a specific subject population or failing to collect data necessary to interpret primary endpoints, as this may compromise the scientific value of the trial.

Important protocol deviations (including deviations related to study inclusion/exclusion criteria, conduct of the trial, patient management or patient assessment) will be summarized. Protocol deviations will be tracked by the study team throughout the conduct of the study. These protocol deviations will be reviewed to identify those considered as important.

Data will be reviewed prior to unblinding and freezing the final Day 29 efficacy database to ensure all important deviations are captured and categorized in the protocol deviations SDTM dataset.

##### 6.1.4. Prior and Concomitant Medications

Concomitant medications will be coded using both the GSK Drug and WHO Drug coding dictionaries. However, they will only be summarized using the GSK Drug dictionary. The summary of concomitant medications will show the number and percentage of subjects taking concomitant medications by Anatomical Therapeutic Chemical (ATC) Level 1 (Body System) and by Ingredient. Standard of care concomitant medications taken for COVID-19 will also be summarized. Multi-ingredient products will be summarized by their separate ingredients rather than as a combination of ingredients. Concomitant Medications will be summarized and listed, while prior medications will be listed.

#### 6.2. Appendix 2 Data Derivations Rule

##### 6.2.1. Criteria for Potential Clinical Importance

This study will not be utilizing lab parameter ranges defining potential clinical important values.

Instead, summaries of worst-case grade increase from baseline grade will be provided for all the lab tests that are gradable by DAIDS 2017 v2.1, if available.

##### 6.2.2. Laboratory Values with DAIDS 2017 v 2.1 Grading

| Laboratory parameters of interest | Grade | | | |
|---|---|---|---|---|
| | 1<br>Mild | 2<br>Moderate | 3<br>Severe | 4<br>Potentially life-threatening |
| Absolute Neutrophil Count (ANC),<br>Low (cells/mm3; cells/L)<br><br>> 7 days of age | 800 to 1,000<br><br>$0.800 \times 10^9$ to $1.000 \times 10^9$ | 600 to 799<br><br>$0.600 \times 10^9$ to $0.799 \times 10^9$ | 400 to 599<br><br>$0.400 \times 10^9$ to $0.599 \times 10^9$ | < 400<br><br>$< 0.400 \times 10^9$ |
| Absolute Lymphocyte Count,<br>Low (cell/mm3; cells/L)<br><br>> 5 years of age (not HIV infected) | 600 to < 650<br><br>$0.600 \times 10^9$ to $< 0.650 \times 10^9$ | 500 to < 600<br><br>$0.500 \times 10^9$ to $< 0.600 \times 10^9$ | 350 to < 500<br><br>$0.350 \times 10^9$ to $< 0.500 \times 10^9$ | < 350<br><br>$< 0.350 \times 10^9$ |
| WBC, Decreased (cells/mm3; cells/L)<br><br>> 7 days of age | 2,000 to 2,499<br><br>$2.000 \times 10^9$ to $2.499 \times 10^9$ | 1,500 to 1,999<br><br>$1.500 \times 10^9$ to $1.999 \times 10^9$ | 1,000 to 1,499<br><br>$1.000 \times 10^9$ to $1.499 \times 10^9$ | < 1,000<br><br>$< 1.000 \times 10^9$ |

| | | | | |
|---|---|---|---|---|
| Hemoglobin16, Low (g/dL; mmol/L)<br><br>≥ 13 years of age (male only) | 10.0 to 10.9<br><br>6.19 to 6.76 | 9.0 to < 10.0<br><br>5.57 to < 6.19 | 7.0 to < 9.0<br><br>4.34 to < 5.57 | < 7.0<br><br>< 4.34 |
| Hemoglobin16, Low (g/dL; mmol/L)<br><br>≥ 13 years of age (female only) | 9.5 to 10.4<br><br>5.88 to 6.48 | 8.5 to < 9.5<br><br>5.25 to < 5.88 | 6.5 to < 8.5<br><br>4.03 to < 5.25 | < 6.5<br><br><4.03 |
| Platelets, Decreased (cells/mm3; cells/L) | 100,000 to < 125,000<br><br>100.000 x $10^9$ to < 125.000 x $10^9$ | 50,000 to < 100,000<br><br>50.000 x $10^9$ to < 100.000 x $10^9$ | 25,000 to < 50,000<br><br>25.000 x $10^9$ to < 50.000 x $10^9$ | < 25,000<br><br><25,000 x$10^9$ |
| INR, High<br><br>(not on anticoagulation therapy) | 1.1 to < 1.5 x ULN | 1.5 to < 2.0 x ULN | 2.0 to < 3.0 x ULN | ≥ 3.0 x ULN |
| ALT or SGPT, High<br><br>Report only one | 1.25 to < 2.5 x ULN | 2.5 to < 5.0 x ULN | 5.0 to < 10.0 x ULN | ≥ 10.0 x ULN |
| AST or SGOT, High<br><br>Report only one | 1.25 to < 2.5 x ULN | 2.5 to < 5.0 x ULN | 5.0 to < 10.0 x ULN | ≥ 10.0 x ULN |
| Total Bilirubin, High<br><br>> 28 days of age | 1.1 to < 1.6 x ULN | 1.6 to < 2.6 x ULN | 2.6 to < 5.0 x ULN | ≥ 5.0 x ULN |
| Potassium, High (mEq/L; mmol/L) | 5.6 to < 6.0<br>5.6 to < 6.0 | 6.0 to < 6.5<br>6.0 to < 6.5 | 6.5 to < 7.0<br>6.5 to < 7.0 | ≥ 7.0<br>≥ 7.0 |
| Potassium, Low (mEq/L; mmol/L) | 3.0 to < 3.4<br>3.0 to < 3.4 | 2.5 to < 3.0<br>2.5 to < 3.0 | 2.0 to < 2.5<br>2.0 to < 2.5 | < 2.0<br>< 2.0 |
| Creatinine, High<br><br>*Report only one | 1.1 to 1.3 x ULN | > 1.3 to 1.8 x ULN<br>OR<br>Increase to 1.3 to < 1.5 x subject's baseline | > 1.8 to < 3.5 x ULN<br>OR<br>Increase to 1.5 to < 2.0 x subject's baseline | ≥ 3.5 x ULN<br>OR<br>Increase ≥ 2.0 x subject's baseline |
| Creatinine Clearance or eGFR, Low<br><br>*Report only one | NA | < 90 to 60 ml/min or ml/min/1.73 m2 OR 10 to < 30% decrease from subject's baseline | < 60 to 30 ml/min or ml/min/1.73 m2 OR 30 to < 50% decrease from subject's baseline | < 30 ml/min or ml/min/1.73 m2 OR ≥ 50% decrease from subject's baseline or dialysis needed |

| | | | | |
|---|---|---|---|---|
| Cardiac Troponin I, High | NA | NA | NA | Levels consistent with myocardial infarction or unstable angina as defined by the local laboratory |

##### 6.2.3. Study Period

**Study phases for Concomitant Medication**

| Study Phase | Definition |
|---|---|
| Prior | If medication end date is not missing and is prior treatment start, or if end date is missing but the medication end reference equals BEFORE. |
| Concomitant | Any medication that is not a prior |

**Study Treatment Emergent Flag for Adverse Event**

| Flag | Definition |
|---|---|
| Study Treatment Emergent | If AE onset date is on or after study treatment start date:<br>• Study Treatment Start Date ≤ AE Start Date |

**NOTES:**

- Time of study treatment dosing and start/stop time of AEs should be considered, if collected.
- All Adverse Events tables and figures will be presented for Treatment Emergent AEs. AE listings will display all data.

##### 6.2.4. Study Day and Reference Dates

The safety reference date is the study treatment start date and will be used to calculate study day for all safety measures.

For efficacy, measurements of progressions including: all Hospitalizations, Oxygen Supplementation, and Death will measure from treatment start date to align with safety measurements. Efficacy measurements of all-cause mortality and incidence & duration of ICU, Hospitalization, and Ventilation will calculate study day from date of randomization.

- Ref Date = Missing → Study Day = Missing
- Ref Date < First Dose Date → Study Day = Ref Date – First Dose Date
- Ref Data ≥ First Dose Date → Study Day = Ref Date – (First Dose Date) + 1

##### 6.2.5. Multiple measurements at One Analysis Time Point

Where duplicate records exist per scheduled visit/time point/participant (if applicable) in vital signs (including oxygen saturation), laboratory data, and ECG measurements the latest record will be used for summaries.

Subjects having both High and Low values for DAIDS 2017 v2.1 or Normal Ranges at any post-baseline visit for safety parameters will be counted in both the High and Low categories of "Any visit post-baseline" row of related summary tables. This will also be applicable to relevant Potential Clinical Importance (PCI) summary tables where instead of PCI ranges DAIDS 2017 v2.1 or Normal ranges will be considered for reporting of clinically important safety findings.

All data from scheduled and unscheduled visits will be reported in the listings.

##### 6.2.6. Handling of Missing Data

| Element | Reporting Detail |
|---|---|
| General | • Missing data occurs when any requested data is not provided, leading to blank fields on the collection instrument:<br>o These data will be indicated using a "blank" in subject listing displays. Unless all data for a specific visit are missing in which case the data is excluded from the listing.<br>o Answers such as "Not applicable", "Not evaluable" and "Not Done" are not considered to be missing data and should be displayed as such. |
| Outliers | • Any subjects excluded from the summaries and/or statistical analyses will be documented along with the reason for exclusion in the clinical study report. |
| Efficacy Endpoints | Details around the missing data handling for efficacy endpoints are described in section 6.2.8. In addition:<br>• For the secondary binary endpoint of proportion of subjects who have progression of COVID-19 (i.e., hospitalisation (any duration) OR emergency room visit OR death) through Day 29, missing data will be handled as for the primary endpoint. |
| Safety Endpoints | • No missing data imputation will be performed for safety endpoints.<br>• Data will be reported as captured. |

##### 6.2.7. Handling of Missing and Partial Dates

| Element | Reporting Detail |
|---|---|
| General | • Partial dates will be displayed as captured in subject listing displays.<br>• However, where necessary, display macros may impute dates as temporary variables for sorting data in listings only. In addition, partial dates may be imputed for 'slotting' data to study phases or for specific analysis purposes as outlined below.<br>• Imputed partial dates will not be used to derive study day, time to onset or duration (e.g., time to onset or duration of adverse events), or elapsed time variables (e.g., time since diagnosis). In addition, imputed dates are not used for deriving the last contact date in overall survival analysis dataset. |

<table>
<tr><th>Element</th><th>Reporting Detail</th></tr>
<tr><td>Adverse Events</td><td>
<ul><li>Partial dates for AE recorded in the CRF will be imputed using the following conventions:</li></ul>
<table>
<tr><td>Missing start day</td><td><ul><li>If study treatment start date is missing (i.e. subject did not start study treatment), then set start date = 1st of month.</li><li>Else if study treatment start date is not missing:<ul><li>If month and year of start date = month and year of study treatment start date, then<ul><li>If stop date contains a full date and stop date is earlier than study treatment start date, then set start date= 1st of month.</li><li>Else set start date = study treatment start date.</li></ul></li></ul></li><li>Else set start date = 1st of month.</li></ul></td></tr>
<tr><td>Missing start day and month</td><td><ul><li>If study treatment start date is missing (i.e. subject did not start study treatment), then set start date = January 1.</li><li>Else if study treatment start date is not missing:<ul><li>If year of start date = year of study treatment start date, then<ul><li>If stop date contains a full date and stop date is earlier than study treatment start date, then set start date = January 1.</li><li>Else set start date = study treatment start date.</li></ul></li></ul></li><li>Else set start date = January 1.</li></ul></td></tr>
<tr><td>Missing end day</td><td>A '28/29/30/31' will be used for the day (dependent on the month and year)</td></tr>
<tr><td>Missing end day and month</td><td>No Imputation</td></tr>
<tr><td>Completely missing start/end date</td><td>No imputation</td></tr>
</table>
</td></tr>
<tr><td>Concomitant Medications</td><td>
<ul><li>Partial dates for any concomitant medications recorded in the CRF will be imputed using the following convention:</li></ul>
<table>
<tr><td>Missing start day</td><td><ul><li>If study treatment start date is missing (i.e. subject did not start study treatment), then set start date = 1st of month.</li><li>Else if study treatment start date is not missing:<ul><li>If month and year of start date = month and year of study treatment start date, then<ul><li>If stop date contains a full date and stop date is earlier than study</li></ul></li></ul></li></ul></td></tr>
</table>
</td></tr>
</table>

| Element | Reporting Detail | |
|---|---|---|
| | | treatment start date, then set start date= 1st of month.<br>▪ Else set start date = study treatment start date.<br>• Else set start date = 1st of month. |
| | Missing start day and month | • If study treatment start date is missing (i.e. subject did not start study treatment), then set start date = January 1.<br>• Else if study treatment start date is not missing:<br>o If year of start date = year of study treatment start date, then<br>▪ If stop date contains a full date and stop date is earlier than study treatment start date, then set start date = January 1.<br>▪ Else set start date = study treatment start date.<br>• Else set start date = January 1. |
| | Missing end day | A '28/29/30/31' will be used for the day (dependent on the month and year) |
| | Missing end day and month | A '31' will be used for the day and 'Dec' will be used for the month. |
| | Completely missing start/end date | No imputation |
| Oxygen Supplementation | Completely missing end date | If the end date is missing and subsequent record exists and is not missing the start date, impute the missing end date as the start date of the subsequent record.<br><br>If end date is missing and no subsequent record exists, the method of oxygen supplementation will be considered ongoing at time of database extraction. |
| Age | Age will be imputed from year of birth. The calculation will use 30 June as the day and month and will calculate the age relative to Screening date. | |

##### 6.2.8. Statistical Modelling - Multiple Imputation

Multiple imputation will be used to impute data that is missing following withdrawal of the subject from the study, or following an intercurrent event for hypothetical estimands (if applicable), for the following two binary endpoints:

- Progression of COVID-19 through Day 29 as defined by hospitalization > 24 hours for acute management of illness due to any cause or death

- Progression of COVID-19 through Day 29 as defined by visit to a hospital emergency room for management of illness or hospitalization for acute management of illness for any duration and for any cause or death

Multiple imputation of endpoint data will be performed to impute data post study withdrawal or intercurrent event, as follows:

- Data will have a monotone missing structure so there will be no intermittent missing data.
- Imputation of missing data will proceed iteratively for each study day starting with Day 2 and ending at Day 29 using a logistic regression imputation model including the analysis model covariates and a parameter for COVID-19 progression status for the previous study day, i.e. imputation of COVID-19 progression status at Day X will include a covariate for status at Day X-1 from the previous imputation step.
- The “likelihood=augment” option in PROC MI will be used in the imputation to avoid quasi complete separation of data for the preceding timepoint covariate included in the imputation model. If logistic regression step fails to converge the following back-up options may be used:
    i) A discriminant function [Brand, 1999] may be used in place of logistic.
    ii) MCMC with adaptive rounding.

A sufficient number of imputations will be performed to ensure the stability of the estimates. The results of analysis from each complete imputed dataset will be combined using Rubin’s rule.

Table 3 Multiple Imputation Specifications) details the models, covariates and the seed to be used for the analyses.

**Table 3 Multiple Imputation Specifications**

| Endpoint | Model | Covariates | Post-Baseline Timepoints Included in Prediction Model for Day X (X=2,…,29) | Initial Seed |
|---|---|---|---|---|
| Proportion of participants who have progression of COVID-19 at Day 29 (hospitalized >24h/death) | Monotone Binary Logistic Regression | Treatment, Age Group at Baseline, Sex | Day X-1 | 6862 |
| Proportion of participants who have progression of COVID-19 at Day 29 (ER visit/hospitalized/death) | Monotone Binary Logistic Regression | Treatment, Age Group at Baseline, Sex | Day X-1 | 376 |

The seeds were generated using the following code:

```
DATA seeds;
DO i=1 to 3;
seed=int(10000*ranuni(214366));
```

```
OUTPUT;
END;
RUN;
```

The same seed is used for all estimands and sensitivity analyses. If additional seeds are required, then the initial seed as specified will be incremented by 1.

##### 6.2.9. Count Data Model Checking

Distributional assumptions underlying the model used for analysis will be checked. If there are any departures from the distributional assumptions, alternative models may be explored.

##### 6.2.10. Continuous Data Model Checking

Distributional assumptions underlying the model used for analysis will be examined by obtaining a normal probability plot of the residuals and a plot of the residuals versus the fitted values (*i.e.* checking the normality assumption and constant variance assumption of the model respectively) to gain confidence that the model assumptions are reasonable.

If there are any departures from the distributional assumptions, alternative models will be explored using appropriate transformed data.

##### 6.2.11. Early PK Access Key Activities

PK dummy activities will be performed as per the standard process. The details of dummy activities will be explained in the OPS

##### 6.2.12. Trademarks

| Trademarks of the GlaxoSmithKline Group of Companies |
|---|
| None |

| Trademarks not owned by the GlaxoSmithKline Group of Companies |
|---|
| None |
